# Global, Regional, and National Burden of Pulmonary Arterial Hypertension, 1990-2021: a Systematic Analysis of the Global Burden of Disease Study 2021

**DOI:** 10.1101/2024.10.25.24316118

**Authors:** Yafang Deng, Xingbei Dong, Junyan Qian, Jiuliang Zhao, Dong Xu, Yanhong Wang, Mengtao Li, Xiaofeng Zeng, Qian Wang

## Abstract

**Aims:** Pulmonary arterial hypertension (PAH) is a severe pulmonary vascular disease with high mortality rates. This research aimed to examine the disease burden of PAH including prevalence, incidence, mortality, and disability-adjusted life-years (DALYs) from 1990 to 2021 across various age groups, sexes, and Socio-demographic Index (SDI) levels, at global, regional and national levels.

**Methods:** Data was collected from the Global Burden of Diseases, Injuries, and Risk Factors study (GBD) 2021, which established models to estimate the burden of PAH for 204 countries and territories from 1990 to 2021. Age-standardized rates for PAH prevalence, incidence, mortality, and DALYs were calculated. Joinpoint regression was used to assess temporal trends. The correlation between disease burden and socioeconomic development was analyzed.

**Results:** In 2021, the global age-standardized prevalence and incidence rates were 2.28 and 0.52 per 100,000 population, respectively. From 1990 to 2021, the age-standardized mortality rate decreased from 0.35 to 0.27 per 100,000, with an average annual trend of -0.82%. The **Corresponding Author** Dr. Qian Wang, Department of Rheumatology and Clinical Immunology, Peking Union Medical College Hospital, No. 1, Shuaifuyuan, Wangfujing Ave, Dongcheng District, Beijing 100730, China E-Mail: age-standardized DALY rate decreased from 13.21 to 8.24 per 100,000 by -1.52%. Females exhibited a higher incidence rate but lower mortality and DALY rates. The burden of PAH distributed unequally, with regions possessing higher socioeconomic statuses exhibiting lower mortality and DALY rates. Such inequalities have been improved over past decades.

**Conclusions:** The decline in mortality and DALY rates of PAH indicates improvements in disease management and patient outcomes, while the health inequalities among different regions and countries persist. Improving survival and life quality, and global health equality are crucial for future progress.

## Introduction

Pulmonary arterial hypertension (PAH) is a life-threatening disease characterized by extensive pulmonary vasoconstriction and pulmonary vascular remodeling, and the progressive increased pulmonary vascular resistance eventually leads to right ventricular failure. ^1,2^ Despite existing studies on the disease burden of PAH across diverse geographies and etiologies, ^3^ a comprehensive, global investigation remains lacking. Prevailing studies report a mean prevalence ranging from 3 to 5 cases per 100,000 individuals, with incidence rates fluctuating between 0.40 to 0.58 cases per 100,000 person-years according to systematic reviews. ^4,5^ Such variation is often ascribed to the heterogeneity of included studies and the inconsistent application of diagnostic criteria. With the continuous changes in global demographics, the evolving definitions and classifications of PAH, and dynamic therapeutic landscape, ^6,7^ it is essential to systematically evaluate the contemporary global disease burden.

The Global Burden of Disease (GBD) study is the largest worldwide program to quantify health loss across global regions and over time, aiming to improve health outcomes. ^8^ The latest GBD database notably includes, for the first time, comprehensive data on the burden of PAH from 1990 to 2021. ^9^ Such data is instrumental for advancing our understanding of PAH’s epidemiological trends. In this study, we used the GBD database to investigate PAH associated prevalence, incidence, mortality, and disability adjusted life years (DALYs) at global, regional, and national level from 1990 to 2021, stratified by age, sex, and sociodemographic development. This comprehensive analysis of the global PAH burden is expected to inform future strategies for disease screening, surveillance, and management.

## Methods

### Overview and data source

The GBD 2021 study encompassed a comprehensive spectrum of 371 diseases and injuries, along with 88 risk factors, across 204 countries and territories. This study provides an in-depth look at the health metrics including prevalence, incidence, mortality, DALYs, and age-standardized rates (ASRs) by cause, age, sex, year, and location. ^9,10^ For this research, data on the global, regional and national disease burden of PAH from 1990 to 2021 was collected from The Global Health Data Exchange GBD Results Tool (https://vizhub.healthdata.org/gbd-results/).

### Disease definition

PAH in this study is limited to only the World Health Organization’s (WHO) Group 1 pulmonary hypertension. ^3^ The diagnosis of PAH was established through either right heart catheterization or echocardiography and was confirmed by physician.

### Statistical analysis

Numbers, crude rates, ASRs including age-standardized prevalence rates (ASPRs), age-standardized incidence rates (ASIRs), age-standardized mortality rates (ASMRs), and age-standardized disability-adjusted life-year rates (ASDRs), and 95% uncertainty interval (95% UI) data were extracted from GBD. ASRs were calculated with the following formula: 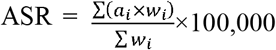, in which *i* denotes the *i*^th^ age group, *a*_*i*_ indicates the age-specific rates in this group, and *w*_*i*_ represents the population weight for the corresponding age group in the reference standard population. DALYs were calculated as the sum of years lived with disability (YLD) and years of life lost (YLL).

The joinpoint regression model was utilized to examine the linear trends in the disease burden of PAH from 1990 to 2021. This model employs the least squares method to minimize potential subjectivity in trend estimation. The average annual percent change (AAPC) was calculated to quantify significant trends using the following formula: 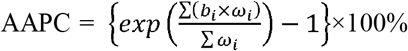, in which *i* denotes the *i*^th^ desired segment of the time series, *b*_*i*_ indicates the slope coefficient of this segment, and *w*_*i*_ represents the duration of the segment in years. A positive AAPC, with both the estimate and its 95% confidence interval (CI) above zero, indicates an increasing trend, while a negative AAPC, with both values below zero, signifies a decreasing trend for the specific variable. The Socio-demographic Index (SDI) serves as a comprehensive measure of a country or region’s developmental progress, taking into account a spectrum of socioeconomic factors including income equality, educational levels, and fertility rates among young females. ^9^ With a scale ranging from 0 to 1, a higher SDI value indicates a more advanced state of socioeconomic development. The correlation between the disease burden of PAH and socioeconomic development in different countries and regions was analyzed to evaluate global health inequalities. The slope index of inequality (SII) was employed to quantify absolute inequalities by measuring the gradient of the regression line between the ASDR of PAH and weighted ranking of each country and territory. ^11^

All data analysis and visualization were performed using Joinpoint Regression Program (version 5.1.0.0, Statistical Research and Applications Branch, National Cancer Institute, USA) and the open-source software R (version 4.3.2; R Foundation for Statistical Computing, Vienna, Austria).

## Results

### Global trends

In 2021, the global prevalence of PAH was 191.81×10^3^ cases (95% UI, 155.36×10^3^ to 235.79×10^3^), increased by 81.47% compared to which of 1990 (105.7×10^3^, 95% UI 86.38×10^3^ to 130.33×10^3^). However, the ASPR of PAH showed a slight decline between 1990 and 2021 from 2.30/100,000 (95% UI 1.87/100,000 to 2.82/100,000) to 2.28/100,000 (95% UI 1.85/100,000 to 2.80/100,000). The ASIR of PAH was 0.50/100,000 (95% UI 0.40/100,000 to 0.60/100,000) in 1990 and increased to 0.52/100,000 (95% UI 0.42/100,000 to 0.62/100,000) in 2021, with an AAPC of 0.10% (95% CI, 0.10% to 0.11%) (Table 1).

**Table 1.** Global incidence, deaths, and DALYs and their annual trends of pulmonary arterial hypertension (PAH).

|  | Incidence (95% UI) |  |  | Deaths (95% UI) |  |  | DALYs (95% UI) |  |  |
| --- | --- | --- | --- | --- | --- | --- | --- | --- | --- |
|  | Incident cases in 2021 (000s) | ASIR in 2021 (per 100,000) | AAPC of ASIR (95% CI) | Death cases in 2021 (000s) | ASMR in 2021 (per 100,000) | AAPC of ASMR (95% CI) | DALY cases in 2021 (000s) | ASDR in 2021 (per 100,000) | AAPC of ASDR (95% CI) |
| **Global** | 43.25 (34.7 to 52.44) | 0.52 (0.42 to 0.62) | 0.10 (0.10 to 0.11) | 22.02 (18.24 to 25.35) | 0.27 (0.23 to 0.32) | -0.82 (-0.95 to -0.68) | 642.1 (552.27 to 728.99) | 8.24 (7.14 to 9.39) | -1.52 (-1.64 to -1.40) |
| **Sex** |  |  |  |  |  |  |  |  |  |
| Female | 23.42 (18.89 to 28.35) | 0.55 (0.45 to 0.66) | 0.12 (0.11 to 0.14) | 12.44 (10 to 15.38) | 0.28 (0.22 to 0.34) | -0.61 (-0.73 to -0.49) | 342.47 (282.65 to 430.64) | 8.39 (6.92 to 10.53) | -1.26 (-1.37 to -1.15) |
| Male | 19.83 (15.82 to 24.09) | 0.48 (0.39 to 0.58) | 0.08 (0.08 to 0.09) | 9.58 (7.52 to 11.7) | 0.27 (0.21 to 0.33) | -1.04 (-1.17 to -0.92) | 299.64 (246.73 to 348.33) | 8.06 (6.72 to 9.36) | -1.80 (-1.90 to -1.70) |
| **SDI levels** |  |  |  |  |  |  |  |  |  |
| High SDI | 5.61 (4.5 to 6.96) | 0.37 (0.29 to 0.44) | -0.02 (-0.03 to -0.01) | 4.62 (3.92 to 5.05) | 0.22 (0.19 to 0.23) | -0.59 (-0.75 to -0.44) | 93.18 (84.87 to 99.19) | 6.16 (5.76 to 6.49) | -1.29 (-1.45 to -1.14) |
| High-middle SDI | 7.73 (6.19 to 9.59) | 0.46 (0.37 to 0.55) | 0.19 (0.17 to 0.21) | 4.33 (3.59 to 5.14) | 0.24 (0.2 to 0.29) | -1.22 (-1.50 to -0.95) | 99.45 (85.76 to 117.64) | 6.48 (5.61 to 7.87) | -2.27 (-2.48 to -2.07) |
| Middle SDI | 14.17 (11.32 to 17.38) | 0.53 (0.43 to 0.64) | -0.02 (-0.03 to -0.01) | 7.55 (5.14 to 9.03) | 0.33 (0.22 to 0.39) | -1.06 (-1.19 to -0.93) | 197.17 (148.78 to 232.32) | 8.23 (6.26 to 9.7) | -1.78 (-1.92 to -1.64) |
| Low-middle SDI | 9.98 (8.09 to 12.02) | 0.59 (0.47 to 0.7) | -0.07 (-0.10 to -0.03) | 3.73 (2.76 to 5.09) | 0.26 (0.18 to 0.38) | -0.89 (-1.03 to -0.75) | 156.4 (122.43 to 194.17) | 9.07 (7.05 to 11.6) | -1.59 (-1.70 to -1.47) |
| Low SDI | 5.71 (4.68 to 6.85) | 0.71 (0.58 to 0.85) | -0.30 (-0.33 to -0.27) | 1.78 (1.15 to 2.54) | 0.27 (0.15 to 0.40) | -0.53 (-0.81 to -0.25) | 95.34 (67.47 to 133.05) | 9.3 (6.08 to 13.2) | -0.92 (-1.04 to -0.80) |
| **Age** |  |  |  |  |  |  |  |  |  |
| <5 years | 1.20 (0.89 to 1.63) | 0.18 (0.14 to 0.25) | 0.11 (0.10 to 0.12) | 1.41 (1.08 to 1.78) | 0.21 (0.16 to 0.27) | -3.16 (-3.37 to -2.94) | 125.96 (96.51 to 159.4) | 19.14 (14.66 to 24.22) | -3.16 (-3.37 to -2.95) |
| 5-9 years | 1.26 (0.91 to 1.68) | 0.18 (0.13 to 0.24) | 0.11 (0.10 to 0.12) | 0.15 (0.12 to 0.19) | 0.02 (0.02 to 0.03) | -2.07 (-2.31 to -1.84) | 12.81 (10.35 to 15.87) | 1.86 (1.51 to 2.31) | -2.04 (-2.27 to -1.81) |

| Age | Incident cases in 2021 (000s) | Incidence rate in 2021 (per 100,000) | AAPC of ASIR (95% CI) | Death cases in 2021 (000s) | Mortality rate in 2021 (per 100,000) | AAPC of ASMR (95% CI) | DALY cases in 2021 (000s) | DALY rate in 2021 (per 100,000) | AAPC of ASDR (95% CI) |
| --- | --- | --- | --- | --- | --- | --- | --- | --- | --- |
| 10-14 years | 1.24 (0.74 to 1.93) | 0.19 (0.11 to 0.29) | 0.14 (0.13 to 0.15) | 0.15 (0.12 to 0.19) | 0.02 (0.02 to 0.03) | -1.34 (-1.58 to -1.10) | 12.33 (9.82 to 15.35) | 1.85 (1.47 to 2.3) | -1.31 (-1.54 to -1.07) |
| 15-19 years | 1.34 (0.87 to 2.04) | 0.21 (0.14 to 0.33) | 0.21 (0.17 to 0.24) | 0.26 (0.21 to 0.33) | 0.04 (0.03 to 0.05) | -1.04 (-1.18 to -0.91) | 19.27 (15.49 to 24.75) | 3.09 (2.48 to 3.97) | -1.02 (-1.15 to -0.88) |
| 20-24 years | 1.59 (1.03 to 2.35) | 0.27 (0.17 to 0.39) | 0.21 (0.19 to 0.23) | 0.33 (0.26 to 0.42) | 0.05 (0.04 to 0.07) | -0.76 (-0.82 to -0.71) | 22.87 (18.25 to 28.91) | 3.83 (3.06 to 4.84) | -0.71 (-0.80 to -0.61) |
| 25-29 years | 1.85 (1.03 to 2.86) | 0.31 (0.18 to 0.49) | 0.19 (0.18 to 0.20) | 0.42 (0.35 to 0.51) | 0.07 (0.06 to 0.09) | -0.90 (-0.98 to -0.82) | 27.21 (22.50 to 32.56) | 4.62 (3.82 to 5.53) | -0.88 (-0.95 to -0.80) |
| 30-34 years | 2.32 (1.48 to 3.42) | 0.38 (0.25 to 0.57) | 0.15 (0.13 to 0.17) | 0.4 (0.34 to 0.48) | 0.07 (0.06 to 0.08) | -1.03 (-1.21 to -0.85) | 24.18 (20.20 to 28.86) | 4.00 (3.34 to 4.77) | -0.99 (-1.16 to -0.81) |
| 35-39 years | 2.69 (1.79 to 3.89) | 0.48 (0.32 to 0.69) | 0.14 (0.13 to 0.16) | 0.48 (0.41 to 0.56) | 0.09 (0.07 to 0.10) | -1.14 (-1.28 to -0.99) | 26.53 (22.78 to 30.79) | 4.73 (4.06 to 5.49) | -1.09 (-1.23 to -0.95) |
| 40-44 years | 2.87 (1.59 to 4.57) | 0.57 (0.32 to 0.91) | 0.15 (0.13 to 0.17) | 0.62 (0.52 to 0.73) | 0.12 (0.10 to 0.15) | -1.17 (-1.37 to -0.97) | 30.89 (26.22 to 35.88) | 6.17 (5.24 to 7.17) | -1.13 (-1.32 to -0.94) |
| 45-49 years | 3.33 (2.07 to 4.91) | 0.70 (0.44 to 1.04) | 0.08 (0.07 to 0.09) | 0.52 (0.45 to 0.62) | 0.11 (0.09 to 0.13) | -1.26 (-1.41 to -1.10) | 23.74 (20.37 to 27.85) | 5.01 (4.30 to 5.88) | -1.20 (-1.34 to -1.05) |
| 50-54 years | 3.86 (2.57 to 5.59) | 0.87 (0.58 to 1.26) | 0.08 (0.07 to 0.09) | 0.74 (0.62 to 0.86) | 0.17 (0.14 to 0.19) | -1.40 (-1.57 to -1.23) | 29.66 (25.33 to 34.27) | 6.67 (5.69 to 7.70) | -1.34 (-1.52 to -1.15) |
| 55-59 years | 4.08 (2.43 to 6.40) | 1.03 (0.61 to 1.62) | 0.05 (0.02 to 0.08) | 1.10 (0.86 to 1.32) | 0.28 (0.22 to 0.33) | -1.13 (-1.30 to -0.97) | 38.59 (30.61 to 46.13) | 9.75 (7.74 to 11.66) | -1.08 (-1.24 to -0.92) |
| 60-64 years | 3.89 (2.56 to 5.67) | 1.22 (0.80 to 1.77) | 0.07 (0.05 to 0.09) | 1.15 (0.91 to 1.34) | 0.36 (0.28 to 0.42) | -1.17 (-1.31 to -1.04) | 34.71 (27.82 to 39.95) | 10.85 (8.69 to 12.48) | -1.13 (-1.26 to -1.01) |
| 65-69 years | 3.94 (2.85 to 5.29) | 1.43 (1.03 to 1.92) | 0.07 (0.05 to 0.09) | 1.75 (1.38 to 2.08) | 0.63 (0.50 to 0.75) | -0.95 (-1.08 to -0.81) | 44.13 (34.81 to 52.09) | 16.00 (12.62 to 18.88) | -0.92 (-1.03 to -0.81) |
| 70-74 years | 3.32 (2.19 to 4.84) | 1.61 (1.06 to 2.35) | 0.03 (-0.01 to 0.07) | 2.23 (1.69 to 2.66) | 1.08 (0.82 to 1.29) | -0.75 (-0.89 to -0.61) | 46.04 (34.97 to 54.86) | 22.37 (16.99 to 26.65) | -0.73 (-0.86 to -0.60) |
| 75-79 years | 2.19 (1.43 to 3.15) | 1.66 (1.09 to 2.39) | 0.12 (0.10 to 0.13) | 2.55 (1.88 to 3.07) | 1.93 (1.43 to 2.33) | -0.46 (-0.78 to -0.14) | 41.58 (31.08 to 49.95) | 31.53 (23.57 to 37.88) | -0.47 (-0.76 to -0.18) |
| 80-84 years | 1.34 (0.95 to 1.82) | 1.53 (1.09 to 2.08) | 0.09 (0.07 to 0.11) | 2.79 (2.07 to 3.39) | 3.19 (2.36 to 3.87) | -0.15 (-0.38 to 0.09) | 35.42 (26.37 to 42.82) | 40.44 (30.11 to 48.89) | -0.13 (-0.35 to 0.08) |
| 85-89 years | 0.63 (0.39 to 0.99) | 1.38 (0.86 to 2.18) | 0.07 (0.05 to 0.10) | 2.62 (2.03 to 3.07) | 5.72 (4.43 to 6.72) | 0.28 (0.10 to 0.47) | 26.17 (20.30 to 30.62) | 57.24 (44.39 to 66.97) | 0.26 (0.08 to 0.45) |
| 90-94 years | 0.24 (0.14 to 0.38) | 1.34 (0.76 to 2.15) | 0.09 (0.07 to 0.10) | 1.63 (1.27 to 1.92) | 9.13 (7.10 to 10.74) | 0.60 (0.40 to 0.80) | 14.18 (11.02 to 16.65) | 79.27 (61.63 to 93.09) | 0.60 (0.40 to 0.79) |
| 95+ years | 0.08 (0.04 to 0.14) | 1.39 (0.67 to 2.64) | 0.06 (0.03 to 0.10) | 0.72 (0.52 to 0.85) | 13.19 (9.51 to 15.60) | 0.91 (0.78 to 1.05) | 5.82 (4.20 to 6.87) | 106.77 (77.07 to 126.03) | 0.88 (0.75 to 1.01) |

DALYs, disability-adjusted life-years; ASIR, age-standardized incidence rate; ASMR, age-standardized mortality rate; ASDR, age-standardized disability-adjusted life-year rate; AAPC, average annual percentage change; UI, uncertainty interval; CI, confidence interval.

Despite an increase in the number of death cases, the ASMR in 2021 (0.27/100,000, 95% UI 0.23/100,000 to 0.32/100,000) was lower than which in 1990 (0.35/100,000, 95% UI 0.29/100,000 to 0.42/100,000), with an AAPC of -0.82% (95% CI -0.95% to -0.68%) between 1990 and 2021. A similar and more pronounced trend was observed in the ASDR, which decreased from 13.21/100,000 (95% UI 10.78/100,000 to 15.36/100,000) in 1990 to 8.24/100,000 (95% UI 7.14/100,000 to 9.39/100,000) in 2021, with an AAPC of -1.52% (95% CI -1.64% to -1.40%) (Table 1 and Structured Graphical Abstract).

### Age and sex patterns

In both 1990 and 2021, the incidence of PAH was generally higher in females than in males. The ASIR in 2021 was 0.48/100,000 (95% UI 0.39/100,000 to 0.58/100,000) for males and 0.55/100,000 (0.45/100,000 to 0.66/100,000) for females. The AAPC in ASIR from 1990 to 2021 was 0.08% (95% CI 0.08% to 0.09%) for males and 0.12% (95% CI, 0.11% to 0.14%) for females, indicating rising trends in PAH incidence across both sexes (Table1 and Supplementary Fig 1A). In 2021, the highest incident cases and rates were observed in the 55-59 years age group and the 75-79 years age group, respectively for both sexes, and the incidence rate increased with age in the population under 75 years. Across all age groups, the incidence rate in females consistently exceeded that in males (Fig 1A).

**Figure 1.**
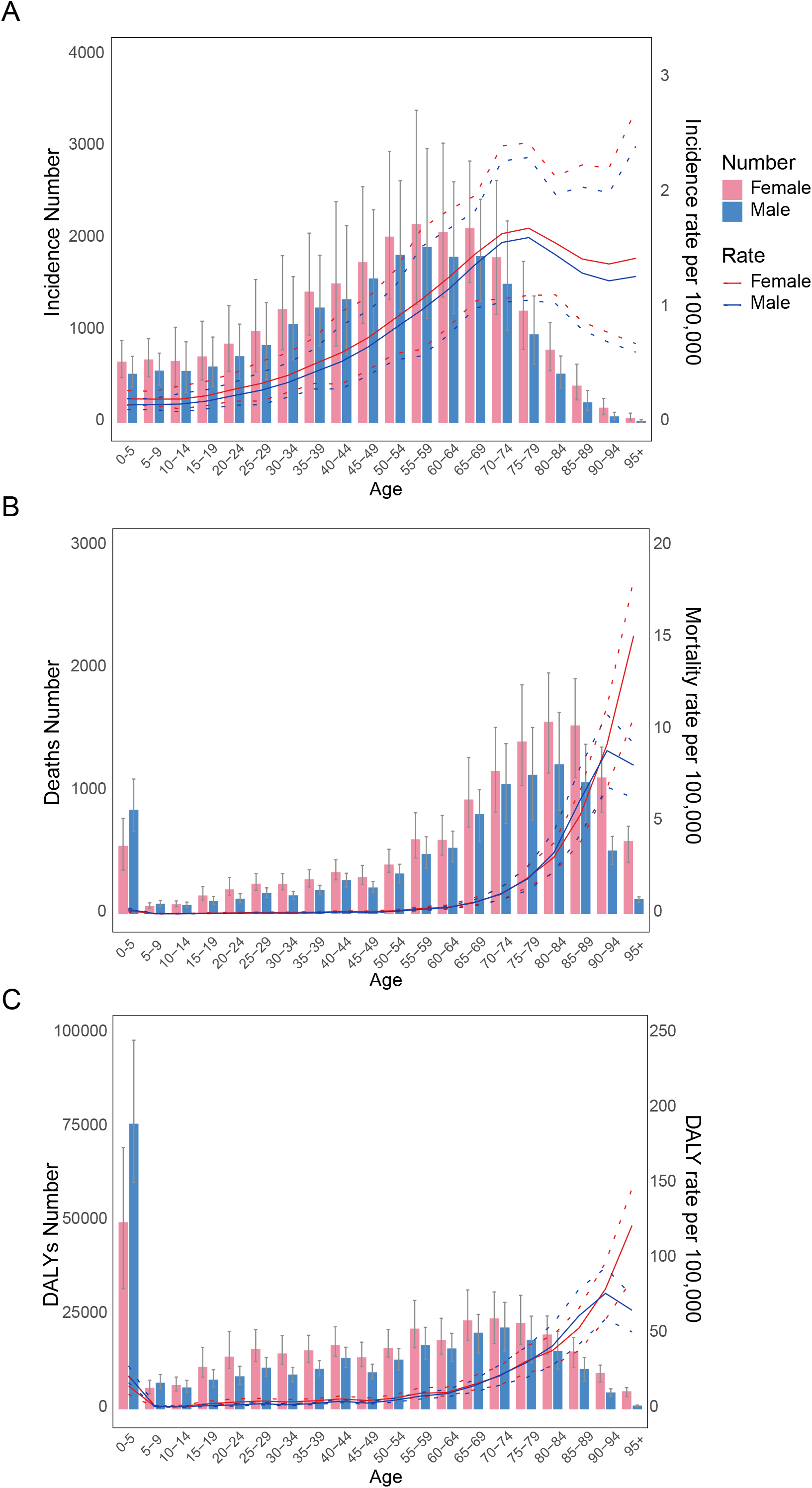
Global numbers and rates of incidence, deaths and DALYs of PAH by age and sex in 2021. The dotted lines indicate 95% upper and lower uncertainty intervals. DALY, disability-adjusted life-year. (A) The numbers and rates of incidence. (B) The numbers and rates of deaths. (C) The numbers and rates of DALYs.

In 2021, females exhibited a higher number of death cases and a greater ASMR compared to males. However, females typically had lower ASMRs in most years, as depicted in Supplementary Fig 1b. The ASMR declined more markedly in males than in females between 1990 and 2021, with an AAPC of -1.04% (95% CI -1.17% to -0.92%) for males and -0.61% (95% CI -0.73% to -0.49%) for females (Table 1). Within the patient population aged 5 to 80 years, both the number of death cases and the mortality rate increased with age for both sexes. Notably, children aged 0 to 5 years represented a significant part of death cases, with 1.41×10^3^ cases (95% UI 1.08×10^3^ to 1.78×10^3^) (Fig 1B). YLL constituted the majority of DALYs for PAH, as shown in Supplementary Fig 2. Consistent with mortality trends, females generally presented lower ASDRs than males over the past three decades (Supplementary Fig 1C). Declining trends in ASDRs were observed for both sexes from 1990 to 2021, with a more significant decrease in males, as indicated by an AAPC of -1.80% (95% CI -1.90% to -1.70%) compared to -1.26% (95% CI -1.37% to -1.15%) in females (Table 1). Children under 5 years contributed the most to the total DALY numbers in both sexes, and the DALY rates increased with age in populations over 5 years old (Fig 1C). From 1990 to 2021, most age groups showed downward trends in mortality and DALY rates, particularly in those under 5 years, with AAPCs of -3.16% (95% CI -3.37% to -2.94%) for mortality rate and -3.16% (95% CI -3.37% to -2.95%) for DALY rate. However, in individuals over 85 years, both mortality and DALY rates continued to increase from 1990 to 2021 (Table 1).

### Regional trends

Among the 21 regions analyzed, the ASIR of PAH in 2021 varied from 0.30/100,000 (95% UI 0.24/100,000 to 0.37/100,000) in High-income North America to 0.92/100,000 (95% UI 0.75/100,000 to 1.09/100,000) in Eastern Sub-Saharan Africa (Supplemental Table 2). Generally, regions with higher SDI, such as High-income North America, High-income Asia Pacific, and Australasia exhibited lower incidence rates, while regions with lower socioeconomic development, including Central, Western, and Eastern Sub-Saharan Africa, had higher rates (Fig 2A). Temporal trends in incidence rates differed by region. Central Europe experienced the most rapid increase in ASIR, with an AAPC of 0.60% (95% CI 0.57% to 0.63%), whereas Western Sub-Saharan Africa showed the most significant decline, with an AAPC of -0.84% (95% CI -0.90% to -0.79%) (Supplemental Table 2). In most regions, the incidence rates over the past three decades showed moderate fluctuations (Fig 2A).

**Figure 2.**
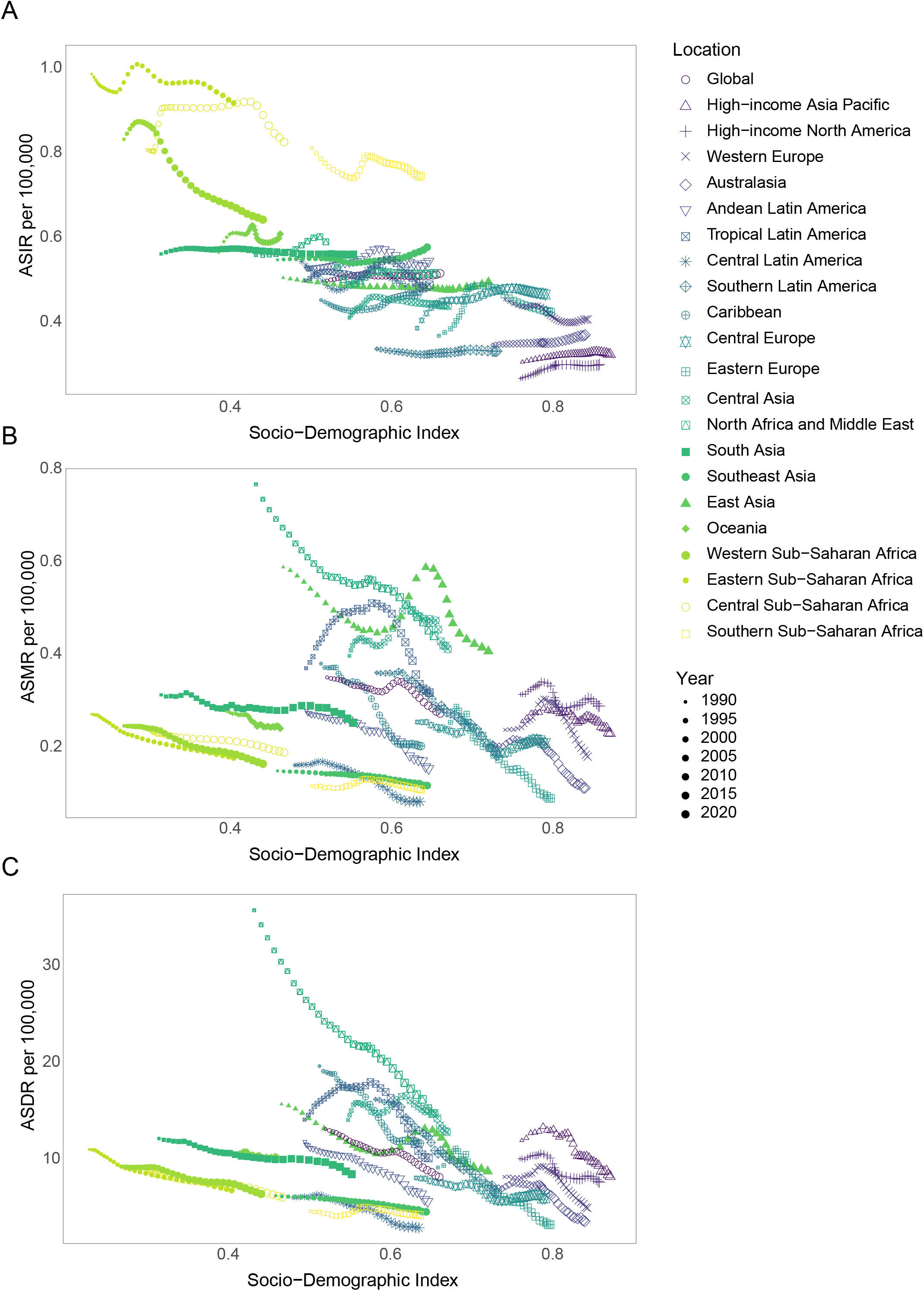
Age-standardized rates of PAH in global and 21 GBD regions by SDI from 1990 to 2021. 32 continuous points plotted for each GBD region represent the age-standardized rates from 1990 to 2021 for that region. (A) Disease burden of ASIRs. (B) Disease burden of ASMRs. (C) Disease burden of ASDRs. GBD, Global Burden of Disease; SDI, Socio-Demographic Index; ASIR, age-standardized incidence rate; ASMR, age-standardized mortality rate; ASDR, age-standardized disability-adjusted life-year rate.

In 2021, the highest ASMR and ASDR for PAH were observed in North Africa and Middle East, at 0.44/100,000 (95% UI 0.31/100,000 to 0.53/100,000) and 14.81/100,000 (95% UI 10.76/100,000 to 17.96/100,000), respectively. In contrast, Central Latin America reported the lowest rates, with an ASMR of 0.08/100,000 (95% UI 0.07/100,000 to 0.10/100,000) and an ASDR of 3.00/100,000 (95% UI 2.63/100,000 to 3.51/100,000) (Supplemental Table 3 and Supplemental Table 4). Between 1990 and 2021, all regions except Central Asia, which showed a non-significant trend, experienced significant declines in both ASMR and ASDR. The most significant decreases were noted in Eastern Europe, with an AAPC of ASMR at -2.98% (95% CI -3.74% to -2.21%) and an AAPC of ASDR at -3.22% (95% CI, -4.10% to -2.34%) (Supplemental Table 3 and Supplemental Table 4). In most regions, there was a general trend of decreasing age-standardized mortality and DALY rates with increasing SDI, although some regions exhibited considerable variations (Fig 2B and Fig 2C).

### National trends

In 2021, the ASIRs of PAH varied significantly across countries and territories, ranging from 0.18 to 1.06 per 100,000 persons. Zambia reported the highest ASIR at 1.06/100,000 (95% UI 0.88/100,000 to 1.28/100,000), followed by Uganda (1.00/100,000, 95% UI 0.82/100,000 to 1.20/100,000), and Ethiopia (1.00/100,000, 95% UI 0.82/100,000 to 1.19/100,000). In contrast, the lowest rates were observed in Ireland (0.18/100,000, 95% UI 0.15/100,000 to 0.23/100,000), Canada (0.26/100,000, 95% UI 0.21/100,000 to 0.32/100,000), and Portugal (0.28/100,000, 95% UI 0.23/100,000 to 0.34/100,000) (Supplemental Table 2 and Supplemental Fig 3A). From 1990 to 2021, about half of the countries and territories experienced an increase in age-standardized incidence rates (Supplemental Fig 4A). Slovakia experienced the most substantial increase (AAPC 1.07%, 95% CI 0.95% to 1.19%), while Senegal showed the greatest decrease (AAPC -1.61%, 95% CI -1.70% to -1.52%). Among 204 countries and territories, an inverse correlation was observed between the SDI and the incidence rate, aligning with the patterns seen in regional trends (Supplemental Fig 5A).

The age-standardized rate of mortality and DALYs also varied in distinct countries and territories. In 2021, the highest burden of disease was observed in Mongolia and Georgia, with ASMRs of 1.59/100,000 (95% UI 0.91/100,000 to 2.05/100,000) and 1.01/100,000 (95% UI 0.79/100,000 to 1.27/100,000), respectively, and ASDRs of 43.92/100,000 (95% UI 25.60/100,000 to 56.54/100,000) and 27.83/100,000 (95% UI 22.18/100,000 to 34.82/100,000), respectively. The Republic of Moldova reported the lowest ASMR at 0.01/100,000 (95% UI 0.01/100,000 to 0.01/100,000) and ASDR at 0.54/100,000 (95% UI 0.44/100,000 to 0.68/100,000), followed by Montenegro and Slovenia, which also had notably low rates (Fig 3A, Supplemental Fig 3B, Supplemental Table 3 and Supplemental Table 4). Between 1990 and 2021, approximately 90% of countries and territories demonstrated a decline in both ASMRs and ASDRs, while few of them showed upward trends, such as Latvia and Mauritius (Fig 3B and Supplemental Fig 4B). In 2021, a statistically significant negative correlation between increasing SDI and both mortality and DALY rates was observed at the national level (Supplemental Fig 5B-C).

**Figure 3.**
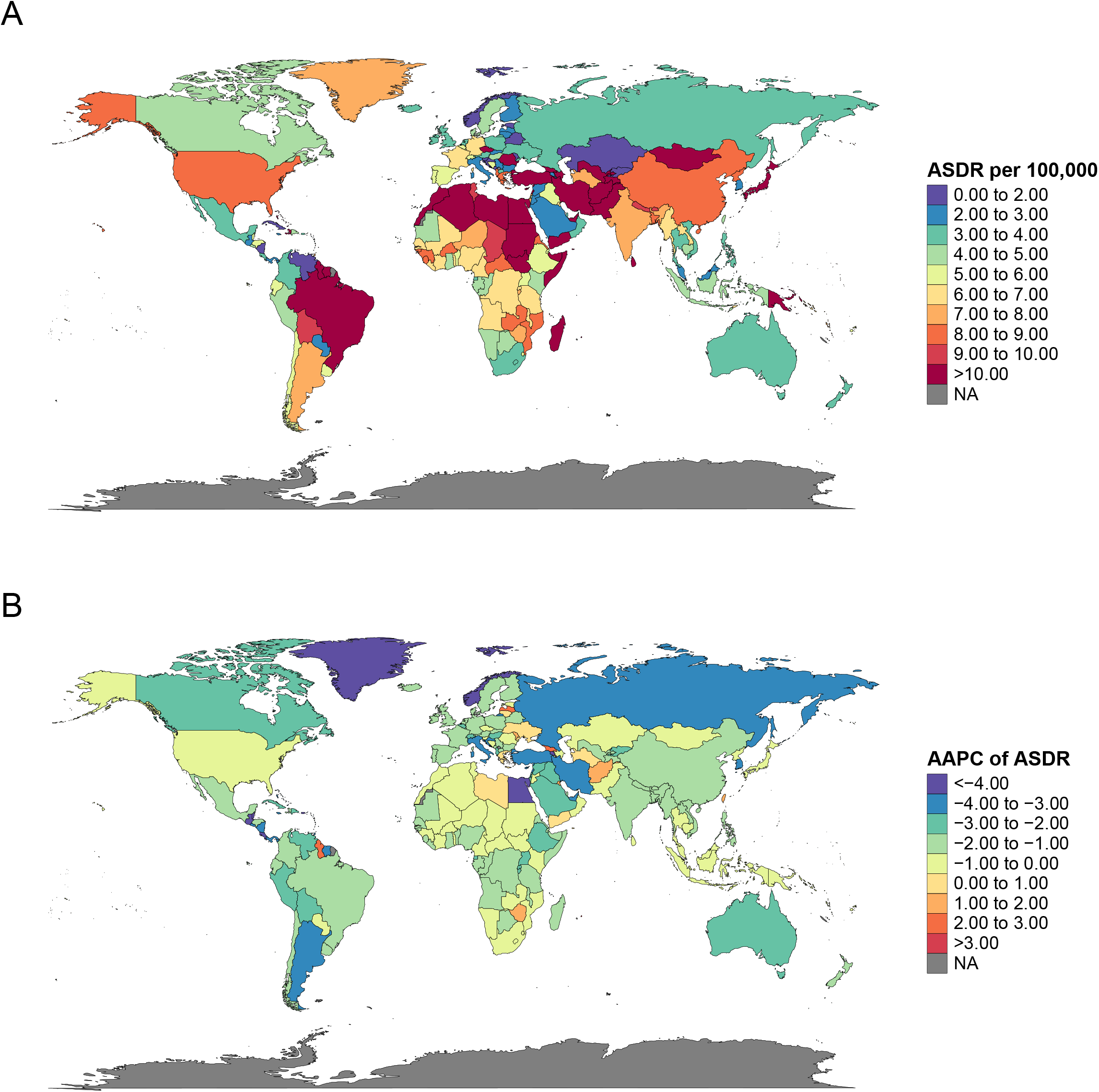
Age-standardized DALY rates and the annual trends of PAH in 204 countries and territories. (A) Disease burden of ASDRs in 2021. (B) The average annual percent changes of ASDRs between 1990 and 2021.

**Figure 4.**
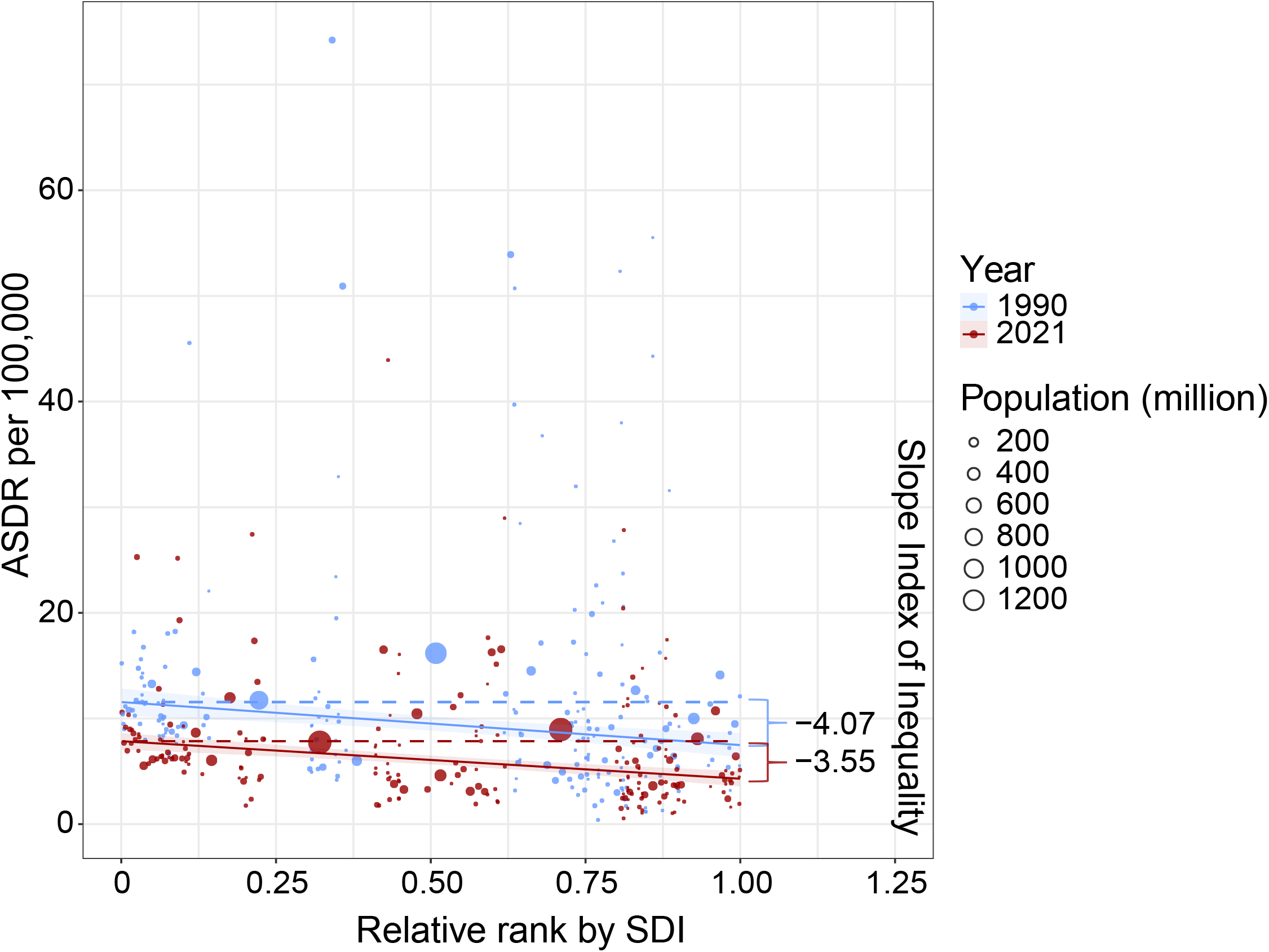
The inequality of PAH disease burden worldwide in 1990 and 2021. Absolute inequality of disease burden was evaluated by SII analysis.

### Health Inequalities

Overall, regions with lower SDI exhibited higher age-standardized incidence and DALY rates for PAH, while regions with higher socioeconomic status showed the opposite trend (Table 1). Regarding health disparities, the SII of ASDR in 1990 was -4.07 per 100,000 (95% CI -6.19 to -1.95), indicating an inverse relationship between PAH disease burden and socioeconomic development. By 2021, the SII had reduced to -3.55 per 100,000 (95% CI -4.87 to -2.23). The convergence of the SII towards zero between 1990 and 2021 indicates a more equitable global distribution of PAH burden.

## Discussion

Despite the advent of novel drugs and treatment strategies, PAH remains a life-threatening disease imposing a significant burden in the modern era. Accurate epidemiological data is challenging to obtain due to evolving diagnostic criteria and the subtle early symptoms of the disease. This study, utilizing the GBD 2021 data, provides the first comprehensive summary and analysis of the global PAH burden over the past three decades. It offers valuable insights into disease patterns, aiming to inform enhanced diagnosis and management strategies for the future.

The overall prevalence and incidence rates of PAH observed in this study are consistent with data from other registries, ^12-15^ and have remained relatively stable over the past three decades. According to GBD 2021, both age-standardized prevalence and incidence rate of PAH were higher in females than in males, which is also observed across many other cohorts. ^16-18^ The higher incidence of PAH in females may be attributed to distinctions in estrogen metabolism, function, and receptor expression, ^19^ as well as autoimmune factors and variable penetrance of gene mutations in specific PAH subtypes. ^20,21^ In this study, females have generally exhibited lower age-standardized mortality and DALY rates compared to males, and better survival in females was also observed in previous clinical trials. ^22^ This survival advantage may be due to superior right ventricular contractility and adaptation and better response to treatment in females, potentially enhanced by the protective effects of estrogen during the disease process. ^19,23^

Our study observed a shift in the median age of PAH diagnosis from 45-49 years in 1990 to 50-54 years by 2021, a trend consistent with previous reports. ^12,16,17,24^ This demographic shift underscores the growing need of focusing on elderly patients, who frequently present with increased cardiovascular comorbidities and higher mortality rate, and face challenges in receiving an early and accurate diagnosis. ^25^ Another age group that calls for emphasis is children under the age of 5. Pediatric PAH, predominantly idiopathic PAH and PAH associated with congenital heart disease, is particularly challenging due to the delayed diagnosis often resulting from the inability of young children to report their symptoms effectively. ^26^ Despite a decline from 1990 to 2021, the DALY burden remains significantly high in this age group, highlighting the need for improved pediatric PAH management. Future efforts should focus on non-invasive diagnostic methods, support for maintaining normal daily activities, and strategies aimed at prolonging survival. ^27^

Over the past three decades, there has been a gradual decline in the global disease burden of PAH, as measured by mortality and DALYs. The predominant component of DALYs is YLL, indicating that the reduction in life expectancy due to PAH accounts for the most significant portion of its disease burden. Therefore, improving patient survival is of great importance. Although increased disease awareness and early diagnosis also contribute to improved survival in PAH, the marked reduction in mortality and DALYs is likely primarily due to the advent of targeted therapies. Previous studies have demonstrated significant improvements in the 1-, 3-, and 5-year survival of PAH patients since the introduction of these therapies. ^17,18,28,29^ With the continuous update of high-level clinical evidence, combination therapy with targeted drugs has become the first-line recommendation in international guidelines for most of the PAH patients. ^30-33^ Moreover, beyond the traditional pathways of prostacyclin, endothelin and nitric oxide, a variety of novel therapeutic targets are emerging, with candidates such as sotatercept—a fusion protein inhibiting activin receptor type IIA signaling—and imatinib—a tyrosine kinase inhibitor—currently under clinical investigation. ^2^ For future advancements, the relatively high mortality rate in PAH necessitates further transformative innovations, and enhancing the quality of life for patients remains a critical issue.

Despite the overall decline in PAH mortality and DALYs, the disease burden remains unevenly distributed across regions and nations. Some previous researches have described health disparities in PAH diagnosis and treatment, influenced by socioeconomic status, race, and ethnicity in certain countries. ^34^ While the environmental and genetic factors associated with geographic variations in PAH incidence are not fully understood, differences in etiological distributions may contribute to the observed disparities. In Africa, PAH prevalence is often linked to HIV infection, whereas in South America, schistosomiasis is a significant cause, with these infectious diseases being more prevalent in low- and middle-income countries. ^35^ The global distribution of mortality and DALYs is of even greater concern, with the GBD 2021 indicating a general correlation between lower SDI and higher age-standardized mortality and DALY rates. One significant reason is the delayed diagnosis and treatments in underdeveloped regions, resulting in more advanced disease stages at diagnosis and poorer survival outcomes. ^36^ Lower socioeconomic status has been identified as an independent risk factor for disease progression and mortality in idiopathic PAH. ^37^ In contrast, in more developed regions, both underlying diseases and PAH are more likely to receive specialized care. Additionally, the limited availability and high cost of targeted therapies may result in suboptimal treatment in low SDI countries. Therefore, the development of region- or country-specific guidelines and consensus will be instrumental in advancing the specialized management of PAH patients. ^38^ The analysis of health disparities reveals a promising trend towards a more equitable global distribution of PAH burden in 2021 compared to 1990, underscoring the need for continued focus on this issue in clinical practice and research moving forward.

This study also has some limitations. Firstly, the majority of data in GBD database are secondary data collected from other research organizations, which may potentially lead to the underestimation of the disease burden. The further modeling and estimation process may introduce a degree of uncertainty. Secondly, the diagnostic criteria and classification systems for PAH have evolved over the years, and the difference between diagnostic methods of right heart catheterization and echocardiography also contribute to the heterogeneity. Finally, due to constraints in the available data, this study was unable to assess the disease burden associated with each subtype of PAH, and a comprehensive investigation of potential risk factors for PAH remains a task for future research.

In conclusion, this study presents the first systematic assessment of the global, regional, and national disease burden of PAH spanning the past three decades. The advancements in early diagnosis and targeted therapies have led to a significant decline in the mortality and DALYs. However, disparities in PAH burden persist across regions and countries, with health inequalities being closely linked to socioeconomic disparities. Enhancing survival, improving the quality of life, and promoting equitable access to PAH diagnosis and treatment are crucial for future progress in the field.

## Data Availability

All data produced are available online at https://vizhub.healthdata.org/gbd-results/.

## Data availability

The data used for analysis are publicly available from The Global Health Data Exchange GBD Results Tool (https://vizhub.healthdata.org/gbd-results/).

## Author contributions

YD and QW conceived and designed the study. YD, XD, and JQ conducted the literature review. YD conducted data analysis and visualization. YD, YW, and QW drafted the original manuscript. ML, XZ, and QW contributed to the funding acquisition. All authors contributed to the interpretation of findings and critical revision of the article. All authors had approved the final version and accepted responsibility to submit for publication.

## Acknowledgements

The authors would like to thank the GBD 2021 and its collaborators for providing the publicly available data.

