## Supplemental materials for "Global, Regional, and National Burden of Pulmonary Arterial Hypertension, 1990-2021: a Systematic Analysis of the Global Burden of Disease Study 2021"

**Table of contents**

| **Contents** | **Page** |
| --- | --- |
| Supplemental Table 1. Global incidence, deaths, and DALYs of pulmonary arterial hypertension (PAH) in 1990 and 2021. | 2 |
| Supplemental Table 2. Incidence of PAH between 1990 and 2021 at the global and regional levels. | 6 |
| Supplemental Table 3. Morality of PAH between 1990 and 2021 at the global and regional levels. | 17 |
| Supplemental Table 4. DALYs of PAH between 1990 and 2021 at the global and regional levels. | 27 |
| Supplemental Figure 1. Temporal trends in the all-age cases and age-standardized incidence, mortality, and DALY rates of PAH by sex from 1990 to 2021. | 37 |
| Supplemental Figure 2. Age-standardized rate of DALY, YLL and YLD of PAH from 1990 to 2021. | 38 |
| Supplemental Figure 3. Age-standardized incidence and mortality rates of PAH in 204 countries and territories in 2021. | 39 |
| Supplemental Figure 4. Annual trends of age-standardized incidence and mortality rates of PAH in 204 countries and territories from 1990 to 2021. | 40 |
| Supplemental Figure 5. Age-standardized incidence, mortality and DALY rates of PAH by 204 countries and territories and Socio-Demographic Index in 2021. | 41 |

**Supplemental Table 1. Global incidence, deaths, and DALYs of pulmonary arterial hypertension (PAH) in 1990 and 2021.**

|  | **Incidence (95% UI)** | | | | **AAPC (95% CI)** |
| --- | --- | --- | --- | --- | --- |
|  | **Incident cases in 1990 (000s)** | **ASIR in 1990 (per 100,000)** | **Incident cases in 2021 (000s)** | **ASIR in 2021 (per 100,000)** |  |
| **Global** | 23.30 (19.04 to 27.81) | 0.50 (0.40 to 0.60) | 43.25 (34.7 to 52.44) | 0.52 (0.42 to 0.62) | 0.10 (0.10 to 0.11) |
| **Sex** |  |  |  |  |  |
| Female | 12.62 (10.32 to 14.99) | 0.53 (0.43 to 0.63) | 23.42 (18.89 to 28.35) | 0.55 (0.45 to 0.66) | 0.12 (0.11 to 0.14) |
| Male | 10.68 (8.69 to 12.83) | 0.47 (0.38 to 0.57) | 19.83 (15.82 to 24.09) | 0.48 (0.39 to 0.58) | 0.08 (0.08 to 0.09) |
| **Age** |  | **Incidence rate in 1990 (per 100,000)** |  | **Incidence rate in 2021 (per 100,000)** |  |
| <5 years | 1.10 (0.81 to 1.48) | 0.18 (0.13 to 0.24) | 1.20 (0.89 to 1.63) | 0.18 (0.14 to 0.25) | 0.11 (0.10 to 0.12) |
| 5-9years | 1.04 (0.75 to 1.38) | 0.18 (0.13 to 0.24) | 1.26 (0.91 to 1.68) | 0.18 (0.13 to 0.24) | 0.11 (0.10 to 0.12) |
| 10-14 years | 0.95 (0.57 to 1.48) | 0.18 (0.11 to 0.28) | 1.24 (0.74 to 1.93) | 0.19 (0.11 to 0.29) | 0.14 (0.13 to 0.15) |
| 15-19 years | 1.05 (0.67 to 1.61) | 0.20 (0.13 to 0.31) | 1.34 (0.87 to 2.04) | 0.21 (0.14 to 0.33) | 0.21 (0.17 to 0.24) |
| 20-24 years | 1.23 (0.79 to 1.82) | 0.25 (0.16 to 0.37) | 1.59 (1.03 to 2.35) | 0.27 (0.17 to 0.39) | 0.21 (0.19 to 0.23) |
| 25-29 years | 1.31 (0.73 to 2.05) | 0.30 (0.16 to 0.46) | 1.85 (1.03 to 2.86) | 0.31 (0.18 to 0.49) | 0.19 (0.18 to 0.20) |
| 30-34 years | 1.41 (0.90 to 2.07) | 0.37 (0.23 to 0.54) | 2.32 (1.48 to 3.42) | 0.38 (0.25 to 0.57) | 0.15 (0.13 to 0.17) |
| 35-39 years | 1.61 (1.08 to 2.34) | 0.46 (0.31 to 0.67) | 2.69 (1.79 to 3.89) | 0.48 (0.32 to 0.69) | 0.14 (0.13 to 0.16) |
| 40-44 years | 1.57 (0.86 to 2.50) | 0.55 (0.30 to 0.87) | 2.87 (1.59 to 4.57) | 0.57 (0.32 to 0.91) | 0.15 (0.13 to 0.17) |
| 45-49 years | 1.59 (0.99 to 2.34) | 0.68 (0.43 to 1.01) | 3.33 (2.07 to 4.91) | 0.70 (0.44 to 1.04) | 0.08 (0.07 to 0.09) |
| 50-54 years | 1.80 (1.20 to 2.61) | 0.85 (0.56 to 1.23) | 3.86 (2.57 to 5.59) | 0.87 (0.58 to 1.26) | 0.08 (0.07 to 0.09) |
| 55-59 years | 1.88 (1.11 to 2.94) | 1.01 (0.60 to 1.59) | 4.08 (2.43 to 6.40) | 1.03 (0.61 to 1.62) | 0.05 (0.02 to 0.08) |
| 60-64 years | 1.91 (1.26 to 2.78) | 1.19 (0.78 to 1.73) | 3.89 (2.56 to 5.67) | 1.22 (0.80 to 1.77) | 0.07 (0.05 to 0.09) |
| 65-69 years | 1.73 (1.25 to 2.32) | 1.40 (1.01 to 1.88) | 3.94 (2.85 to 5.29) | 1.43 (1.03 to 1.92) | 0.07 (0.05 to 0.09) |
| 70-74 years | 1.35 (0.89 to 1.97) | 1.60 (1.05 to 2.33) | 3.32 (2.19 to 4.84) | 1.61 (1.06 to 2.35) | 0.03 (-0.01 to 0.07) |
| 75-79 years | 0.99 (0.65 to 1.42) | 1.60 (1.05 to 2.31) | 2.19 (1.43 to 3.15) | 1.66 (1.09 to 2.39) | 0.12 (0.10 to 0.13) |
| 80-84 years | 0.52 (0.37 to 0.72) | 1.48 (1.05 to 2.02) | 1.34 (0.95 to 1.82) | 1.53 (1.09 to 2.08) | 0.09 (0.07 to 0.11) |
| 85-89 years | 0.20 (0.13 to 0.32) | 1.35 (0.84 to 2.14) | 0.63 (0.39 to 0.99) | 1.38 (0.86 to 2.18) | 0.07 (0.05 to 0.10) |
| 90-94 years | 0.06 (0.03 to 0.09) | 1.30 (0.74 to 2.09) | 0.24 (0.14 to 0.38) | 1.34 (0.76 to 2.15) | 0.09 (0.07 to 0.10) |
| 95+ years | 0.01 (0.01 to 0.03) | 1.37 (0.66 to 2.58) | 0.08 (0.04 to 0.14) | 1.39 (0.67 to 2.64) | 0.06 (0.03 to 0.10) |
|  | **Deaths (95% UI)** | | | | **AAPC (95% CI)** |
|  | **Death cases in 1990 (000s)** | **ASMR in 1990 (per 100,000)** | **Death cases in 2021 (000s)** | **ASMR in 2021 (per 100,000)** |  |
| **Global** | 14.84 (12.37 to 17.48) | 0.35 (0.29 to 0.42) | 22.02 (18.24 to 25.35) | 0.27 (0.23 to 0.32) | -0.82 (-0.95 to -0.68) |
| **Sex** |  |  |  |  |  |
| Female | 7.52 (5.06 to 10.16) | 0.34 (0.23 to 0.45) | 12.44 (10 to 15.38) | 0.28 (0.22 to 0.34) | -0.61 (-0.73 to -0.49) |
| Male | 7.33 (5.92 to 8.71) | 0.37 (0.29 to 0.46) | 9.58 (7.52 to 11.7) | 0.27 (0.21 to 0.33) | -1.04 (-1.17 to -0.92) |
| **Age** |  | **Mortality rate in 1990 (per 100,000)** |  | **Mortality rate in 2021 (per 100,000)** |  |
| <5 years | 3.62 (2.04 to 5.15) | 0.58 (0.33 to 0.83) | 1.41 (1.08 to 1.78) | 0.21 (0.16 to 0.27) | -3.16 (-3.37 to -2.94) |
| 5-9years | 0.25 (0.18 to 0.32) | 0.04 (0.03 to 0.06) | 0.15 (0.12 to 0.19) | 0.02 (0.02 to 0.03) | -2.07 (-2.31 to -1.84) |
| 10-14 years | 0.19 (0.14 to 0.24) | 0.04 (0.03 to 0.04) | 0.15 (0.12 to 0.19) | 0.02 (0.02 to 0.03) | -1.34 (-1.58 to -1.10) |
| 15-19 years | 0.30 (0.23 to 0.39) | 0.06 (0.04 to 0.07) | 0.26 (0.21 to 0.33) | 0.04 (0.03 to 0.05) | -1.04 (-1.18 to -0.91) |
| 20-24 years | 0.34 (0.25 to 0.44) | 0.07 (0.05 to 0.09) | 0.33 (0.26 to 0.42) | 0.05 (0.04 to 0.07) | -0.76 (-0.82 to -0.71) |
| 25-29 years | 0.41 (0.32 to 0.52) | 0.09 (0.07 to 0.12) | 0.42 (0.35 to 0.51) | 0.07 (0.06 to 0.09) | -0.90 (-0.98 to -0.82) |
| 30-34 years | 0.35 (0.27 to 0.44) | 0.09 (0.07 to 0.11) | 0.4 (0.34 to 0.48) | 0.07 (0.06 to 0.08) | -1.03 (-1.21 to -0.85) |
| 35-39 years | 0.43 (0.34 to 0.53) | 0.12 (0.10 to 0.15) | 0.48 (0.41 to 0.56) | 0.09 (0.07 to 0.10) | -1.14 (-1.28 to -0.99) |
| 40-44 years | 0.51 (0.41 to 0.64) | 0.18 (0.14 to 0.22) | 0.62 (0.52 to 0.73) | 0.12 (0.10 to 0.15) | -1.17 (-1.37 to -0.97) |
| 45-49 years | 0.38 (0.30 to 0.47) | 0.16 (0.13 to 0.20) | 0.52 (0.45 to 0.62) | 0.11 (0.09 to 0.13) | -1.26 (-1.41 to -1.10) |
| 50-54 years | 0.54 (0.43 to 0.68) | 0.25 (0.20 to 0.32) | 0.74 (0.62 to 0.86) | 0.17 (0.14 to 0.19) | -1.40 (-1.57 to -1.23) |
| 55-59 years | 0.73 (0.58 to 0.91) | 0.39 (0.31 to 0.49) | 1.10 (0.86 to 1.32) | 0.28 (0.22 to 0.33) | -1.13 (-1.30 to -0.97) |
| 60-64 years | 0.83 (0.66 to 1.03) | 0.52 (0.41 to 0.64) | 1.15 (0.91 to 1.34) | 0.36 (0.28 to 0.42) | -1.17 (-1.31 to -1.04) |
| 65-69 years | 1.06 (0.84 to 1.32) | 0.86 (0.68 to 1.06) | 1.75 (1.38 to 2.08) | 0.63 (0.50 to 0.75) | -0.95 (-1.08 to -0.81) |
| 70-74 years | 1.16 (0.90 to 1.46) | 1.37 (1.07 to 1.72) | 2.23 (1.69 to 2.66) | 1.08 (0.82 to 1.29) | -0.75 (-0.89 to -0.61) |
| 75-79 years | 1.37 (1.07 to 1.67) | 2.22 (1.74 to 2.71) | 2.55 (1.88 to 3.07) | 1.93 (1.43 to 2.33) | -0.46 (-0.78 to -0.14) |
| 80-84 years | 1.17 (0.89 to 1.48) | 3.31 (2.53 to 4.19) | 2.79 (2.07 to 3.39) | 3.19 (2.36 to 3.87) | -0.15 (-0.38 to 0.09) |
| 85-89 years | 0.79 (0.60 to 0.99) | 5.24 (3.99 to 6.58) | 2.62 (2.03 to 3.07) | 5.72 (4.43 to 6.72) | 0.28 (0.10 to 0.47) |
| 90-94 years | 0.33 (0.25 to 0.40) | 7.66 (5.87 to 9.30) | 1.63 (1.27 to 1.92) | 9.13 (7.1 to 10.74) | 0.60 (0.40 to 0.80) |
| 95+ years | 0.10 (0.08 to 0.12) | 9.96 (7.58 to 12.06) | 0.72 (0.52 to 0.85) | 13.19 (9.51 to 15.6) | 0.91 (0.78 to 1.05) |
|  | **DALYs (95% UI)** | | | | **AAPC (95% CI)** |
|  | **DALY cases in 1990 (000s)** | **ASDR in 1990 (per 100,000)** | **DALY cases in 2021 (000s)** | **ASDR in 2021 (per 100,000)** |  |
| **Global** | 687.42 (535.24 to 813.09) | 13.21 (10.78 to 15.36) | 642.1 (552.27 to 728.99) | 8.24 (7.14 to 9.39) | -1.52 (-1.64 to -1.40) |
| **Sex** |  |  |  |  |  |
| Female | 317.73 (194.73 to 444.01) | 12.32 (7.78 to 16.8) | 342.47 (282.65 to 430.64) | 8.39 (6.92 to 10.53) | -1.26 (-1.37 to -1.15) |
| Male | 369.69 (316.03 to 424.98) | 14.09 (11.89 to 16.35) | 299.64 (246.73 to 348.33) | 8.06 (6.72 to 9.36) | -1.80 (-1.90 to -1.70) |
| **Age** |  | **DALY rate in 1990 (per 100,000)** |  | **DALY rate in 2021 (per 100,000)** |  |
| <5 years | 323.27 (182.38 to 459.66) | 52.15 (29.42 to 74.15) | 125.96 (96.51 to 159.4) | 19.14 (14.66 to 24.22) | -3.16 (-3.37 to -2.95) |
| 5-9years | 20.59 (14.83 to 27.10) | 3.53 (2.54 to 4.64) | 12.81 (10.35 to 15.87) | 1.86 (1.51 to 2.31) | -2.04 (-2.27 to -1.81) |
| 10-14 years | 14.89 (11.38 to 18.57) | 2.78 (2.12 to 3.47) | 12.33 (9.82 to 15.35) | 1.85 (1.47 to 2.3) | -1.31 (-1.54 to -1.07) |
| 15-19 years | 22.07 (17.29 to 28.51) | 4.25 (3.33 to 5.49) | 19.27 (15.49 to 24.75) | 3.09 (2.48 to 3.97) | -1.02 (-1.15 to -0.88) |
| 20-24 years | 23.29 (17.88 to 30.04) | 4.73 (3.63 to 6.11) | 22.87 (18.25 to 28.91) | 3.83 (3.06 to 4.84) | -0.71 (-0.80 to -0.61) |
| 25-29 years | 26.52 (20.74 to 33.06) | 5.99 (4.69 to 7.47) | 27.21 (22.50 to 32.56) | 4.62 (3.82 to 5.53) | -0.88 (-0.95 to -0.80) |
| 30-34 years | 20.85 (16.29 to 26.11) | 5.41 (4.23 to 6.77) | 24.18 (20.20 to 28.86) | 4.00 (3.34 to 4.77) | -0.99 (-1.16 to -0.81) |
| 35-39 years | 23.34 (18.57 to 28.83) | 6.63 (5.27 to 8.18) | 26.53 (22.78 to 30.79) | 4.73 (4.06 to 5.49) | -1.09 (-1.23 to -0.95) |
| 40-44 years | 25.03 (20.20 to 31.45) | 8.74 (7.05 to 10.98) | 30.89 (26.22 to 35.88) | 6.17 (5.24 to 7.17) | -1.13 (-1.32 to -0.94) |
| 45-49 years | 16.91 (13.56 to 20.77) | 7.28 (5.84 to 8.95) | 23.74 (20.37 to 27.85) | 5.01 (4.30 to 5.88) | -1.20 (-1.34 to -1.05) |
| 50-54 years | 21.42 (17.25 to 26.56) | 10.08 (8.12 to 12.5) | 29.66 (25.33 to 34.27) | 6.67 (5.69 to 7.70) | -1.34 (-1.52 to -1.15) |
| 55-59 years | 25.22 (20.15 to 31.04) | 13.62 (10.88 to 16.76) | 38.59 (30.61 to 46.13) | 9.75 (7.74 to 11.66) | -1.08 (-1.24 to -0.92) |
| 60-64 years | 24.87 (19.90 to 30.70) | 15.48 (12.39 to 19.11) | 34.71 (27.82 to 39.95) | 10.85 (8.69 to 12.48) | -1.13 (-1.26 to -1.01) |
| 65-69 years | 26.47 (21.14 to 32.63) | 21.41 (17.10 to 26.39) | 44.13 (34.81 to 52.09) | 16.00 (12.62 to 18.88) | -0.92 (-1.03 to -0.81) |
| 70-74 years | 23.81 (18.63 to 29.69) | 28.12 (22.01 to 35.07) | 46.04 (34.97 to 54.86) | 22.37 (16.99 to 26.65) | -0.73 (-0.86 to -0.60) |
| 75-79 years | 22.32 (17.64 to 27.22) | 36.26 (28.66 to 44.23) | 41.58 (31.08 to 49.95) | 31.53 (23.57 to 37.88) | -0.47 (-0.76 to -0.18) |
| 80-84 years | 14.91 (11.39 to 18.78) | 42.15 (32.21 to 53.10) | 35.42 (26.37 to 42.82) | 40.44 (30.11 to 48.89) | -0.13 (-0.35 to 0.08) |
| 85-89 years | 7.97 (6.10 to 10.01) | 52.73 (40.37 to 66.26) | 26.17 (20.30 to 30.62) | 57.24 (44.39 to 66.97) | 0.26 (0.08 to 0.45) |
| 90-94 years | 2.85 (2.19 to 3.46) | 66.60 (51.12 to 80.75) | 14.18 (11.02 to 16.65) | 79.27 (61.63 to 93.09) | 0.60 (0.40 to 0.79) |
| 95+ years | 0.83 (0.63 to 1.00) | 81.46 (62.20 to 98.38) | 5.82 (4.20 to 6.87) | 106.77 (77.07 to 126.03) | 0.88 (0.75 to 1.01) |

ASIR, age-standardized incidence rate; ASMR, age-standardized mortality rate; ASDR, age-standardized disability-adjusted life-year rate; AAPC, average annual percent change.

**Supplemental Table 2. Incidence of PAH between 1990 and 2021 at the global and regional levels.**

| **Location** | **Incidence (95% UI)** | | | | **AAPC (95% CI)** |
| --- | --- | --- | --- | --- | --- |
|  | **Incident cases in 1990** | **ASIR in 1990 (per 100,000)** | **Incident cases in 2021** | **ASIR in 2021 (per 100,000)** |  |
| **Global** | 23301 (19037 to 27809) | 0.50 (0.40 to 0.60) | 43251 (34705 to 52441) | 0.52 (0.42 to 0.62) | 0.10 (0.10 to 0.11) |
| **Socio-demographic index** |  |  |  |  |  |
| High SDI | 3677 (2945 to 4509) | 0.37 (0.30 to 0.45) | 5612 (4500 to 6959) | 0.37 (0.29 to 0.44) | -0.02 (-0.03 to -0.01) |
| High-middle SDI | 4469 (3599 to 5419) | 0.43 (0.35 to 0.52) | 7734 (6187 to 9586) | 0.46 (0.37 to 0.55) | 0.19 (0.17 to 0.21) |
| Middle SDI | 7264 (5922 to 8765) | 0.54 (0.43 to 0.64) | 14174 (11319 to 17383) | 0.53 (0.43 to 0.64) | -0.02 (-0.03 to -0.01) |
| Low-middle SDI | 5120 (4183 to 6102) | 0.60 (0.48 to 0.71) | 9984 (8086 to 12020) | 0.59 (0.47 to 0.70) | -0.07 (-0.10 to -0.03) |
| Low SDI | 2749 (2266 to 3274) | 0.78 (0.64 to 0.93) | 5712 (4685 to 6847) | 0.71 (0.58 to 0.85) | -0.30 (-0.33 to -0.27) |
| **Super regions** |  |  |  |  |  |
| Central Europe, Eastern Europe, and Central Asia | 1710 (1376 to 2103) | 0.38 (0.31 to 0.46) | 2392 (1928 to 2945) | 0.44 (0.36 to 0.54) | 0.48 (0.43 to 0.53) |
| High-income | 3855 (3078 to 4727) | 0.37 (0.29 to 0.44) | 5471 (4387 to 6772) | 0.35 (0.28 to 0.43) | -0.11 (-0.12 to -0.09) |
| Latin America and Caribbean | 1569 (1276 to 1893) | 0.52 (0.42 to 0.63) | 3087 (2483 to 3743) | 0.50 (0.40 to 0.60) | -0.17 (-0.20 to -0.14) |
| North Africa and Middle East | 1353 (1106 to 1612) | 0.56 (0.45 to 0.67) | 2881 (2326 to 3524) | 0.52 (0.42 to 0.63) | -0.26 (-0.33 to -0.20) |
| South Asia | 4573 (3718 to 5444) | 0.56 (0.46 to 0.67) | 9520 (7651 to 11461) | 0.56 (0.45 to 0.67) | -0.01 (-0.02 to 0.00) |
| Southeast Asia, East Asia, and Oceania | 7243 (5856 to 8757) | 0.52 (0.42 to 0.63) | 13719 (10890 to 16999) | 0.52 (0.42 to 0.63) | 0.02 (0.01 to 0.04) |
| Sub-Saharan Africa | 2998 (2485 to 3576) | 0.89 (0.72 to 1.05) | 6181 (5063 to 7442) | 0.78 (0.63 to 0.93) | -0.44 (-0.48 to -0.40) |
| **Regions** |  |  |  |  |  |
| Andean Latin America | 151 (124 to 182) | 0.53 (0.43 to 0.63) | 347 (282 to 418) | 0.55 (0.44 to 0.66) | 0.11 (0.09 to 0.13) |
| Australasia | 77 (62 to 94) | 0.35 (0.28 to 0.43) | 153 (123 to 190) | 0.37 (0.30 to 0.45) | 0.21 (0.19 to 0.22) |
| Caribbean | 137 (112 to 166) | 0.45 (0.37 to 0.55) | 247 (199 to 299) | 0.48 (0.39 to 0.58) | 0.20 (0.18 to 0.22) |
| Central Asia | 233 (190 to 279) | 0.41 (0.33 to 0.50) | 399 (320 to 487) | 0.44 (0.36 to 0.54) | 0.21 (0.16 to 0.26) |
| Central Europe | 537 (430 to 657) | 0.39 (0.31 to 0.47) | 794 (644 to 986) | 0.47 (0.38 to 0.57) | 0.60 (0.57 to 0.63) |
| Central Latin America | 679 (556 to 818) | 0.56 (0.46 to 0.68) | 1267 (1024 to 1535) | 0.49 (0.40 to 0.59) | -0.44 (-0.50 to -0.38) |
| Central Sub-Saharan Africa | 302 (247 to 363) | 0.81 (0.65 to 0.97) | 776 (636 to 938) | 0.83 (0.67 to 0.98) | 0.05 (-0.04 to 0.14) |
| East Asia | 5295 (4275 to 6415) | 0.51 (0.41 to 0.61) | 9572 (7599 to 11898) | 0.50 (0.40 to 0.60) | -0.06 (-0.08 to -0.05) |
| Eastern Europe | 941 (757 to 1162) | 0.37 (0.30 to 0.45) | 1198 (962 to 1488) | 0.43 (0.35 to 0.52) | 0.45 (0.41 to 0.50) |
| Eastern Sub-Saharan Africa | 1251 (1036 to 1497) | 0.99 (0.81 to 1.17) | 2687 (2209 to 3243) | 0.92 (0.75 to 1.09) | -0.23 (-0.25 to -0.21) |
| High-income Asia Pacific | 595 (475 to 733) | 0.31 (0.25 to 0.37) | 942 (761 to 1175) | 0.33 (0.26 to 0.40) | 0.18 (0.15 to 0.20) |
| High-income North America | 845 (683 to 1031) | 0.27 (0.22 to 0.33) | 1524 (1209 to 1891) | 0.30 (0.24 to 0.37) | 0.38 (0.36 to 0.40) |
| North Africa and Middle East | 1353 (1106 to 1612) | 0.56 (0.45 to 0.67) | 2881 (2326 to 3524) | 0.52 (0.42 to 0.63) | -0.26 (-0.33 to -0.20) |
| Oceania | 26 (21 to 31) | 0.57 (0.46 to 0.68) | 65 (53 to 79) | 0.61 (0.49 to 0.73) | 0.23 (0.18 to 0.27) |
| South Asia | 4573 (3718 to 5444) | 0.56 (0.46 to 0.67) | 9520 (7651 to 11461) | 0.56 (0.45 to 0.67) | -0.01 (-0.02 to 0.00) |
| Southeast Asia | 1922 (1567 to 2311) | 0.55 (0.44 to 0.66) | 4082 (3268 to 4966) | 0.58 (0.47 to 0.69) | 0.17 (0.15 to 0.18) |
| Southern Latin America | 162 (132 to 198) | 0.34 (0.28 to 0.41) | 259 (208 to 320) | 0.33 (0.27 to 0.41) | -0.06 (-0.08 to -0.04) |
| Southern Sub-Saharan Africa | 317 (260 to 378) | 0.81 (0.67 to 0.97) | 537 (439 to 649) | 0.75 (0.61 to 0.90) | -0.27 (-0.30 to -0.24) |
| Tropical Latin America | 602 (488 to 725) | 0.50 (0.40 to 0.60) | 1226 (984 to 1492) | 0.49 (0.39 to 0.59) | -0.07 (-0.14 to -0.00) |
| Western Europe | 2176 (1739 to 2656) | 0.46 (0.37 to 0.55) | 2593 (2092 to 3209) | 0.41 (0.33 to 0.49) | -0.34 (-0.38 to -0.30) |
| Western Sub-Saharan Africa | 1128 (931 to 1343) | 0.83 (0.68 to 0.99) | 2181 (1786 to 2634) | 0.64 (0.52 to 0.78) | -0.84 (-0.90 to -0.79) |
| **Countries and territories** |  |  |  |  |  |
| Afghanistan | 48 (39 to 58) | 0.61 (0.49 to 0.73) | 115 (93 to 139) | 0.60 (0.48 to 0.72) | -0.03 (-0.07 to 0.01) |
| Albania | 11 (9 to 14) | 0.42 (0.34 to 0.51) | 18 (14 to 22) | 0.51 (0.41 to 0.62) | 0.64 (0.61 to 0.67) |
| Algeria | 94 (77 to 111) | 0.53 (0.43 to 0.64) | 206 (165 to 249) | 0.50 (0.40 to 0.60) | -0.21 (-0.28 to -0.14) |
| American Samoa | 0 (0 to 0) | 0.53 (0.42 to 0.65) | 0 (0 to 0) | 0.56 (0.44 to 0.67) | 0.15 (0.11 to 0.19) |
| Andorra | 0 (0 to 0) | 0.44 (0.36 to 0.54) | 0 (0 to 1) | 0.41 (0.33 to 0.49) | -0.29 (-0.31 to -0.27) |
| Angola | 57 (47 to 68) | 0.83 (0.68 to 1.00) | 183 (151 to 221) | 0.86 (0.70 to 1.02) | 0.09 (0.07 to 0.12) |
| Antigua and Barbuda | 0 (0 to 0) | 0.46 (0.37 to 0.55) | 0 (0 to 1) | 0.47 (0.38 to 0.57) | 0.09 (0.07 to 0.11) |
| Argentina | 110 (89 to 134) | 0.34 (0.27 to 0.41) | 168 (135 to 206) | 0.33 (0.27 to 0.40) | -0.07 (-0.09 to -0.05) |
| Armenia | 12 (10 to 15) | 0.39 (0.32 to 0.48) | 16 (12 to 19) | 0.42 (0.34 to 0.51) | 0.20 (0.18 to 0.22) |
| Australia | 63 (51 to 77) | 0.34 (0.28 to 0.42) | 125 (100 to 156) | 0.36 (0.29 to 0.44) | 0.20 (0.19 to 0.22) |
| Austria | 41 (33 to 50) | 0.42 (0.34 to 0.51) | 50 (40 to 62) | 0.39 (0.31 to 0.47) | -0.25 (-0.31 to -0.20) |
| Azerbaijan | 27 (22 to 32) | 0.45 (0.37 to 0.54) | 42 (34 to 54) | 0.39 (0.32 to 0.47) | -0.47 (-0.54 to -0.40) |
| Bahamas | 1 (1 to 1) | 0.43 (0.35 to 0.52) | 2 (2 to 3) | 0.49 (0.40 to 0.59) | 0.48 (0.43 to 0.53) |
| Bahrain | 2 (1 to 2) | 0.46 (0.38 to 0.56) | 6 (5 to 8) | 0.46 (0.37 to 0.56) | -0.03 (-0.10 to 0.04) |
| Bangladesh | 329 (269 to 397) | 0.45 (0.36 to 0.54) | 798 (644 to 967) | 0.52 (0.42 to 0.62) | 0.46 (0.38 to 0.53) |
| Barbados | 1 (1 to 1) | 0.43 (0.35 to 0.52) | 2 (1 to 2) | 0.44 (0.36 to 0.53) | 0.10 (0.08 to 0.12) |
| Belarus | 38 (30 to 47) | 0.32 (0.26 to 0.39) | 51 (40 to 63) | 0.39 (0.32 to 0.49) | 0.65 (0.60 to 0.70) |
| Belgium | 57 (45 to 70) | 0.46 (0.37 to 0.55) | 70 (56 to 88) | 0.44 (0.35 to 0.53) | -0.14 (-0.18 to -0.11) |
| Belize | 1 (1 to 1) | 0.49 (0.40 to 0.59) | 2 (1 to 2) | 0.48 (0.38 to 0.57) | -0.07 (-0.10 to -0.05) |
| Benin | 31 (26 to 37) | 0.98 (0.80 to 1.17) | 59 (48 to 70) | 0.65 (0.53 to 0.78) | -1.30 (-1.35 to -1.25) |
| Bermuda | 0 (0 to 0) | 0.40 (0.32 to 0.50) | 0 (0 to 1) | 0.41 (0.33 to 0.51) | 0.10 (0.09 to 0.12) |
| Bhutan | 2 (2 to 3) | 0.59 (0.48 to 0.72) | 5 (4 to 6) | 0.66 (0.54 to 0.78) | 0.35 (0.33 to 0.36) |
| Bolivia (Plurinational State of) | 27 (22 to 33) | 0.59 (0.47 to 0.71) | 65 (52 to 78) | 0.61 (0.49 to 0.73) | 0.12 (0.09 to 0.14) |
| Bosnia and Herzegovina | 20 (16 to 24) | 0.45 (0.37 to 0.55) | 25 (20 to 32) | 0.52 (0.43 to 0.64) | 0.48 (0.43 to 0.54) |
| Botswana | 8 (7 to 10) | 0.90 (0.73 to 1.09) | 19 (15 to 23) | 0.90 (0.74 to 1.10) | 0.03 (0.00 to 0.05) |
| Brazil | 590 (478 to 711) | 0.50 (0.40 to 0.61) | 1198 (961 to 1459) | 0.49 (0.40 to 0.59) | -0.07 (-0.14 to -0.01) |
| Brunei Darussalam | 1 (0 to 1) | 0.32 (0.25 to 0.39) | 1 (1 to 2) | 0.33 (0.26 to 0.40) | 0.09 (0.05 to 0.12) |
| Bulgaria | 36 (29 to 45) | 0.34 (0.28 to 0.42) | 48 (38 to 60) | 0.45 (0.36 to 0.55) | 0.88 (0.81 to 0.95) |
| Burkina Faso | 62 (51 to 74) | 0.95 (0.78 to 1.12) | 93 (76 to 112) | 0.60 (0.49 to 0.72) | -1.45 (-1.58 to -1.31) |
| Burundi | 37 (30 to 44) | 0.98 (0.80 to 1.17) | 76 (62 to 92) | 0.86 (0.70 to 1.03) | -0.44 (-0.50 to -0.38) |
| Cabo Verde | 2 (1 to 2) | 0.62 (0.49 to 0.75) | 3 (2 to 3) | 0.51 (0.41 to 0.63) | -0.59 (-0.64 to -0.55) |
| Cambodia | 42 (34 to 50) | 0.61 (0.49 to 0.72) | 90 (73 to 110) | 0.60 (0.48 to 0.73) | -0.03 (-0.05 to -0.02) |
| Cameroon | 70 (57 to 83) | 0.98 (0.80 to 1.16) | 161 (131 to 197) | 0.73 (0.60 to 0.88) | -0.92 (-0.96 to -0.88) |
| Canada | 63 (50 to 78) | 0.21 (0.17 to 0.26) | 138 (109 to 171) | 0.26 (0.21 to 0.32) | 0.64 (0.59 to 0.70) |
| Central African Republic | 16 (13 to 20) | 0.88 (0.72 to 1.06) | 33 (27 to 40) | 0.86 (0.70 to 1.03) | -0.09 (-0.11 to -0.08) |
| Chad | 32 (26 to 38) | 0.77 (0.62 to 0.92) | 68 (55 to 82) | 0.61 (0.49 to 0.74) | -0.77 (-0.81 to -0.72) |
| Chile | 43 (35 to 52) | 0.36 (0.29 to 0.44) | 78 (62 to 97) | 0.35 (0.28 to 0.42) | -0.14 (-0.19 to -0.09) |
| China | 5126 (4139 to 6211) | 0.51 (0.41 to 0.62) | 9257 (7350 to 11508) | 0.50 (0.40 to 0.60) | -0.07 (-0.09 to -0.05) |
| Colombia | 126 (103 to 152) | 0.51 (0.42 to 0.62) | 244 (198 to 298) | 0.46 (0.38 to 0.56) | -0.31 (-0.35 to -0.27) |
| Comoros | 2 (2 to 3) | 0.74 (0.60 to 0.88) | 5 (4 to 6) | 0.84 (0.68 to 1.01) | 0.42 (0.32 to 0.51) |
| Congo | 13 (11 to 16) | 0.78 (0.63 to 0.95) | 30 (24 to 37) | 0.72 (0.58 to 0.86) | -0.26 (-0.35 to -0.17) |
| Cook Islands | 0 (0 to 0) | 0.50 (0.40 to 0.61) | 0 (0 to 0) | 0.54 (0.43 to 0.66) | 0.22 (0.18 to 0.26) |
| Costa Rica | 12 (10 to 14) | 0.52 (0.42 to 0.63) | 23 (19 to 29) | 0.44 (0.36 to 0.54) | -0.53 (-0.57 to -0.49) |
| Côte d'Ivoire | 82 (68 to 100) | 1.04 (0.86 to 1.24) | 139 (113 to 169) | 0.71 (0.58 to 0.86) | -1.24 (-1.38 to -1.11) |
| Croatia | 22 (17 to 27) | 0.39 (0.31 to 0.47) | 30 (24 to 37) | 0.46 (0.37 to 0.56) | 0.58 (0.56 to 0.60) |
| Cuba | 44 (35 to 53) | 0.41 (0.33 to 0.50) | 67 (54 to 84) | 0.43 (0.35 to 0.52) | 0.13 (0.12 to 0.15) |
| Cyprus | 4 (3 to 5) | 0.49 (0.39 to 0.59) | 7 (6 to 9) | 0.41 (0.33 to 0.49) | -0.58 (-0.65 to -0.51) |
| Czechia | 46 (37 to 57) | 0.38 (0.31 to 0.47) | 74 (61 to 92) | 0.46 (0.38 to 0.56) | 0.59 (0.55 to 0.63) |
| Democratic People's Republic of Korea | 90 (73 to 111) | 0.49 (0.39 to 0.59) | 159 (127 to 196) | 0.51 (0.41 to 0.61) | 0.14 (0.13 to 0.14) |
| Democratic Republic of the Congo | 207 (169 to 250) | 0.80 (0.64 to 0.96) | 511 (420 to 619) | 0.83 (0.67 to 0.98) | 0.12 (0.01 to 0.23) |
| Denmark | 34 (27 to 41) | 0.51 (0.41 to 0.63) | 33 (27 to 42) | 0.40 (0.32 to 0.49) | -0.82 (-0.85 to -0.79) |
| Djibouti | 2 (2 to 3) | 0.82 (0.67 to 0.98) | 8 (6 to 9) | 0.78 (0.64 to 0.94) | -0.13 (-0.19 to -0.06) |
| Dominica | 0 (0 to 0) | 0.51 (0.41 to 0.61) | 0 (0 to 0) | 0.47 (0.38 to 0.57) | -0.23 (-0.25 to -0.21) |
| Dominican Republic | 27 (22 to 33) | 0.51 (0.41 to 0.62) | 56 (45 to 68) | 0.53 (0.42 to 0.63) | 0.12 (0.10 to 0.13) |
| Ecuador | 39 (32 to 47) | 0.52 (0.42 to 0.62) | 89 (73 to 108) | 0.51 (0.42 to 0.63) | -0.05 (-0.07 to -0.03) |
| Egypt | 300 (245 to 361) | 0.75 (0.61 to 0.90) | 454 (366 to 559) | 0.53 (0.43 to 0.65) | -1.09 (-1.15 to -1.03) |
| El Salvador | 23 (19 to 28) | 0.59 (0.48 to 0.71) | 27 (22 to 33) | 0.44 (0.35 to 0.54) | -0.95 (-0.99 to -0.91) |
| Equatorial Guinea | 2 (2 to 3) | 0.76 (0.62 to 0.91) | 7 (6 to 9) | 0.75 (0.60 to 0.89) | -0.11 (-0.17 to -0.04) |
| Eritrea | 19 (15 to 23) | 0.88 (0.71 to 1.05) | 41 (33 to 49) | 0.87 (0.71 to 1.05) | -0.05 (-0.11 to 0.00) |
| Estonia | 6 (5 to 7) | 0.33 (0.27 to 0.40) | 7 (6 to 9) | 0.38 (0.31 to 0.47) | 0.51 (0.47 to 0.55) |
| Eswatini | 4 (3 to 5) | 0.79 (0.65 to 0.95) | 7 (6 to 8) | 0.78 (0.63 to 0.93) | -0.07 (-0.08 to -0.06) |
| Ethiopia | 343 (285 to 411) | 1.02 (0.84 to 1.21) | 755 (620 to 903) | 1.00 (0.82 to 1.19) | -0.07 (-0.10 to -0.03) |
| Fiji | 3 (3 to 4) | 0.60 (0.48 to 0.71) | 6 (5 to 7) | 0.68 (0.55 to 0.81) | 0.40 (0.32 to 0.47) |
| Finland | 31 (25 to 38) | 0.50 (0.41 to 0.61) | 34 (27 to 42) | 0.41 (0.33 to 0.50) | -0.66 (-0.75 to -0.57) |
| France | 394 (317 to 477) | 0.57 (0.46 to 0.69) | 440 (358 to 547) | 0.47 (0.38 to 0.57) | -0.60 (-0.65 to -0.55) |
| Gabon | 7 (5 to 8) | 0.88 (0.72 to 1.06) | 11 (9 to 13) | 0.75 (0.61 to 0.90) | -0.52 (-0.54 to -0.49) |
| Gambia | 4 (3 to 5) | 0.68 (0.55 to 0.81) | 11 (9 to 13) | 0.68 (0.55 to 0.82) | -0.03 (-0.14 to 0.08) |
| Georgia | 23 (19 to 29) | 0.39 (0.32 to 0.48) | 20 (16 to 25) | 0.41 (0.33 to 0.50) | 0.14 (0.11 to 0.17) |
| Germany | 500 (402 to 615) | 0.48 (0.39 to 0.58) | 476 (385 to 598) | 0.37 (0.30 to 0.45) | -0.78 (-0.90 to -0.66) |
| Ghana | 96 (78 to 114) | 0.92 (0.75 to 1.10) | 169 (138 to 206) | 0.65 (0.53 to 0.79) | -1.13 (-1.20 to -1.06) |
| Greece | 53 (42 to 66) | 0.41 (0.33 to 0.50) | 65 (53 to 80) | 0.41 (0.34 to 0.50) | 0.02 (-0.04 to 0.08) |
| Greenland | 0 (0 to 0) | 0.27 (0.22 to 0.33) | 0 (0 to 0) | 0.33 (0.26 to 0.40) | 0.60 (0.55 to 0.66) |
| Grenada | 0 (0 to 0) | 0.47 (0.38 to 0.58) | 1 (0 to 1) | 0.50 (0.40 to 0.61) | 0.22 (0.19 to 0.24) |
| Guam | 1 (0 to 1) | 0.51 (0.41 to 0.62) | 1 (1 to 1) | 0.55 (0.44 to 0.67) | 0.23 (0.19 to 0.27) |
| Guatemala | 35 (29 to 42) | 0.63 (0.51 to 0.75) | 72 (58 to 87) | 0.53 (0.43 to 0.64) | -0.56 (-0.64 to -0.48) |
| Guinea | 38 (31 to 45) | 0.85 (0.69 to 1.02) | 58 (48 to 70) | 0.63 (0.51 to 0.76) | -0.99 (-1.04 to -0.93) |
| Guinea-Bissau | 6 (5 to 7) | 0.84 (0.68 to 1.01) | 9 (7 to 11) | 0.65 (0.52 to 0.78) | -0.84 (-0.89 to -0.80) |
| Guyana | 3 (2 to 3) | 0.50 (0.40 to 0.60) | 4 (3 to 5) | 0.52 (0.42 to 0.64) | 0.15 (0.13 to 0.17) |
| Haiti | 24 (20 to 29) | 0.52 (0.42 to 0.63) | 56 (46 to 69) | 0.54 (0.44 to 0.66) | 0.14 (0.11 to 0.16) |
| Honduras | 19 (15 to 23) | 0.60 (0.49 to 0.73) | 42 (34 to 50) | 0.50 (0.40 to 0.61) | -0.57 (-0.62 to -0.52) |
| Hungary | 52 (41 to 64) | 0.41 (0.33 to 0.50) | 70 (56 to 88) | 0.48 (0.39 to 0.59) | 0.52 (0.47 to 0.56) |
| Iceland | 1 (1 to 1) | 0.40 (0.32 to 0.49) | 2 (1 to 2) | 0.37 (0.30 to 0.44) | -0.29 (-0.30 to -0.28) |
| India | 3738 (3041 to 4469) | 0.58 (0.47 to 0.69) | 7512 (6046 to 9059) | 0.56 (0.46 to 0.68) | -0.09 (-0.10 to -0.09) |
| Indonesia | 723 (588 to 872) | 0.53 (0.42 to 0.63) | 1684 (1350 to 2054) | 0.62 (0.50 to 0.74) | 0.50 (0.46 to 0.55) |
| Iran (Islamic Republic of) | 182 (148 to 218) | 0.46 (0.37 to 0.56) | 393 (317 to 482) | 0.45 (0.37 to 0.55) | -0.05 (-0.17 to 0.07) |
| Iraq | 74 (61 to 88) | 0.61 (0.49 to 0.73) | 187 (152 to 228) | 0.57 (0.46 to 0.69) | -0.24 (-0.32 to -0.16) |
| Ireland | 9 (7 to 10) | 0.23 (0.18 to 0.27) | 12 (9 to 15) | 0.18 (0.15 to 0.23) | -0.70 (-0.81 to -0.60) |
| Israel | 31 (26 to 38) | 0.65 (0.53 to 0.80) | 69 (56 to 84) | 0.64 (0.53 to 0.79) | -0.05 (-0.07 to -0.04) |
| Italy | 311 (249 to 385) | 0.43 (0.35 to 0.52) | 388 (313 to 482) | 0.42 (0.34 to 0.50) | -0.11 (-0.15 to -0.07) |
| Jamaica | 8 (7 to 10) | 0.42 (0.34 to 0.52) | 12 (10 to 15) | 0.41 (0.33 to 0.51) | -0.06 (-0.08 to -0.04) |
| Japan | 472 (374 to 584) | 0.31 (0.25 to 0.38) | 694 (561 to 866) | 0.34 (0.27 to 0.41) | 0.23 (0.20 to 0.25) |
| Jordan | 11 (9 to 14) | 0.49 (0.40 to 0.60) | 49 (40 to 60) | 0.48 (0.39 to 0.60) | -0.06 (-0.10 to -0.01) |
| Kazakhstan | 59 (48 to 72) | 0.41 (0.33 to 0.50) | 84 (67 to 104) | 0.44 (0.36 to 0.55) | 0.30 (0.22 to 0.38) |
| Kenya | 129 (106 to 156) | 0.88 (0.72 to 1.05) | 308 (251 to 369) | 0.84 (0.68 to 1.00) | -0.16 (-0.19 to -0.12) |
| Kiribati | 0 (0 to 0) | 0.61 (0.50 to 0.75) | 1 (1 to 1) | 0.66 (0.54 to 0.80) | 0.21 (0.17 to 0.25) |
| Kuwait | 5 (4 to 6) | 0.43 (0.35 to 0.53) | 19 (15 to 24) | 0.43 (0.35 to 0.52) | 0.00 (-0.05 to 0.06) |
| Kyrgyzstan | 15 (12 to 18) | 0.42 (0.34 to 0.51) | 23 (18 to 28) | 0.38 (0.31 to 0.47) | -0.27 (-0.35 to -0.19) |
| Lao People's Democratic Republic | 20 (16 to 23) | 0.66 (0.54 to 0.80) | 43 (35 to 51) | 0.69 (0.56 to 0.83) | 0.15 (0.12 to 0.17) |
| Latvia | 10 (8 to 13) | 0.33 (0.27 to 0.40) | 10 (8 to 12) | 0.35 (0.29 to 0.43) | 0.28 (0.25 to 0.31) |
| Lebanon | 12 (9 to 15) | 0.47 (0.38 to 0.58) | 27 (22 to 33) | 0.47 (0.38 to 0.56) | -0.03 (-0.09 to 0.03) |
| Lesotho | 9 (7 to 11) | 0.79 (0.64 to 0.94) | 11 (9 to 14) | 0.77 (0.62 to 0.92) | -0.06 (-0.09 to -0.04) |
| Liberia | 19 (16 to 23) | 1.11 (0.91 to 1.31) | 28 (23 to 34) | 0.73 (0.60 to 0.87) | -1.31 (-1.42 to -1.20) |
| Libya | 15 (12 to 18) | 0.52 (0.42 to 0.63) | 35 (28 to 44) | 0.54 (0.43 to 0.65) | 0.10 (0.05 to 0.15) |
| Lithuania | 13 (11 to 16) | 0.32 (0.26 to 0.40) | 15 (12 to 19) | 0.37 (0.30 to 0.45) | 0.42 (0.34 to 0.50) |
| Luxembourg | 2 (2 to 3) | 0.47 (0.38 to 0.57) | 3 (3 to 4) | 0.38 (0.31 to 0.46) | -0.65 (-0.71 to -0.59) |
| Madagascar | 70 (57 to 84) | 0.86 (0.70 to 1.04) | 172 (140 to 207) | 0.87 (0.70 to 1.03) | 0.03 (-0.02 to 0.07) |
| Malawi | 61 (51 to 73) | 0.94 (0.76 to 1.11) | 120 (99 to 146) | 0.92 (0.75 to 1.10) | -0.06 (-0.09 to -0.03) |
| Malaysia | 66 (53 to 80) | 0.51 (0.41 to 0.62) | 149 (119 to 183) | 0.48 (0.38 to 0.59) | -0.20 (-0.22 to -0.18) |
| Maldives | 1 (1 to 1) | 0.58 (0.47 to 0.71) | 2 (2 to 3) | 0.51 (0.41 to 0.62) | -0.46 (-0.49 to -0.43) |
| Mali | 53 (44 to 63) | 0.88 (0.72 to 1.05) | 96 (79 to 115) | 0.61 (0.49 to 0.73) | -1.16 (-1.21 to -1.12) |
| Malta | 2 (2 to 2) | 0.47 (0.37 to 0.57) | 3 (2 to 3) | 0.38 (0.31 to 0.47) | -0.64 (-0.70 to -0.59) |
| Marshall Islands | 0 (0 to 0) | 0.57 (0.46 to 0.70) | 0 (0 to 0) | 0.61 (0.50 to 0.74) | 0.22 (0.17 to 0.26) |
| Mauritania | 11 (9 to 13) | 0.79 (0.64 to 0.95) | 18 (15 to 21) | 0.56 (0.45 to 0.67) | -1.12 (-1.25 to -0.98) |
| Mauritius | 7 (6 to 9) | 0.78 (0.63 to 0.95) | 12 (10 to 15) | 0.73 (0.59 to 0.88) | -0.22 (-0.23 to -0.22) |
| Mexico | 363 (298 to 436) | 0.59 (0.48 to 0.71) | 661 (536 to 803) | 0.50 (0.41 to 0.60) | -0.51 (-0.59 to -0.43) |
| Micronesia (Federated States of) | 0 (0 to 0) | 0.59 (0.48 to 0.72) | 1 (0 to 1) | 0.64 (0.52 to 0.79) | 0.23 (0.19 to 0.27) |
| Monaco | 0 (0 to 0) | 0.47 (0.38 to 0.57) | 0 (0 to 0) | 0.41 (0.33 to 0.50) | -0.39 (-0.42 to -0.36) |
| Mongolia | 6 (5 to 7) | 0.38 (0.31 to 0.47) | 14 (11 to 17) | 0.48 (0.38 to 0.58) | 0.71 (0.68 to 0.75) |
| Montenegro | 2 (2 to 3) | 0.36 (0.29 to 0.43) | 3 (3 to 4) | 0.40 (0.33 to 0.49) | 0.39 (0.34 to 0.44) |
| Morocco | 100 (81 to 120) | 0.53 (0.42 to 0.65) | 191 (153 to 234) | 0.52 (0.42 to 0.63) | -0.04 (-0.09 to 0.01) |
| Mozambique | 84 (69 to 100) | 0.89 (0.73 to 1.07) | 184 (152 to 222) | 0.91 (0.74 to 1.08) | 0.04 (-0.00 to 0.08) |
| Myanmar | 187 (152 to 225) | 0.60 (0.48 to 0.73) | 321 (259 to 391) | 0.60 (0.49 to 0.72) | -0.04 (-0.05 to -0.02) |
| Namibia | 9 (7 to 11) | 0.92 (0.75 to 1.10) | 17 (14 to 20) | 0.86 (0.70 to 1.02) | -0.25 (-0.32 to -0.19) |
| Nauru | 0 (0 to 0) | 0.61 (0.49 to 0.74) | 0 (0 to 0) | 0.66 (0.53 to 0.81) | 0.24 (0.19 to 0.28) |
| Nepal | 72 (59 to 86) | 0.52 (0.43 to 0.63) | 155 (125 to 187) | 0.57 (0.46 to 0.69) | 0.26 (0.23 to 0.29) |
| Netherlands | 95 (77 to 114) | 0.55 (0.44 to 0.65) | 128 (102 to 159) | 0.51 (0.42 to 0.62) | -0.20 (-0.23 to -0.17) |
| New Zealand | 14 (11 to 17) | 0.39 (0.31 to 0.47) | 28 (22 to 35) | 0.42 (0.34 to 0.51) | 0.23 (0.21 to 0.24) |
| Nicaragua | 15 (13 to 18) | 0.61 (0.50 to 0.73) | 30 (25 to 36) | 0.51 (0.41 to 0.62) | -0.56 (-0.62 to -0.50) |
| Niger | 39 (32 to 47) | 0.77 (0.62 to 0.92) | 92 (76 to 110) | 0.59 (0.47 to 0.71) | -0.85 (-0.89 to -0.82) |
| Nigeria | 475 (391 to 565) | 0.73 (0.60 to 0.88) | 1020 (837 to 1241) | 0.63 (0.51 to 0.77) | -0.48 (-0.57 to -0.39) |
| Niue | 0 (0 to 0) | 0.53 (0.43 to 0.65) | 0 (0 to 0) | 0.57 (0.46 to 0.69) | 0.22 (0.18 to 0.26) |
| North Macedonia | 8 (6 to 10) | 0.40 (0.33 to 0.49) | 14 (11 to 17) | 0.48 (0.39 to 0.58) | 0.57 (0.53 to 0.60) |
| Northern Mariana Islands | 0 (0 to 0) | 0.50 (0.40 to 0.60) | 0 (0 to 0) | 0.54 (0.43 to 0.66) | 0.24 (0.19 to 0.28) |
| Norway | 24 (19 to 29) | 0.46 (0.37 to 0.55) | 33 (26 to 40) | 0.45 (0.36 to 0.54) | -0.06 (-0.08 to -0.05) |
| Oman | 7 (5 to 8) | 0.53 (0.43 to 0.64) | 20 (16 to 25) | 0.54 (0.43 to 0.66) | 0.04 (-0.03 to 0.10) |
| Pakistan | 431 (352 to 510) | 0.54 (0.44 to 0.66) | 1051 (848 to 1261) | 0.59 (0.48 to 0.71) | 0.25 (0.19 to 0.32) |
| Palau | 0 (0 to 0) | 0.55 (0.44 to 0.67) | 0 (0 to 0) | 0.59 (0.47 to 0.71) | 0.19 (0.15 to 0.24) |
| Palestine | 6 (5 to 7) | 0.45 (0.37 to 0.55) | 19 (15 to 22) | 0.50 (0.40 to 0.61) | 0.30 (0.19 to 0.41) |
| Panama | 10 (8 to 12) | 0.52 (0.43 to 0.64) | 19 (16 to 24) | 0.44 (0.36 to 0.54) | -0.55 (-0.59 to -0.51) |
| Papua New Guinea | 16 (13 to 19) | 0.57 (0.46 to 0.69) | 47 (38 to 57) | 0.60 (0.49 to 0.72) | 0.19 (0.15 to 0.23) |
| Paraguay | 12 (10 to 15) | 0.41 (0.33 to 0.51) | 28 (23 to 34) | 0.43 (0.35 to 0.52) | 0.10 (0.02 to 0.18) |
| Peru | 85 (69 to 103) | 0.51 (0.42 to 0.62) | 193 (157 to 233) | 0.54 (0.44 to 0.66) | 0.18 (0.16 to 0.21) |
| Philippines | 284 (232 to 339) | 0.63 (0.51 to 0.76) | 639 (518 to 772) | 0.64 (0.52 to 0.77) | 0.05 (0.03 to 0.06) |
| Poland | 174 (140 to 211) | 0.42 (0.34 to 0.51) | 265 (214 to 327) | 0.48 (0.39 to 0.58) | 0.41 (0.33 to 0.49) |
| Portugal | 35 (28 to 44) | 0.29 (0.23 to 0.36) | 47 (37 to 58) | 0.28 (0.23 to 0.34) | -0.13 (-0.21 to -0.05) |
| Puerto Rico | 15 (12 to 18) | 0.41 (0.33 to 0.50) | 21 (17 to 27) | 0.43 (0.34 to 0.52) | 0.12 (0.10 to 0.14) |
| Qatar | 1 (1 to 2) | 0.45 (0.37 to 0.55) | 11 (8 to 14) | 0.45 (0.36 to 0.55) | -0.03 (-0.09 to 0.02) |
| Republic of Korea | 114 (93 to 139) | 0.29 (0.24 to 0.35) | 220 (173 to 280) | 0.30 (0.24 to 0.37) | 0.07 (0.05 to 0.09) |
| Republic of Moldova | 16 (13 to 20) | 0.36 (0.29 to 0.43) | 21 (17 to 26) | 0.43 (0.35 to 0.52) | 0.62 (0.55 to 0.70) |
| Romania | 94 (76 to 117) | 0.37 (0.30 to 0.44) | 126 (102 to 159) | 0.45 (0.36 to 0.55) | 0.65 (0.60 to 0.70) |
| Russian Federation | 622 (498 to 770) | 0.37 (0.30 to 0.45) | 804 (647 to 999) | 0.42 (0.34 to 0.50) | 0.34 (0.27 to 0.42) |
| Rwanda | 46 (38 to 56) | 0.98 (0.80 to 1.16) | 92 (76 to 110) | 0.94 (0.77 to 1.12) | -0.12 (-0.19 to -0.06) |
| Saint Kitts and Nevis | 0 (0 to 0) | 0.47 (0.38 to 0.57) | 0 (0 to 0) | 0.52 (0.42 to 0.63) | 0.37 (0.34 to 0.40) |
| Saint Lucia | 1 (0 to 1) | 0.48 (0.39 to 0.58) | 1 (1 to 1) | 0.46 (0.37 to 0.56) | -0.13 (-0.16 to -0.10) |
| Saint Vincent and the Grenadines | 0 (0 to 1) | 0.50 (0.40 to 0.60) | 1 (1 to 1) | 0.48 (0.39 to 0.58) | -0.14 (-0.19 to -0.08) |
| Samoa | 1 (0 to 1) | 0.48 (0.39 to 0.58) | 1 (1 to 1) | 0.53 (0.43 to 0.65) | 0.31 (0.23 to 0.40) |
| San Marino | 0 (0 to 0) | 0.44 (0.36 to 0.54) | 0 (0 to 0) | 0.40 (0.32 to 0.49) | -0.33 (-0.34 to -0.32) |
| Sao Tome and Principe | 1 (0 to 1) | 0.63 (0.51 to 0.76) | 1 (1 to 1) | 0.53 (0.43 to 0.65) | -0.53 (-0.58 to -0.47) |
| Saudi Arabia | 55 (45 to 67) | 0.54 (0.43 to 0.66) | 156 (123 to 198) | 0.47 (0.38 to 0.58) | -0.43 (-0.52 to -0.33) |
| Senegal | 59 (50 to 71) | 1.15 (0.95 to 1.37) | 80 (66 to 96) | 0.69 (0.56 to 0.83) | -1.61 (-1.70 to -1.52) |
| Serbia | 35 (28 to 44) | 0.33 (0.26 to 0.40) | 57 (46 to 72) | 0.45 (0.36 to 0.54) | 0.97 (0.91 to 1.03) |
| Seychelles | 0 (0 to 0) | 0.52 (0.41 to 0.63) | 1 (0 to 1) | 0.50 (0.41 to 0.61) | -0.12 (-0.13 to -0.11) |
| Sierra Leone | 27 (22 to 32) | 0.90 (0.74 to 1.08) | 39 (32 to 48) | 0.62 (0.51 to 0.75) | -1.18 (-1.23 to -1.14) |
| Singapore | 9 (7 to 11) | 0.31 (0.25 to 0.39) | 26 (21 to 33) | 0.35 (0.28 to 0.43) | 0.34 (0.32 to 0.36) |
| Slovakia | 20 (16 to 24) | 0.35 (0.28 to 0.43) | 37 (30 to 47) | 0.49 (0.40 to 0.59) | 1.07 (0.95 to 1.19) |
| Slovenia | 8 (7 to 10) | 0.37 (0.30 to 0.45) | 14 (11 to 18) | 0.44 (0.36 to 0.54) | 0.57 (0.52 to 0.62) |
| Solomon Islands | 1 (1 to 2) | 0.59 (0.48 to 0.72) | 3 (3 to 4) | 0.64 (0.52 to 0.78) | 0.22 (0.16 to 0.28) |
| Somalia | 44 (36 to 53) | 0.88 (0.72 to 1.05) | 115 (95 to 141) | 0.88 (0.72 to 1.06) | -0.02 (-0.07 to 0.03) |
| South Africa | 227 (187 to 272) | 0.79 (0.65 to 0.95) | 398 (324 to 482) | 0.74 (0.60 to 0.88) | -0.25 (-0.28 to -0.21) |
| South Sudan | 33 (28 to 40) | 0.83 (0.68 to 1.00) | 58 (47 to 70) | 0.88 (0.71 to 1.06) | 0.17 (0.14 to 0.21) |
| Spain | 181 (145 to 223) | 0.39 (0.32 to 0.47) | 251 (202 to 314) | 0.37 (0.30 to 0.45) | -0.13 (-0.18 to -0.09) |
| Sri Lanka | 84 (69 to 101) | 0.61 (0.49 to 0.74) | 148 (118 to 184) | 0.59 (0.47 to 0.71) | -0.11 (-0.12 to -0.10) |
| Sudan | 78 (65 to 94) | 0.57 (0.46 to 0.69) | 167 (137 to 201) | 0.54 (0.44 to 0.65) | -0.17 (-0.22 to -0.12) |
| Suriname | 2 (1 to 2) | 0.51 (0.41 to 0.62) | 3 (3 to 4) | 0.53 (0.43 to 0.64) | 0.09 (0.07 to 0.12) |
| Sweden | 86 (70 to 104) | 0.77 (0.63 to 0.92) | 114 (93 to 141) | 0.78 (0.64 to 0.94) | 0.04 (0.01 to 0.06) |
| Switzerland | 67 (55 to 81) | 0.79 (0.65 to 0.96) | 95 (77 to 117) | 0.75 (0.62 to 0.90) | -0.17 (-0.22 to -0.12) |
| Syrian Arab Republic | 50 (41 to 59) | 0.61 (0.49 to 0.74) | 86 (69 to 106) | 0.61 (0.50 to 0.74) | -0.03 (-0.14 to 0.08) |
| Taiwan (Province of China) | 79 (63 to 96) | 0.43 (0.34 to 0.52) | 156 (124 to 195) | 0.45 (0.36 to 0.55) | 0.17 (0.16 to 0.18) |
| Tajikistan | 16 (13 to 19) | 0.42 (0.34 to 0.51) | 35 (29 to 43) | 0.44 (0.36 to 0.53) | 0.14 (0.10 to 0.19) |
| Thailand | 252 (204 to 304) | 0.54 (0.44 to 0.65) | 478 (379 to 599) | 0.53 (0.42 to 0.63) | -0.07 (-0.10 to -0.05) |
| Timor-Leste | 3 (2 to 4) | 0.59 (0.48 to 0.72) | 6 (5 to 7) | 0.57 (0.46 to 0.69) | -0.11 (-0.12 to -0.10) |
| Togo | 21 (17 to 25) | 0.90 (0.73 to 1.06) | 39 (32 to 47) | 0.63 (0.51 to 0.76) | -1.11 (-1.19 to -1.04) |
| Tokelau | 0 (0 to 0) | 0.59 (0.48 to 0.72) | 0 (0 to 0) | 0.57 (0.46 to 0.70) | -0.14 (-0.25 to -0.03) |
| Tonga | 0 (0 to 0) | 0.51 (0.41 to 0.62) | 0 (0 to 1) | 0.54 (0.43 to 0.65) | 0.18 (0.14 to 0.23) |
| Trinidad and Tobago | 5 (4 to 6) | 0.49 (0.40 to 0.59) | 8 (6 to 10) | 0.46 (0.37 to 0.56) | -0.21 (-0.26 to -0.17) |
| Tunisia | 31 (25 to 38) | 0.49 (0.39 to 0.59) | 61 (48 to 75) | 0.46 (0.37 to 0.56) | -0.15 (-0.20 to -0.10) |
| Türkiye | 213 (173 to 254) | 0.48 (0.38 to 0.58) | 478 (381 to 588) | 0.52 (0.42 to 0.63) | 0.28 (0.14 to 0.41) |
| Turkmenistan | 12 (10 to 14) | 0.44 (0.36 to 0.54) | 21 (17 to 26) | 0.46 (0.38 to 0.56) | 0.12 (0.11 to 0.14) |
| Tuvalu | 0 (0 to 0) | 0.58 (0.47 to 0.71) | 0 (0 to 0) | 0.63 (0.51 to 0.77) | 0.27 (0.22 to 0.31) |
| Uganda | 156 (129 to 187) | 1.38 (1.13 to 1.66) | 278 (230 to 338) | 1.00 (0.82 to 1.20) | -1.05 (-1.28 to -0.83) |
| Ukraine | 236 (190 to 296) | 0.38 (0.31 to 0.46) | 290 (234 to 362) | 0.47 (0.38 to 0.58) | 0.71 (0.67 to 0.76) |
| United Arab Emirates | 6 (5 to 7) | 0.47 (0.38 to 0.57) | 46 (34 to 61) | 0.48 (0.38 to 0.58) | 0.01 (-0.05 to 0.07) |
| United Kingdom | 217 (174 to 266) | 0.30 (0.24 to 0.37) | 272 (217 to 336) | 0.29 (0.23 to 0.35) | -0.15 (-0.19 to -0.12) |
| United Republic of Tanzania | 174 (144 to 207) | 1.00 (0.82 to 1.19) | 334 (275 to 399) | 0.81 (0.66 to 0.98) | -0.66 (-0.69 to -0.63) |
| United States of America | 782 (632 to 954) | 0.28 (0.22 to 0.34) | 1386 (1099 to 1723) | 0.31 (0.25 to 0.37) | 0.35 (0.34 to 0.37) |
| United States Virgin Islands | 0 (0 to 1) | 0.43 (0.35 to 0.53) | 1 (0 to 1) | 0.45 (0.36 to 0.55) | 0.10 (0.08 to 0.12) |
| Uruguay | 10 (8 to 12) | 0.28 (0.22 to 0.34) | 13 (11 to 16) | 0.31 (0.25 to 0.37) | 0.35 (0.31 to 0.39) |
| Uzbekistan | 63 (51 to 76) | 0.41 (0.33 to 0.50) | 145 (116 to 177) | 0.47 (0.38 to 0.57) | 0.45 (0.37 to 0.52) |
| Vanuatu | 1 (0 to 1) | 0.54 (0.43 to 0.66) | 1 (1 to 2) | 0.58 (0.47 to 0.70) | 0.21 (0.16 to 0.25) |
| Venezuela (Bolivarian Republic of) | 74 (60 to 90) | 0.53 (0.43 to 0.64) | 149 (120 to 183) | 0.51 (0.42 to 0.62) | -0.14 (-0.23 to -0.06) |
| Viet Nam | 251 (206 to 304) | 0.50 (0.40 to 0.60) | 504 (403 to 631) | 0.48 (0.39 to 0.60) | -0.08 (-0.10 to -0.06) |
| Yemen | 64 (53 to 77) | 0.75 (0.62 to 0.90) | 154 (126 to 187) | 0.66 (0.54 to 0.79) | -0.45 (-0.54 to -0.35) |
| Zambia | 50 (41 to 60) | 0.97 (0.79 to 1.17) | 139 (115 to 168) | 1.06 (0.88 to 1.28) | 0.29 (0.23 to 0.35) |
| Zimbabwe | 60 (49 to 72) | 0.89 (0.73 to 1.07) | 86 (70 to 103) | 0.77 (0.62 to 0.93) | -0.48 (-0.53 to -0.42) |

**Supplemental Table 3. Morality of PAH between 1990 and 2021 at the global and regional levels.**

| **Location** | **Deaths (95% UI)** | | | | **AAPC (95% CI)** |
| --- | --- | --- | --- | --- | --- |
|  | **Death cases in 1990** | **ASMR in 1990 (per 100,000)** | **Death cases in 2021** | **ASMR in 2021 (per 100,000)** |  |
| **Global** | 14842 (12370 to 17485) | 0.35 (0.29 to 0.42) | 22021 (18239 to 25352) | 0.27 (0.23 to 0.32) | -0.82 (-0.95 to -0.68) |
| **Socio-demographic index** |  |  |  |  |  |
| High SDI | 2617 (2379 to 2850) | 0.26 (0.24 to 0.28) | 4621 (3919 to 5054) | 0.22 (0.19 to 0.23) | -0.59 (-0.75 to -0.44) |
| High-middle SDI | 3214 (2772 to 3908) | 0.35 (0.31 to 0.43) | 4326 (3594 to 5141) | 0.24 (0.20 to 0.29) | -1.22 (-1.50 to -0.95) |
| Middle SDI | 4729 (3774 to 5852) | 0.45 (0.35 to 0.58) | 7548 (5141 to 9026) | 0.33 (0.22 to 0.39) | -1.06 (-1.19 to -0.93) |
| Low-middle SDI | 3125 (2220 to 3904) | 0.35 (0.22 to 0.47) | 3728 (2757 to 5091) | 0.26 (0.18 to 0.38) | -0.89 (-1.03 to -0.75) |
| Low SDI | 1145 (740 to 1755) | 0.32 (0.15 to 0.51) | 1782 (1147 to 2544) | 0.27 (0.15 to 0.40) | -0.53 (-0.81 to -0.25) |
| **Super regions** |  |  |  |  |  |
| Central Europe, Eastern Europe, and Central Asia | 1126 (1019 to 1265) | 0.26 (0.24 to 0.29) | 1034 (959 to 1122) | 0.18 (0.17 to 0.20) | -1.26 (-1.64 to -0.87) |
| High-income | 2946 (2681 to 3207) | 0.28 (0.25 to 0.30) | 4924 (4187 to 5350) | 0.22 (0.20 to 0.24) | -0.72 (-0.88 to -0.56) |
| Latin America and Caribbean | 782 (687 to 882) | 0.28 (0.25 to 0.30) | 1167 (1063 to 1266) | 0.20 (0.18 to 0.21) | -1.13 (-1.29 to -0.97) |
| North Africa and Middle East | 2142 (1309 to 2739) | 0.77 (0.56 to 1.00) | 1896 (1328 to 2305) | 0.44 (0.31 to 0.53) | -1.75 (-1.90 to -1.61) |
| South Asia | 2385 (1502 to 3418) | 0.31 (0.17 to 0.50) | 3549 (2321 to 5532) | 0.25 (0.16 to 0.42) | -0.66 (-1.08 to -0.25) |
| Southeast Asia, East Asia, and Oceania | 4632 (3533 to 6458) | 0.47 (0.35 to 0.66) | 8256 (5870 to 10248) | 0.35 (0.26 to 0.44) | -0.97 (-1.19 to -0.76) |
| Sub-Saharan Africa | 830 (514 to 1546) | 0.24 (0.11 to 0.47) | 1194 (649 to 1940) | 0.17 (0.08 to 0.28) | -1.17 (-1.20 to -1.13) |
| **Regions** |  |  |  |  |  |
| Andean Latin America | 84 (56 to 112) | 0.28 (0.21 to 0.35) | 91 (72 to 119) | 0.16 (0.12 to 0.20) | -1.83 (-2.14 to -1.51) |
| Australasia | 45 (39 to 57) | 0.21 (0.18 to 0.26) | 58 (49 to 65) | 0.11 (0.10 to 0.13) | -1.85 (-2.22 to -1.49) |
| Caribbean | 124 (83 to 169) | 0.38 (0.28 to 0.49) | 95 (66 to 130) | 0.20 (0.13 to 0.29) | -1.96 (-2.19 to -1.73) |
| Central Asia | 208 (163 to 240) | 0.40 (0.31 to 0.47) | 319 (261 to 382) | 0.41 (0.34 to 0.48) | 0.09 (-0.22 to 0.41) |
| Central Europe | 355 (309 to 394) | 0.26 (0.22 to 0.28) | 438 (398 to 479) | 0.21 (0.19 to 0.23) | -0.70 (-0.96 to -0.43) |
| Central Latin America | 180 (157 to 210) | 0.16 (0.14 to 0.19) | 201 (177 to 230) | 0.08 (0.07 to 0.10) | -2.13 (-2.36 to -1.90) |
| Central Sub-Saharan Africa | 87 (55 to 163) | 0.24 (0.11 to 0.47) | 131 (62 to 237) | 0.19 (0.08 to 0.37) | -0.76 (-0.80 to -0.73) |
| East Asia | 4115 (3141 to 5526) | 0.59 (0.45 to 0.81) | 7490 (4986 to 9266) | 0.41 (0.28 to 0.50) | -1.23 (-1.49 to -0.96) |
| Eastern Europe | 563 (512 to 651) | 0.24 (0.22 to 0.27) | 278 (258 to 300) | 0.09 (0.08 to 0.10) | -2.98 (-3.74 to -2.21) |
| Eastern Sub-Saharan Africa | 366 (217 to 687) | 0.27 (0.12 to 0.52) | 468 (219 to 878) | 0.18 (0.07 to 0.34) | -1.37 (-1.44 to -1.29) |
| High-income Asia Pacific | 434 (410 to 459) | 0.26 (0.24 to 0.27) | 1049 (826 to 1201) | 0.23 (0.20 to 0.26) | -0.34 (-0.61 to -0.06) |
| High-income North America | 1064 (947 to 1167) | 0.32 (0.28 to 0.35) | 1880 (1620 to 2043) | 0.29 (0.26 to 0.31) | -0.26 (-0.43 to -0.09) |
| North Africa and Middle East | 2142 (1309 to 2739) | 0.77 (0.56 to 1.00) | 1896 (1328 to 2305) | 0.44 (0.31 to 0.53) | -1.75 (-1.90 to -1.61) |
| Oceania | 12 (8 to 19) | 0.28 (0.18 to 0.53) | 25 (17 to 43) | 0.24 (0.16 to 0.48) | -0.47 (-0.55 to -0.38) |
| South Asia | 2385 (1502 to 3418) | 0.31 (0.17 to 0.50) | 3549 (2321 to 5532) | 0.25 (0.16 to 0.42) | -0.66 (-1.08 to -0.25) |
| Southeast Asia | 506 (340 to 1094) | 0.15 (0.09 to 0.43) | 741 (525 to 1850) | 0.12 (0.08 to 0.32) | -0.76 (-0.79 to -0.72) |
| Southern Latin America | 169 (151 to 186) | 0.36 (0.32 to 0.40) | 150 (138 to 162) | 0.18 (0.17 to 0.20) | -2.17 (-2.47 to -1.87) |
| Southern Sub-Saharan Africa | 43 (32 to 58) | 0.12 (0.08 to 0.17) | 72 (53 to 86) | 0.11 (0.08 to 0.13) | -0.26 (-0.40 to -0.12) |
| Tropical Latin America | 394 (373 to 412) | 0.37 (0.35 to 0.39) | 779 (714 to 822) | 0.32 (0.29 to 0.34) | -0.50 (-0.79 to -0.20) |
| Western Europe | 1233 (1094 to 1380) | 0.24 (0.21 to 0.27) | 1788 (1533 to 1943) | 0.18 (0.16 to 0.19) | -0.94 (-1.19 to -0.68) |
| Western Sub-Saharan Africa | 335 (195 to 616) | 0.25 (0.09 to 0.51) | 523 (306 to 774) | 0.17 (0.07 to 0.28) | -1.29 (-1.35 to -1.24) |
| **Countries and territories** |  |  |  |  |  |
| Afghanistan | 38 (19 to 61) | 0.41 (0.21 to 0.73) | 125 (56 to 189) | 0.67 (0.24 to 1.10) | 1.61 (1.51 to 1.71) |
| Albania | 11 (8 to 15) | 0.52 (0.36 to 0.70) | 11 (7 to 19) | 0.28 (0.19 to 0.48) | -2.04 (-2.42 to -1.66) |
| Algeria | 52 (34 to 94) | 0.31 (0.19 to 0.67) | 119 (43 to 185) | 0.40 (0.13 to 0.61) | 0.89 (0.31 to 1.47) |
| American Samoa | 0 (0 to 0) | 0.17 (0.12 to 0.39) | 0 (0 to 0) | 0.13 (0.07 to 0.28) | -0.85 (-0.89 to -0.82) |
| Andorra | 0 (0 to 0) | 0.25 (0.18 to 0.35) | 0 (0 to 0) | 0.11 (0.07 to 0.17) | -2.57 (-2.82 to -2.33) |
| Angola | 18 (11 to 39) | 0.27 (0.13 to 0.56) | 30 (16 to 57) | 0.19 (0.09 to 0.36) | -1.11 (-1.36 to -0.86) |
| Antigua and Barbuda | 0 (0 to 0) | 0.11 (0.10 to 0.12) | 0 (0 to 0) | 0.04 (0.04 to 0.04) | -3.20 (-4.02 to -2.38) |
| Argentina | 137 (120 to 153) | 0.43 (0.38 to 0.48) | 108 (100 to 118) | 0.20 (0.19 to 0.22) | -2.48 (-2.71 to -2.26) |
| Armenia | 7 (6 to 8) | 0.25 (0.21 to 0.31) | 3 (3 to 4) | 0.08 (0.06 to 0.09) | -3.83 (-4.38 to -3.28) |
| Australia | 40 (34 to 52) | 0.22 (0.19 to 0.29) | 50 (42 to 57) | 0.12 (0.10 to 0.13) | -2.01 (-2.45 to -1.57) |
| Austria | 20 (18 to 21) | 0.18 (0.16 to 0.19) | 25 (22 to 28) | 0.13 (0.12 to 0.14) | -1.00 (-1.28 to -0.72) |
| Azerbaijan | 37 (25 to 52) | 0.70 (0.48 to 0.99) | 54 (32 to 83) | 0.57 (0.36 to 0.85) | -0.69 (-0.99 to -0.39) |
| Bahamas | 2 (2 to 2) | 1.06 (0.93 to 1.18) | 2 (1 to 2) | 0.40 (0.32 to 0.50) | -3.02 (-3.41 to -2.64) |
| Bahrain | 0 (0 to 1) | 0.19 (0.14 to 0.51) | 1 (1 to 2) | 0.16 (0.07 to 0.31) | -0.72 (-1.54 to 0.10) |
| Bangladesh | 263 (124 to 437) | 0.43 (0.16 to 0.89) | 383 (189 to 664) | 0.30 (0.14 to 0.57) | -0.89 (-1.16 to -0.62) |
| Barbados | 3 (3 to 3) | 1.06 (0.91 to 1.19) | 2 (1 to 2) | 0.36 (0.29 to 0.45) | -3.16 (-3.56 to -2.76) |
| Belarus | 5 (4 to 6) | 0.04 (0.03 to 0.05) | 6 (5 to 7) | 0.04 (0.03 to 0.05) | -0.35 (-0.54 to -0.16) |
| Belgium | 44 (38 to 56) | 0.31 (0.27 to 0.39) | 45 (38 to 50) | 0.18 (0.16 to 0.20) | -1.79 (-1.91 to -1.66) |
| Belize | 1 (1 to 1) | 0.46 (0.42 to 0.52) | 0 (0 to 1) | 0.15 (0.13 to 0.16) | -3.68 (-4.24 to -3.13) |
| Benin | 7 (4 to 13) | 0.21 (0.08 to 0.43) | 13 (7 to 20) | 0.15 (0.06 to 0.27) | -1.05 (-1.18 to -0.91) |
| Bermuda | 1 (1 to 1) | 1.73 (1.49 to 2.01) | 1 (1 to 1) | 0.57 (0.48 to 0.69) | -3.59 (-3.87 to -3.31) |
| Bhutan | 1 (1 to 2) | 0.39 (0.17 to 0.67) | 2 (1 to 3) | 0.31 (0.15 to 0.52) | -0.75 (-0.95 to -0.55) |
| Bolivia (Plurinational State of) | 23 (12 to 37) | 0.42 (0.26 to 0.62) | 21 (15 to 30) | 0.24 (0.17 to 0.34) | -1.81 (-1.87 to -1.75) |
| Bosnia and Herzegovina | 10 (7 to 14) | 0.27 (0.19 to 0.37) | 12 (9 to 16) | 0.21 (0.15 to 0.27) | -0.78 (-1.09 to -0.47) |
| Botswana | 1 (1 to 2) | 0.18 (0.11 to 0.28) | 2 (1 to 2) | 0.10 (0.07 to 0.14) | -1.73 (-2.09 to -1.37) |
| Brazil | 392 (371 to 410) | 0.38 (0.36 to 0.40) | 775 (710 to 818) | 0.33 (0.30 to 0.34) | -0.51 (-0.80 to -0.21) |
| Brunei Darussalam | 1 (0 to 1) | 0.48 (0.29 to 0.65) | 1 (1 to 1) | 0.27 (0.20 to 0.34) | -1.83 (-2.00 to -1.67) |
| Bulgaria | 16 (13 to 18) | 0.16 (0.13 to 0.18) | 11 (8 to 15) | 0.09 (0.07 to 0.12) | -1.60 (-2.52 to -0.68) |
| Burkina Faso | 20 (9 to 38) | 0.32 (0.09 to 0.67) | 36 (17 to 59) | 0.27 (0.09 to 0.52) | -0.57 (-0.71 to -0.43) |
| Burundi | 16 (9 to 31) | 0.37 (0.15 to 0.83) | 16 (6 to 32) | 0.21 (0.07 to 0.44) | -1.84 (-2.08 to -1.61) |
| Cabo Verde | 1 (0 to 1) | 0.21 (0.06 to 0.48) | 1 (0 to 1) | 0.13 (0.05 to 0.24) | -1.53 (-1.81 to -1.26) |
| Cambodia | 12 (6 to 26) | 0.16 (0.08 to 0.43) | 17 (10 to 38) | 0.13 (0.07 to 0.35) | -0.74 (-0.82 to -0.66) |
| Cameroon | 15 (8 to 28) | 0.24 (0.09 to 0.48) | 32 (19 to 53) | 0.17 (0.08 to 0.31) | -1.16 (-1.33 to -0.99) |
| Canada | 93 (85 to 100) | 0.31 (0.29 to 0.33) | 95 (83 to 104) | 0.14 (0.13 to 0.15) | -2.30 (-2.82 to -1.78) |
| Central African Republic | 5 (3 to 11) | 0.30 (0.14 to 0.65) | 8 (4 to 14) | 0.25 (0.10 to 0.45) | -0.69 (-0.77 to -0.61) |
| Chad | 11 (5 to 24) | 0.26 (0.07 to 0.64) | 27 (15 to 49) | 0.24 (0.08 to 0.47) | -0.13 (-0.31 to 0.05) |
| Chile | 24 (23 to 26) | 0.22 (0.20 to 0.23) | 34 (30 to 37) | 0.14 (0.13 to 0.16) | -1.33 (-1.59 to -1.07) |
| China | 4059 (3099 to 5452) | 0.61 (0.46 to 0.83) | 7318 (4836 to 9076) | 0.42 (0.28 to 0.51) | -1.26 (-1.53 to -0.99) |
| Colombia | 34 (28 to 43) | 0.15 (0.12 to 0.19) | 52 (43 to 63) | 0.10 (0.08 to 0.12) | -1.44 (-1.75 to -1.13) |
| Comoros | 1 (1 to 2) | 0.27 (0.12 to 0.50) | 1 (0 to 2) | 0.15 (0.07 to 0.30) | -1.95 (-2.72 to -1.17) |
| Congo | 3 (2 to 7) | 0.22 (0.13 to 0.45) | 4 (3 to 8) | 0.15 (0.08 to 0.29) | -1.28 (-1.45 to -1.11) |
| Cook Islands | 0 (0 to 0) | 0.26 (0.19 to 0.54) | 0 (0 to 0) | 0.14 (0.08 to 0.33) | -2.03 (-2.17 to -1.89) |
| Costa Rica | 8 (8 to 9) | 0.43 (0.39 to 0.48) | 6 (5 to 6) | 0.10 (0.09 to 0.12) | -4.59 (-5.15 to -4.02) |
| Côte d'Ivoire | 17 (10 to 30) | 0.24 (0.09 to 0.47) | 31 (16 to 50) | 0.18 (0.07 to 0.32) | -0.99 (-1.24 to -0.74) |
| Croatia | 6 (6 to 7) | 0.12 (0.10 to 0.14) | 4 (3 to 5) | 0.05 (0.04 to 0.05) | -3.09 (-4.00 to -2.16) |
| Cuba | 11 (10 to 12) | 0.11 (0.10 to 0.12) | 9 (8 to 10) | 0.05 (0.04 to 0.06) | -2.14 (-2.44 to -1.84) |
| Cyprus | 13 (7 to 17) | 2.09 (1.09 to 2.75) | 15 (7 to 19) | 0.80 (0.38 to 1.02) | -3.05 (-3.50 to -2.60) |
| Czechia | 51 (43 to 60) | 0.39 (0.33 to 0.46) | 88 (76 to 101) | 0.43 (0.36 to 0.49) | 0.31 (-0.31 to 0.94) |
| Democratic People's Republic of Korea | 43 (29 to 78) | 0.34 (0.22 to 0.65) | 95 (67 to 160) | 0.35 (0.24 to 0.58) | 0.07 (-0.01 to 0.15) |
| Democratic Republic of the Congo | 58 (36 to 105) | 0.23 (0.10 to 0.46) | 86 (36 to 161) | 0.19 (0.07 to 0.39) | -0.58 (-0.64 to -0.52) |
| Denmark | 17 (15 to 18) | 0.23 (0.21 to 0.25) | 21 (18 to 23) | 0.18 (0.15 to 0.19) | -0.93 (-1.30 to -0.55) |
| Djibouti | 1 (0 to 1) | 0.23 (0.11 to 0.49) | 1 (1 to 2) | 0.15 (0.07 to 0.29) | -1.24 (-1.57 to -0.90) |
| Dominica | 0 (0 to 0) | 0.18 (0.11 to 0.32) | 0 (0 to 0) | 0.09 (0.07 to 0.23) | -2.01 (-2.19 to -1.83) |
| Dominican Republic | 12 (9 to 16) | 0.17 (0.13 to 0.32) | 9 (6 to 23) | 0.08 (0.06 to 0.23) | -2.23 (-2.52 to -1.93) |
| Ecuador | 18 (14 to 22) | 0.27 (0.20 to 0.34) | 27 (23 to 32) | 0.18 (0.15 to 0.21) | -1.38 (-1.73 to -1.02) |
| Egypt | 727 (365 to 1033) | 1.17 (0.77 to 1.55) | 282 (220 to 377) | 0.36 (0.28 to 0.50) | -3.72 (-4.02 to -3.41) |
| El Salvador | 12 (9 to 16) | 0.29 (0.20 to 0.38) | 8 (5 to 16) | 0.12 (0.08 to 0.24) | -2.87 (-3.08 to -2.66) |
| Equatorial Guinea | 1 (0 to 1) | 0.26 (0.12 to 0.55) | 1 (0 to 1) | 0.11 (0.06 to 0.20) | -2.68 (-2.91 to -2.44) |
| Eritrea | 7 (4 to 14) | 0.30 (0.14 to 0.66) | 9 (4 to 16) | 0.21 (0.09 to 0.43) | -1.14 (-1.23 to -1.04) |
| Estonia | 0 (0 to 0) | 0.02 (0.02 to 0.03) | 1 (1 to 1) | 0.04 (0.03 to 0.04) | 1.67 (-0.19 to 3.55) |
| Eswatini | 1 (1 to 1) | 0.18 (0.12 to 0.29) | 1 (1 to 1) | 0.13 (0.09 to 0.18) | -1.02 (-1.16 to -0.89) |
| Ethiopia | 92 (48 to 174) | 0.27 (0.10 to 0.54) | 97 (38 to 203) | 0.15 (0.05 to 0.30) | -1.93 (-2.01 to -1.84) |
| Fiji | 1 (1 to 2) | 0.21 (0.13 to 0.44) | 1 (1 to 2) | 0.15 (0.08 to 0.32) | -1.05 (-1.13 to -0.97) |
| Finland | 6 (5 to 7) | 0.09 (0.08 to 0.10) | 8 (7 to 9) | 0.06 (0.05 to 0.07) | -1.13 (-1.38 to -0.88) |
| France | 239 (191 to 296) | 0.30 (0.24 to 0.37) | 325 (279 to 361) | 0.21 (0.19 to 0.23) | -1.15 (-1.36 to -0.94) |
| Gabon | 1 (1 to 3) | 0.21 (0.10 to 0.45) | 1 (1 to 3) | 0.13 (0.07 to 0.24) | -1.51 (-1.72 to -1.29) |
| Gambia | 1 (1 to 2) | 0.23 (0.07 to 0.46) | 3 (1 to 5) | 0.19 (0.07 to 0.34) | -0.73 (-1.18 to -0.28) |
| Georgia | 21 (16 to 27) | 0.37 (0.29 to 0.47) | 58 (45 to 73) | 1.01 (0.79 to 1.27) | 3.21 (2.45 to 3.98) |
| Germany | 304 (254 to 350) | 0.27 (0.23 to 0.31) | 544 (453 to 604) | 0.26 (0.22 to 0.28) | -0.14 (-0.57 to 0.29) |
| Ghana | 19 (11 to 37) | 0.22 (0.10 to 0.46) | 26 (14 to 44) | 0.13 (0.06 to 0.23) | -1.70 (-1.79 to -1.61) |
| Greece | 32 (29 to 34) | 0.24 (0.23 to 0.26) | 87 (76 to 98) | 0.32 (0.29 to 0.36) | 0.89 (0.62 to 1.16) |
| Greenland | 0 (0 to 0) | 0.77 (0.38 to 0.99) | 0 (0 to 0) | 0.19 (0.15 to 0.30) | -4.36 (-4.90 to -3.82) |
| Grenada | 1 (1 to 1) | 0.81 (0.68 to 0.98) | 0 (0 to 0) | 0.26 (0.22 to 0.29) | -3.56 (-4.13 to -2.98) |
| Guam | 0 (0 to 0) | 0.15 (0.10 to 0.35) | 0 (0 to 0) | 0.06 (0.03 to 0.12) | -3.26 (-3.83 to -2.68) |
| Guatemala | 14 (12 to 16) | 0.26 (0.22 to 0.30) | 7 (6 to 8) | 0.06 (0.05 to 0.07) | -4.84 (-5.33 to -4.35) |
| Guinea | 13 (7 to 27) | 0.26 (0.09 to 0.57) | 19 (9 to 32) | 0.21 (0.08 to 0.41) | -0.64 (-0.75 to -0.53) |
| Guinea-Bissau | 2 (1 to 4) | 0.32 (0.13 to 0.60) | 3 (1 to 5) | 0.23 (0.10 to 0.42) | -1.05 (-1.16 to -0.95) |
| Guyana | 1 (1 to 1) | 0.17 (0.15 to 0.19) | 3 (2 to 3) | 0.39 (0.30 to 0.50) | 2.88 (2.38 to 3.38) |
| Haiti | 55 (21 to 95) | 0.87 (0.43 to 1.27) | 53 (27 to 87) | 0.49 (0.28 to 0.73) | -1.79 (-1.88 to -1.71) |
| Honduras | 7 (4 to 10) | 0.26 (0.14 to 0.41) | 12 (8 to 18) | 0.20 (0.12 to 0.32) | -0.86 (-1.02 to -0.70) |
| Hungary | 43 (38 to 49) | 0.32 (0.28 to 0.37) | 31 (25 to 38) | 0.16 (0.13 to 0.21) | -2.14 (-2.90 to -1.38) |
| Iceland | 0 (0 to 0) | 0.15 (0.14 to 0.17) | 1 (1 to 1) | 0.13 (0.11 to 0.15) | -0.64 (-1.06 to -0.21) |
| India | 1756 (1132 to 2495) | 0.29 (0.17 to 0.46) | 2612 (1739 to 4060) | 0.24 (0.15 to 0.39) | -0.65 (-1.11 to -0.19) |
| Indonesia | 185 (106 to 426) | 0.14 (0.08 to 0.42) | 268 (170 to 672) | 0.12 (0.07 to 0.37) | -0.53 (-0.60 to -0.46) |
| Iran (Islamic Republic of) | 471 (307 to 593) | 1.40 (0.89 to 1.94) | 429 (286 to 494) | 0.61 (0.40 to 0.70) | -2.62 (-2.76 to -2.48) |
| Iraq | 37 (21 to 57) | 0.24 (0.13 to 0.49) | 38 (20 to 80) | 0.15 (0.08 to 0.38) | -1.60 (-1.73 to -1.47) |
| Ireland | 6 (5 to 6) | 0.15 (0.14 to 0.16) | 8 (7 to 9) | 0.11 (0.10 to 0.12) | -1.03 (-1.31 to -0.75) |
| Israel | 22 (19 to 29) | 0.48 (0.41 to 0.61) | 35 (30 to 39) | 0.27 (0.23 to 0.30) | -1.80 (-1.96 to -1.63) |
| Italy | 171 (159 to 179) | 0.21 (0.20 to 0.23) | 138 (115 to 153) | 0.09 (0.08 to 0.10) | -2.84 (-3.12 to -2.55) |
| Jamaica | 4 (4 to 5) | 0.20 (0.18 to 0.23) | 2 (2 to 3) | 0.07 (0.05 to 0.09) | -3.30 (-4.06 to -2.53) |
| Japan | 385 (366 to 401) | 0.29 (0.28 to 0.30) | 1002 (786 to 1147) | 0.29 (0.25 to 0.32) | -0.05 (-0.37 to 0.27) |
| Jordan | 2 (1 to 7) | 0.06 (0.03 to 0.29) | 4 (2 to 14) | 0.06 (0.02 to 0.20) | -0.03 (-0.43 to 0.38) |
| Kazakhstan | 6 (5 to 7) | 0.05 (0.03 to 0.06) | 7 (6 to 9) | 0.05 (0.04 to 0.06) | 0.08 (-0.24 to 0.40) |
| Kenya | 19 (10 to 40) | 0.14 (0.05 to 0.32) | 33 (14 to 65) | 0.12 (0.05 to 0.23) | -0.55 (-0.69 to -0.42) |
| Kiribati | 0 (0 to 0) | 0.30 (0.17 to 0.68) | 0 (0 to 0) | 0.25 (0.16 to 0.57) | -0.57 (-0.62 to -0.52) |
| Kuwait | 1 (1 to 1) | 0.07 (0.07 to 0.08) | 4 (3 to 5) | 0.14 (0.12 to 0.16) | 2.40 (0.11 to 4.73) |
| Kyrgyzstan | 3 (3 to 4) | 0.09 (0.08 to 0.11) | 2 (2 to 3) | 0.05 (0.04 to 0.06) | -2.16 (-2.89 to -1.43) |
| Lao People's Democratic Republic | 8 (3 to 19) | 0.23 (0.11 to 0.54) | 9 (5 to 20) | 0.15 (0.09 to 0.36) | -1.30 (-1.39 to -1.21) |
| Latvia | 1 (1 to 1) | 0.02 (0.02 to 0.03) | 4 (4 to 5) | 0.10 (0.09 to 0.12) | 4.71 (3.42 to 6.01) |
| Lebanon | 21 (13 to 31) | 0.89 (0.55 to 1.34) | 23 (17 to 38) | 0.37 (0.28 to 0.59) | -2.76 (-2.98 to -2.54) |
| Lesotho | 1 (1 to 2) | 0.14 (0.09 to 0.24) | 2 (1 to 3) | 0.14 (0.10 to 0.20) | 0.06 (-0.23 to 0.34) |
| Liberia | 5 (3 to 10) | 0.27 (0.11 to 0.53) | 6 (3 to 11) | 0.19 (0.07 to 0.36) | -1.21 (-1.51 to -0.91) |
| Libya | 15 (9 to 27) | 0.32 (0.20 to 0.64) | 18 (8 to 31) | 0.37 (0.17 to 0.64) | 0.51 (0.00 to 1.01) |
| Lithuania | 1 (1 to 2) | 0.04 (0.03 to 0.04) | 4 (4 to 5) | 0.08 (0.07 to 0.09) | 2.58 (0.61 to 4.58) |
| Luxembourg | 1 (1 to 1) | 0.27 (0.25 to 0.29) | 2 (2 to 3) | 0.21 (0.18 to 0.23) | -0.87 (-1.15 to -0.59) |
| Madagascar | 38 (23 to 75) | 0.42 (0.20 to 0.80) | 58 (31 to 112) | 0.31 (0.15 to 0.61) | -0.94 (-1.00 to -0.87) |
| Malawi | 17 (10 to 32) | 0.22 (0.10 to 0.43) | 19 (9 to 33) | 0.16 (0.07 to 0.30) | -0.92 (-1.18 to -0.65) |
| Malaysia | 10 (7 to 30) | 0.08 (0.05 to 0.30) | 19 (11 to 65) | 0.07 (0.04 to 0.26) | -0.65 (-1.15 to -0.14) |
| Maldives | 1 (1 to 1) | 0.52 (0.37 to 0.69) | 1 (1 to 1) | 0.19 (0.15 to 0.30) | -3.25 (-3.38 to -3.11) |
| Mali | 14 (8 to 30) | 0.23 (0.08 to 0.55) | 27 (13 to 53) | 0.18 (0.06 to 0.38) | -0.76 (-0.93 to -0.60) |
| Malta | 0 (0 to 0) | 0.09 (0.08 to 0.10) | 1 (1 to 1) | 0.08 (0.07 to 0.10) | -0.38 (-0.77 to 0.01) |
| Marshall Islands | 0 (0 to 0) | 0.33 (0.20 to 0.69) | 0 (0 to 0) | 0.23 (0.14 to 0.50) | -1.14 (-1.19 to -1.08) |
| Mauritania | 3 (1 to 6) | 0.20 (0.09 to 0.46) | 4 (2 to 7) | 0.14 (0.06 to 0.28) | -1.15 (-1.45 to -0.85) |
| Mauritius | 2 (2 to 2) | 0.26 (0.24 to 0.28) | 11 (10 to 12) | 0.71 (0.63 to 0.77) | 3.38 (1.84 to 4.93) |
| Mexico | 83 (75 to 100) | 0.15 (0.13 to 0.17) | 101 (89 to 112) | 0.09 (0.08 to 0.10) | -1.66 (-2.17 to -1.15) |
| Micronesia (Federated States of) | 0 (0 to 0) | 0.36 (0.22 to 0.80) | 0 (0 to 0) | 0.24 (0.15 to 0.55) | -1.30 (-1.34 to -1.25) |
| Monaco | 0 (0 to 0) | 0.18 (0.13 to 0.25) | 0 (0 to 0) | 0.15 (0.10 to 0.21) | -0.56 (-0.62 to -0.49) |
| Mongolia | 20 (11 to 32) | 1.73 (0.99 to 2.86) | 34 (20 to 44) | 1.59 (0.91 to 2.05) | -0.29 (-0.65 to 0.06) |
| Montenegro | 0 (0 to 1) | 0.04 (0.02 to 0.24) | 0 (0 to 2) | 0.03 (0.02 to 0.20) | -0.77 (-1.07 to -0.47) |
| Morocco | 72 (40 to 126) | 0.33 (0.18 to 0.67) | 131 (50 to 201) | 0.43 (0.15 to 0.67) | 0.92 (0.78 to 1.06) |
| Mozambique | 26 (14 to 47) | 0.27 (0.11 to 0.60) | 40 (18 to 70) | 0.24 (0.09 to 0.46) | -0.39 (-0.51 to -0.27) |
| Myanmar | 70 (37 to 151) | 0.22 (0.12 to 0.55) | 75 (48 to 178) | 0.15 (0.10 to 0.42) | -1.14 (-1.22 to -1.06) |
| Namibia | 1 (1 to 2) | 0.16 (0.11 to 0.23) | 2 (1 to 3) | 0.13 (0.09 to 0.19) | -0.60 (-0.71 to -0.49) |
| Nauru | 0 (0 to 0) | 0.31 (0.20 to 0.63) | 0 (0 to 0) | 0.37 (0.24 to 1.00) | 0.50 (0.39 to 0.61) |
| Nepal | 54 (31 to 85) | 0.38 (0.17 to 0.67) | 68 (34 to 117) | 0.31 (0.15 to 0.58) | -0.66 (-0.74 to -0.59) |
| Netherlands | 30 (28 to 33) | 0.16 (0.15 to 0.17) | 48 (41 to 53) | 0.14 (0.12 to 0.15) | -0.56 (-1.06 to -0.06) |
| New Zealand | 5 (5 to 6) | 0.14 (0.13 to 0.15) | 8 (7 to 8) | 0.10 (0.09 to 0.11) | -1.04 (-2.02 to -0.05) |
| Nicaragua | 4 (2 to 7) | 0.14 (0.07 to 0.30) | 2 (1 to 9) | 0.05 (0.03 to 0.19) | -3.51 (-3.73 to -3.28) |
| Niger | 15 (7 to 27) | 0.28 (0.08 to 0.70) | 31 (11 to 61) | 0.23 (0.05 to 0.52) | -0.63 (-0.79 to -0.48) |
| Nigeria | 161 (92 to 296) | 0.25 (0.09 to 0.52) | 228 (153 to 332) | 0.14 (0.07 to 0.23) | -1.79 (-1.91 to -1.67) |
| Niue | 0 (0 to 0) | 0.26 (0.17 to 0.61) | 0 (0 to 0) | 0.23 (0.14 to 0.52) | -0.28 (-0.61 to 0.05) |
| North Macedonia | 3 (2 to 4) | 0.15 (0.10 to 0.26) | 3 (2 to 6) | 0.12 (0.07 to 0.23) | -0.58 (-1.32 to 0.17) |
| Northern Mariana Islands | 0 (0 to 0) | 0.13 (0.08 to 0.30) | 0 (0 to 0) | 0.11 (0.07 to 0.28) | -0.56 (-0.70 to -0.43) |
| Norway | 7 (7 to 8) | 0.14 (0.13 to 0.15) | 5 (4 to 5) | 0.05 (0.05 to 0.05) | -3.59 (-5.64 to -1.49) |
| Oman | 1 (1 to 4) | 0.11 (0.05 to 0.39) | 3 (1 to 5) | 0.11 (0.04 to 0.24) | 0.14 (-0.39 to 0.67) |
| Pakistan | 311 (190 to 458) | 0.34 (0.18 to 0.55) | 485 (311 to 747) | 0.32 (0.18 to 0.56) | -0.20 (-0.37 to -0.03) |
| Palau | 0 (0 to 0) | 0.19 (0.11 to 0.42) | 0 (0 to 0) | 0.13 (0.06 to 0.30) | -1.21 (-1.29 to -1.12) |
| Palestine | 4 (2 to 8) | 0.23 (0.10 to 0.59) | 4 (2 to 11) | 0.13 (0.05 to 0.41) | -1.86 (-2.02 to -1.69) |
| Panama | 3 (3 to 4) | 0.19 (0.16 to 0.21) | 3 (2 to 3) | 0.06 (0.05 to 0.07) | -3.68 (-4.10 to -3.27) |
| Papua New Guinea | 9 (5 to 14) | 0.30 (0.18 to 0.58) | 20 (13 to 36) | 0.27 (0.17 to 0.55) | -0.31 (-0.42 to -0.21) |
| Paraguay | 2 (2 to 7) | 0.09 (0.05 to 0.27) | 4 (3 to 12) | 0.07 (0.04 to 0.22) | -0.54 (-0.77 to -0.31) |
| Peru | 42 (29 to 57) | 0.25 (0.19 to 0.32) | 43 (31 to 63) | 0.13 (0.09 to 0.18) | -2.19 (-2.93 to -1.44) |
| Philippines | 52 (34 to 115) | 0.11 (0.06 to 0.33) | 70 (48 to 184) | 0.08 (0.05 to 0.24) | -1.14 (-1.54 to -0.73) |
| Poland | 69 (60 to 79) | 0.17 (0.14 to 0.19) | 77 (69 to 84) | 0.12 (0.11 to 0.13) | -1.14 (-1.48 to -0.81) |
| Portugal | 28 (26 to 30) | 0.25 (0.23 to 0.26) | 56 (48 to 63) | 0.22 (0.19 to 0.24) | -0.46 (-1.13 to 0.20) |
| Puerto Rico | 21 (19 to 23) | 0.60 (0.55 to 0.65) | 6 (5 to 7) | 0.10 (0.08 to 0.12) | -5.69 (-6.36 to -5.02) |
| Qatar | 0 (0 to 1) | 0.17 (0.11 to 0.38) | 1 (1 to 2) | 0.13 (0.07 to 0.19) | -0.90 (-1.84 to 0.05) |
| Republic of Korea | 42 (29 to 72) | 0.13 (0.09 to 0.27) | 39 (22 to 136) | 0.06 (0.03 to 0.17) | -2.67 (-2.89 to -2.45) |
| Republic of Moldova | 0 (0 to 0) | 0.00 (0.00 to 0.00) | 0 (0 to 0) | 0.01 (0.01 to 0.01) | 3.96 (2.83 to 5.10) |
| Romania | 120 (96 to 144) | 0.48 (0.39 to 0.57) | 170 (146 to 195) | 0.49 (0.42 to 0.55) | -0.04 (-0.39 to 0.31) |
| Russian Federation | 515 (465 to 600) | 0.34 (0.31 to 0.39) | 221 (204 to 238) | 0.10 (0.10 to 0.11) | -3.70 (-4.43 to -2.97) |
| Rwanda | 18 (11 to 39) | 0.35 (0.17 to 0.75) | 14 (6 to 30) | 0.17 (0.07 to 0.37) | -2.29 (-2.54 to -2.03) |
| Saint Kitts and Nevis | 0 (0 to 0) | 0.21 (0.18 to 0.28) | 0 (0 to 0) | 0.07 (0.06 to 0.09) | -3.19 (-4.13 to -2.25) |
| Saint Lucia | 1 (1 to 1) | 0.80 (0.72 to 0.89) | 1 (0 to 1) | 0.24 (0.20 to 0.29) | -3.82 (-4.12 to -3.53) |
| Saint Vincent and the Grenadines | 0 (0 to 0) | 0.09 (0.08 to 0.10) | 0 (0 to 0) | 0.06 (0.05 to 0.07) | -1.08 (-1.54 to -0.62) |
| Samoa | 0 (0 to 1) | 0.27 (0.18 to 0.61) | 0 (0 to 1) | 0.20 (0.13 to 0.47) | -1.00 (-1.08 to -0.93) |
| San Marino | 0 (0 to 0) | 0.10 (0.07 to 0.22) | 0 (0 to 0) | 0.05 (0.03 to 0.12) | -2.52 (-2.90 to -2.12) |
| Sao Tome and Principe | 0 (0 to 0) | 0.16 (0.06 to 0.37) | 0 (0 to 0) | 0.13 (0.05 to 0.24) | -0.71 (-0.86 to -0.56) |
| Saudi Arabia | 14 (9 to 31) | 0.14 (0.08 to 0.39) | 18 (9 to 36) | 0.08 (0.04 to 0.20) | -1.79 (-2.18 to -1.39) |
| Senegal | 13 (7 to 25) | 0.25 (0.09 to 0.49) | 18 (7 to 32) | 0.18 (0.06 to 0.33) | -1.02 (-1.20 to -0.84) |
| Serbia | 13 (8 to 32) | 0.14 (0.09 to 0.39) | 15 (9 to 30) | 0.09 (0.06 to 0.19) | -1.37 (-1.60 to -1.13) |
| Seychelles | 0 (0 to 0) | 0.12 (0.07 to 0.36) | 0 (0 to 0) | 0.08 (0.04 to 0.25) | -1.28 (-1.40 to -1.16) |
| Sierra Leone | 10 (6 to 18) | 0.27 (0.12 to 0.49) | 12 (7 to 21) | 0.20 (0.08 to 0.34) | -1.01 (-1.17 to -0.85) |
| Singapore | 6 (5 to 6) | 0.23 (0.21 to 0.24) | 6 (5 to 7) | 0.09 (0.08 to 0.10) | -3.08 (-3.83 to -2.31) |
| Slovakia | 6 (4 to 12) | 0.10 (0.06 to 0.23) | 6 (3 to 11) | 0.06 (0.04 to 0.13) | -1.55 (-1.79 to -1.31) |
| Slovenia | 1 (1 to 1) | 0.04 (0.03 to 0.04) | 2 (1 to 2) | 0.04 (0.03 to 0.04) | -0.12 (-0.79 to 0.56) |
| Solomon Islands | 0 (0 to 1) | 0.26 (0.14 to 0.54) | 1 (0 to 1) | 0.22 (0.13 to 0.51) | -0.59 (-0.70 to -0.48) |
| Somalia | 20 (8 to 37) | 0.36 (0.11 to 0.75) | 34 (9 to 73) | 0.27 (0.06 to 0.58) | -0.98 (-1.11 to -0.86) |
| South Africa | 29 (24 to 37) | 0.11 (0.08 to 0.14) | 46 (35 to 55) | 0.09 (0.07 to 0.11) | -0.46 (-0.65 to -0.28) |
| South Sudan | 15 (8 to 29) | 0.32 (0.14 to 0.68) | 17 (9 to 30) | 0.24 (0.09 to 0.44) | -0.98 (-1.15 to -0.81) |
| Spain | 131 (121 to 140) | 0.27 (0.25 to 0.29) | 241 (200 to 270) | 0.22 (0.19 to 0.24) | -0.67 (-1.00 to -0.33) |
| Sri Lanka | 55 (39 to 92) | 0.42 (0.32 to 0.70) | 101 (63 to 148) | 0.42 (0.27 to 0.60) | 0.11 (-0.31 to 0.54) |
| Sudan | 72 (37 to 132) | 0.37 (0.20 to 0.70) | 124 (60 to 183) | 0.42 (0.18 to 0.61) | 0.39 (0.22 to 0.57) |
| Suriname | 2 (1 to 3) | 0.74 (0.49 to 0.90) | 2 (1 to 3) | 0.32 (0.24 to 0.47) | -2.61 (-2.93 to -2.28) |
| Sweden | 17 (16 to 19) | 0.14 (0.13 to 0.15) | 26 (22 to 29) | 0.12 (0.11 to 0.14) | -0.44 (-1.01 to 0.13) |
| Switzerland | 35 (30 to 43) | 0.36 (0.31 to 0.44) | 37 (31 to 42) | 0.19 (0.16 to 0.21) | -2.11 (-2.46 to -1.76) |
| Syrian Arab Republic | 10 (5 to 38) | 0.09 (0.05 to 0.43) | 6 (3 to 33) | 0.06 (0.03 to 0.29) | -1.72 (-2.30 to -1.13) |
| Taiwan (Province of China) | 13 (12 to 13) | 0.10 (0.09 to 0.10) | 77 (66 to 85) | 0.18 (0.16 to 0.20) | 1.76 (0.49 to 3.04) |
| Tajikistan | 41 (25 to 54) | 1.21 (0.74 to 1.66) | 53 (34 to 73) | 0.81 (0.53 to 1.09) | -1.36 (-1.82 to -0.90) |
| Thailand | 40 (25 to 123) | 0.09 (0.06 to 0.38) | 62 (35 to 282) | 0.07 (0.04 to 0.28) | -0.90 (-1.20 to -0.60) |
| Timor-Leste | 1 (1 to 3) | 0.19 (0.09 to 0.49) | 2 (1 to 4) | 0.15 (0.08 to 0.43) | -0.68 (-0.90 to -0.47) |
| Togo | 5 (3 to 9) | 0.22 (0.09 to 0.45) | 9 (4 to 15) | 0.18 (0.07 to 0.31) | -0.72 (-0.89 to -0.54) |
| Tokelau | 0 (0 to 0) | 0.29 (0.18 to 0.61) | 0 (0 to 0) | 0.29 (0.18 to 0.73) | 0.18 (-0.00 to 0.36) |
| Tonga | 0 (0 to 0) | 0.20 (0.13 to 0.45) | 0 (0 to 0) | 0.14 (0.09 to 0.34) | -1.10 (-1.28 to -0.92) |
| Trinidad and Tobago | 5 (4 to 5) | 0.47 (0.44 to 0.52) | 3 (2 to 4) | 0.18 (0.14 to 0.24) | -2.77 (-3.14 to -2.40) |
| Tunisia | 15 (10 to 28) | 0.25 (0.16 to 0.54) | 40 (12 to 67) | 0.36 (0.11 to 0.60) | 1.11 (0.79 to 1.43) |
| Türkiye | 538 (331 to 724) | 1.20 (0.79 to 1.65) | 408 (313 to 562) | 0.50 (0.39 to 0.69) | -2.72 (-3.08 to -2.35) |
| Turkmenistan | 4 (3 to 5) | 0.18 (0.13 to 0.24) | 9 (7 to 12) | 0.23 (0.17 to 0.31) | 0.75 (0.46 to 1.04) |
| Tuvalu | 0 (0 to 0) | 0.37 (0.22 to 0.77) | 0 (0 to 0) | 0.23 (0.15 to 0.52) | -1.58 (-1.64 to -1.52) |
| Uganda | 37 (20 to 68) | 0.28 (0.10 to 0.61) | 41 (22 to 72) | 0.15 (0.07 to 0.28) | -2.03 (-2.24 to -1.81) |
| Ukraine | 41 (37 to 46) | 0.07 (0.06 to 0.07) | 42 (31 to 53) | 0.07 (0.05 to 0.09) | -0.01 (-1.23 to 1.24) |
| United Arab Emirates | 6 (3 to 10) | 0.73 (0.38 to 1.09) | 10 (5 to 12) | 0.38 (0.20 to 0.53) | -2.07 (-4.13 to 0.02) |
| United Kingdom | 106 (96 to 132) | 0.14 (0.13 to 0.18) | 119 (107 to 126) | 0.10 (0.10 to 0.11) | -1.10 (-1.37 to -0.84) |
| United Republic of Tanzania | 47 (28 to 89) | 0.26 (0.11 to 0.52) | 62 (31 to 113) | 0.16 (0.07 to 0.31) | -1.52 (-1.66 to -1.37) |
| United States of America | 971 (855 to 1070) | 0.32 (0.28 to 0.35) | 1785 (1535 to 1945) | 0.31 (0.27 to 0.33) | -0.09 (-0.27 to 0.08) |
| United States Virgin Islands | 0 (0 to 0) | 0.42 (0.24 to 0.53) | 0 (0 to 0) | 0.16 (0.11 to 0.26) | -3.01 (-3.46 to -2.56) |
| Uruguay | 8 (7 to 9) | 0.23 (0.20 to 0.25) | 8 (7 to 9) | 0.16 (0.14 to 0.17) | -1.30 (-1.49 to -1.11) |
| Uzbekistan | 68 (54 to 85) | 0.46 (0.34 to 0.61) | 97 (77 to 119) | 0.37 (0.30 to 0.46) | -0.68 (-1.10 to -0.25) |
| Vanuatu | 0 (0 to 0) | 0.28 (0.17 to 0.57) | 0 (0 to 1) | 0.22 (0.14 to 0.52) | -0.70 (-0.87 to -0.53) |
| Venezuela (Bolivarian Republic of) | 14 (12 to 16) | 0.11 (0.09 to 0.12) | 12 (9 to 15) | 0.04 (0.03 to 0.06) | -2.84 (-3.50 to -2.17) |
| Viet Nam | 70 (38 to 194) | 0.15 (0.07 to 0.50) | 106 (53 to 332) | 0.12 (0.06 to 0.40) | -0.78 (-0.82 to -0.74) |
| Yemen | 44 (21 to 73) | 0.40 (0.16 to 0.81) | 106 (49 to 160) | 0.55 (0.21 to 0.90) | 0.99 (0.63 to 1.34) |
| Zambia | 12 (7 to 25) | 0.24 (0.12 to 0.49) | 26 (9 to 52) | 0.24 (0.08 to 0.49) | 0.11 (-0.01 to 0.23) |
| Zimbabwe | 8 (4 to 14) | 0.15 (0.08 to 0.25) | 18 (12 to 27) | 0.18 (0.12 to 0.26) | 0.69 (0.17 to 1.21) |

**Supplemental Table 4. DALYs of PAH between 1990 and 2021 at the global and regional levels.**

| **Location** | **DALYs (95% UI)** | | | | **AAPC (95% CI)** |
| --- | --- | --- | --- | --- | --- |
|  | **DALY cases in 1990** | **ASDR in 1990 (per 100,000)** | **DALY cases in 2021** | **ASDR in 2021 (per 100,000)** |  |
| **Global** | 687419 (535241 to 813086) | 13.21 (10.78 to 15.36) | 642104 (552273 to 728993) | 8.24 (7.14 to 9.39) | -1.52 (-1.64 to -1.40) |
| **Socio-demographic index** |  |  |  |  |  |
| High SDI | 81792 (77185 to 88110) | 9.16 (8.71 to 9.90) | 93182 (84873 to 99192) | 6.16 (5.76 to 6.49) | -1.29 (-1.45 to -1.14) |
| High-middle SDI | 127638 (106711 to 154438) | 13.14 (10.91 to 16.04) | 99448 (85757 to 117639) | 6.48 (5.61 to 7.87) | -2.27 (-2.48 to -2.07) |
| Middle SDI | 210946 (172857 to 258349) | 14.29 (11.66 to 17.69) | 197171 (148781 to 232321) | 8.23 (6.26 to 9.70) | -1.78 (-1.92 to -1.64) |
| Low-middle SDI | 195281 (117185 to 245591) | 14.92 (10.74 to 18.41) | 156400 (122426 to 194166) | 9.07 (7.05 to 11.60) | -1.59 (-1.70 to -1.47) |
| Low SDI | 71125 (48628 to 111614) | 12.42 (7.78 to 19.19) | 95342 (67471 to 133050) | 9.30 (6.08 to 13.20) | -0.92 (-1.04 to -0.80) |
| **Super regions** |  |  |  |  |  |
| Central Europe, Eastern Europe, and Central Asia | 40726 (36800 to 44820) | 9.62 (8.71 to 10.50) | 30399 (27840 to 33545) | 6.21 (5.65 to 6.92) | -1.44 (-1.83 to -1.06) |
| High-income | 92777 (86818 to 100017) | 10.08 (9.49 to 10.86) | 98576 (89661 to 104138) | 6.44 (6.07 to 6.74) | -1.43 (-1.60 to -1.26) |
| Latin America and Caribbean | 41479 (34569 to 49269) | 11.11 (9.58 to 12.79) | 40095 (36416 to 44567) | 6.90 (6.18 to 7.78) | -1.54 (-1.70 to -1.38) |
| North Africa and Middle East | 145728 (74879 to 204070) | 35.84 (21.25 to 46.14) | 80753 (58086 to 98810) | 14.81 (10.76 to 17.96) | -2.81 (-2.93 to -2.70) |
| South Asia | 136086 (77808 to 184381) | 12.24 (7.79 to 17.12) | 136563 (97809 to 189353) | 8.54 (6.02 to 12.46) | -1.12 (-1.32 to -0.93) |
| Southeast Asia, East Asia, and Oceania | 180087 (137972 to 254840) | 13.16 (10.25 to 18.58) | 187307 (144098 to 242552) | 8.01 (6.32 to 10.66) | -1.60 (-1.84 to -1.36) |
| Sub-Saharan Africa | 50537 (34106 to 97293) | 9.51 (5.63 to 17.43) | 68412 (43351 to 109821) | 6.45 (3.58 to 10.30) | -1.25 (-1.29 to -1.21) |
| **Regions** |  |  |  |  |  |
| Andean Latin America | 5049 (2977 to 7422) | 11.92 (7.83 to 16.43) | 3544 (2795 to 4490) | 5.73 (4.52 to 7.28) | -2.24 (-2.44 to -2.04) |
| Australasia | 1514 (1337 to 1856) | 7.37 (6.49 to 9.06) | 1434 (1305 to 1563) | 3.67 (3.38 to 3.99) | -2.06 (-2.46 to -1.66) |
| Caribbean | 7398 (4170 to 11145) | 19.72 (11.78 to 28.82) | 5071 (2988 to 7877) | 11.73 (6.48 to 18.74) | -1.63 (-1.86 to -1.40) |
| Central Asia | 9071 (7195 to 10646) | 14.14 (11.27 to 16.45) | 11619 (9514 to 14202) | 12.91 (10.61 to 15.60) | -0.30 (-0.64 to 0.03) |
| Central Europe | 11026 (9784 to 12154) | 8.15 (7.25 to 8.95) | 10424 (9512 to 11459) | 6.05 (5.50 to 6.67) | -1.02 (-1.31 to -0.73) |
| Central Latin America | 9966 (8844 to 11855) | 6.12 (5.40 to 7.16) | 7246 (6391 to 8407) | 3.00 (2.63 to 3.51) | -2.26 (-2.54 to -1.97) |
| Central Sub-Saharan Africa | 5367 (3258 to 10525) | 9.09 (5.65 to 17.13) | 6524 (3586 to 11025) | 6.16 (2.94 to 11.09) | -1.25 (-1.30 to -1.21) |
| East Asia | 151596 (117394 to 205773) | 15.78 (12.34 to 21.21) | 154740 (102939 to 190399) | 8.84 (5.99 to 11.01) | -1.86 (-2.17 to -1.55) |
| Eastern Europe | 20628 (18781 to 23860) | 9.23 (8.47 to 10.49) | 8357 (7760 to 8996) | 3.32 (3.10 to 3.56) | -3.22 (-4.10 to -2.34) |
| Eastern Sub-Saharan Africa | 23148 (14694 to 45765) | 11.11 (6.18 to 21.17) | 26605 (14265 to 49615) | 6.85 (3.30 to 12.62) | -1.55 (-1.62 to -1.47) |
| High-income Asia Pacific | 18474 (17689 to 19467) | 12.03 (11.43 to 12.79) | 19988 (17442 to 21997) | 8.31 (7.74 to 8.87) | -1.19 (-1.65 to -0.73) |
| High-income North America | 30206 (27736 to 32715) | 10.13 (9.39 to 10.99) | 38373 (35060 to 40844) | 7.71 (7.17 to 8.18) | -0.86 (-1.07 to -0.65) |
| North Africa and Middle East | 145728 (74879 to 204070) | 35.84 (21.25 to 46.14) | 80753 (58086 to 98810) | 14.81 (10.76 to 17.96) | -2.81 (-2.93 to -2.70) |
| Oceania | 704 (448 to 1090) | 10.85 (7.02 to 17.55) | 1455 (988 to 2411) | 10.14 (6.90 to 17.10) | -0.20 (-0.42 to 0.02) |
| South Asia | 136086 (77808 to 184381) | 12.24 (7.79 to 17.12) | 136563 (97809 to 189353) | 8.54 (6.02 to 12.46) | -1.12 (-1.32 to -0.93) |
| Southeast Asia | 27786 (18242 to 51943) | 6.31 (4.28 to 12.72) | 31112 (22913 to 58708) | 4.65 (3.39 to 9.25) | -0.97 (-1.01 to -0.93) |
| Southern Latin America | 7983 (7181 to 8803) | 16.28 (14.65 to 17.95) | 4739 (4439 to 5092) | 6.55 (6.13 to 7.07) | -2.91 (-3.42 to -2.39) |
| Southern Sub-Saharan Africa | 2188 (1667 to 2875) | 4.70 (3.65 to 6.37) | 3212 (2359 to 3891) | 4.33 (3.21 to 5.20) | -0.29 (-0.48 to -0.09) |
| Tropical Latin America | 19065 (17856 to 20357) | 14.18 (13.35 to 14.98) | 24235 (23002 to 25403) | 10.22 (9.65 to 10.78) | -1.06 (-1.42 to -0.70) |
| Western Europe | 34599 (31743 to 38281) | 8.27 (7.69 to 9.18) | 34043 (31024 to 36440) | 5.05 (4.75 to 5.32) | -1.56 (-1.77 to -1.35) |
| Western Sub-Saharan Africa | 19834 (13553 to 38853) | 9.34 (5.22 to 16.94) | 32071 (22084 to 47222) | 6.50 (3.77 to 9.47) | -1.16 (-1.21 to -1.11) |
| **Countries and territories** |  |  |  |  |  |
| Afghanistan | 2360 (1186 to 3637) | 18.19 (9.20 to 29.19) | 8110 (3768 to 12131) | 25.28 (10.98 to 39.03) | 1.07 (0.78 to 1.36) |
| Albania | 458 (291 to 581) | 16.10 (10.58 to 20.33) | 287 (173 to 512) | 8.43 (5.32 to 14.16) | -2.10 (-2.56 to -1.64) |
| Algeria | 3350 (2116 to 5773) | 12.33 (8.00 to 22.02) | 4335 (2024 to 6496) | 11.09 (4.89 to 16.62) | -0.30 (-0.49 to -0.12) |
| American Samoa | 2 (1 to 3) | 5.13 (3.63 to 10.00) | 2 (1 to 4) | 4.15 (2.45 to 8.06) | -0.59 (-0.62 to -0.55) |
| Andorra | 4 (3 to 6) | 9.24 (6.48 to 12.64) | 4 (3 to 6) | 3.55 (2.51 to 4.90) | -3.13 (-3.50 to -2.76) |
| Angola | 1120 (597 to 2655) | 10.14 (6.07 to 21.29) | 1628 (976 to 2924) | 6.21 (3.27 to 11.56) | -1.55 (-1.79 to -1.32) |
| Antigua and Barbuda | 3 (2 to 3) | 4.42 (3.93 to 5.05) | 1 (1 to 2) | 1.50 (1.37 to 1.64) | -3.42 (-3.89 to -2.94) |
| Argentina | 6519 (5775 to 7312) | 19.89 (17.60 to 22.29) | 3409 (3183 to 3695) | 7.12 (6.62 to 7.76) | -3.28 (-3.89 to -2.66) |
| Armenia | 256 (222 to 298) | 8.12 (7.03 to 9.55) | 81 (67 to 94) | 2.18 (1.82 to 2.53) | -4.10 (-4.93 to -3.26) |
| Australia | 1322 (1151 to 1672) | 7.82 (6.77 to 9.87) | 1207 (1087 to 1333) | 3.69 (3.36 to 4.03) | -2.36 (-2.73 to -1.98) |
| Austria | 509 (475 to 545) | 5.45 (5.12 to 5.82) | 495 (450 to 542) | 3.47 (3.21 to 3.77) | -1.43 (-1.72 to -1.13) |
| Azerbaijan | 1503 (949 to 2126) | 22.60 (14.60 to 31.48) | 1871 (1078 to 2999) | 17.64 (10.69 to 27.74) | -0.84 (-1.07 to -0.60) |
| Bahamas | 105 (93 to 119) | 44.29 (39.24 to 49.69) | 59 (47 to 75) | 14.75 (11.70 to 18.62) | -3.50 (-3.86 to -3.14) |
| Bahrain | 29 (21 to 56) | 6.60 (4.94 to 14.25) | 48 (24 to 83) | 4.52 (2.26 to 8.02) | -1.23 (-1.68 to -0.79) |
| Bangladesh | 13289 (7185 to 19200) | 13.28 (6.39 to 21.85) | 12634 (7167 to 19514) | 8.66 (4.81 to 13.69) | -1.33 (-1.63 to -1.02) |
| Barbados | 97 (84 to 110) | 37.98 (32.89 to 42.51) | 43 (34 to 54) | 11.88 (9.32 to 15.07) | -3.45 (-3.94 to -2.95) |
| Belarus | 222 (185 to 282) | 2.22 (1.86 to 2.94) | 185 (154 to 220) | 1.63 (1.37 to 1.89) | -1.16 (-1.34 to -0.97) |
| Belgium | 1112 (991 to 1389) | 9.49 (8.41 to 11.94) | 900 (801 to 981) | 5.18 (4.74 to 5.60) | -2.13 (-2.25 to -2.00) |
| Belize | 51 (45 to 58) | 23.42 (21.30 to 26.33) | 22 (20 to 25) | 5.72 (5.03 to 6.45) | -4.42 (-5.08 to -3.76) |
| Benin | 442 (271 to 851) | 7.98 (4.18 to 14.04) | 780 (487 to 1192) | 5.73 (2.98 to 9.04) | -1.05 (-1.21 to -0.89) |
| Bermuda | 34 (30 to 39) | 55.52 (48.52 to 64.73) | 16 (13 to 19) | 15.69 (13.06 to 19.07) | -4.06 (-4.75 to -3.37) |
| Bhutan | 92 (45 to 137) | 14.29 (7.77 to 22.41) | 61 (34 to 97) | 9.27 (5.13 to 14.22) | -1.36 (-1.71 to -1.02) |
| Bolivia (Plurinational State of) | 1591 (718 to 2702) | 19.49 (10.24 to 31.46) | 980 (708 to 1392) | 9.02 (6.57 to 12.82) | -2.45 (-2.49 to -2.40) |
| Bosnia and Herzegovina | 344 (245 to 459) | 7.89 (5.70 to 10.57) | 295 (212 to 381) | 5.91 (4.25 to 7.68) | -0.92 (-1.28 to -0.57) |
| Botswana | 69 (46 to 113) | 6.58 (4.32 to 10.65) | 86 (60 to 114) | 4.04 (2.86 to 5.30) | -1.58 (-1.94 to -1.22) |
| Brazil | 18943 (17689 to 20254) | 14.51 (13.64 to 15.35) | 24078 (22850 to 25249) | 10.45 (9.88 to 11.02) | -1.06 (-1.42 to -0.70) |
| Brunei Darussalam | 54 (28 to 72) | 20.62 (11.28 to 27.62) | 45 (27 to 58) | 11.44 (7.20 to 14.88) | -1.89 (-2.13 to -1.65) |
| Bulgaria | 485 (412 to 565) | 4.89 (4.14 to 5.68) | 286 (217 to 376) | 2.87 (2.19 to 3.74) | -1.66 (-2.55 to -0.76) |
| Burkina Faso | 1142 (637 to 1963) | 11.14 (4.83 to 21.45) | 1992 (1162 to 3181) | 8.98 (4.20 to 14.92) | -0.68 (-0.91 to -0.46) |
| Burundi | 981 (592 to 1971) | 15.60 (8.34 to 32.70) | 916 (414 to 1813) | 7.64 (3.08 to 15.11) | -2.31 (-2.54 to -2.08) |
| Cabo Verde | 33 (16 to 64) | 8.63 (3.53 to 18.25) | 22 (9 to 41) | 4.41 (1.75 to 8.30) | -2.13 (-2.35 to -1.92) |
| Cambodia | 722 (351 to 1695) | 6.65 (3.49 to 14.12) | 777 (500 to 1513) | 4.92 (3.12 to 10.08) | -0.96 (-1.03 to -0.89) |
| Cameroon | 846 (539 to 1528) | 8.38 (4.19 to 15.26) | 1814 (1238 to 2862) | 6.27 (3.78 to 10.57) | -0.92 (-1.04 to -0.81) |
| Canada | 3066 (2872 to 3264) | 11.37 (10.64 to 12.11) | 2171 (1982 to 2337) | 4.63 (4.23 to 4.99) | -2.76 (-3.25 to -2.26) |
| Central African Republic | 324 (177 to 698) | 11.36 (6.34 to 21.84) | 429 (232 to 780) | 8.92 (4.34 to 15.95) | -0.82 (-0.91 to -0.73) |
| Chad | 664 (310 to 1316) | 9.49 (4.12 to 20.39) | 1766 (980 to 3156) | 9.18 (4.46 to 16.36) | -0.05 (-0.19 to 0.09) |
| Chile | 1159 (1084 to 1238) | 9.04 (8.49 to 9.64) | 1112 (1028 to 1201) | 5.45 (4.99 to 5.90) | -1.63 (-1.88 to -1.37) |
| China | 149699 (115904 to 202546) | 16.18 (12.63 to 21.60) | 150941 (99583 to 186503) | 8.95 (6.04 to 11.13) | -1.90 (-2.21 to -1.58) |
| Colombia | 1787 (1488 to 2250) | 5.81 (4.85 to 7.26) | 1645 (1380 to 1949) | 3.29 (2.74 to 3.93) | -1.85 (-2.17 to -1.54) |
| Comoros | 62 (35 to 108) | 11.73 (6.64 to 19.72) | 41 (23 to 81) | 5.92 (3.15 to 11.42) | -2.30 (-3.44 to -1.14) |
| Congo | 173 (114 to 372) | 7.91 (5.31 to 16.16) | 206 (131 to 369) | 4.80 (2.93 to 8.78) | -1.56 (-1.80 to -1.31) |
| Cook Islands | 2 (1 to 3) | 9.83 (7.25 to 17.61) | 1 (1 to 2) | 5.26 (3.14 to 10.93) | -1.86 (-2.07 to -1.66) |
| Costa Rica | 311 (289 to 335) | 12.40 (11.51 to 13.34) | 164 (145 to 183) | 3.31 (2.92 to 3.70) | -4.49 (-5.02 to -3.96) |
| Côte d'Ivoire | 1114 (729 to 1989) | 8.96 (4.77 to 15.04) | 1818 (1114 to 2742) | 6.79 (3.56 to 10.99) | -0.89 (-1.13 to -0.65) |
| Croatia | 191 (169 to 218) | 3.46 (3.07 to 3.92) | 92 (78 to 108) | 1.31 (1.11 to 1.53) | -3.16 (-3.77 to -2.55) |
| Cuba | 484 (445 to 534) | 4.69 (4.31 to 5.19) | 265 (228 to 301) | 1.90 (1.64 to 2.14) | -2.66 (-2.97 to -2.35) |
| Cyprus | 377 (205 to 488) | 52.32 (28.85 to 66.39) | 305 (166 to 379) | 17.45 (9.61 to 21.85) | -3.56 (-4.06 to -3.06) |
| Czechia | 1445 (1216 to 1725) | 12.03 (10.05 to 14.42) | 1899 (1608 to 2170) | 11.11 (9.35 to 12.86) | -0.26 (-0.96 to 0.45) |
| Democratic People's Republic of Korea | 1492 (1014 to 2586) | 8.47 (5.83 to 14.64) | 2285 (1613 to 3818) | 8.04 (5.73 to 13.40) | -0.16 (-0.25 to -0.07) |
| Democratic Republic of the Congo | 3648 (2260 to 6621) | 8.75 (5.31 to 15.90) | 4158 (2013 to 7024) | 6.13 (2.66 to 11.34) | -1.14 (-1.28 to -0.99) |
| Denmark | 474 (438 to 513) | 8.64 (7.98 to 9.36) | 426 (384 to 462) | 4.80 (4.39 to 5.20) | -1.83 (-2.24 to -1.42) |
| Djibouti | 39 (20 to 78) | 9.30 (5.13 to 18.30) | 63 (30 to 119) | 5.68 (2.66 to 10.81) | -1.49 (-1.92 to -1.05) |
| Dominica | 4 (3 to 6) | 6.07 (3.90 to 8.16) | 2 (2 to 4) | 3.46 (2.49 to 5.66) | -1.80 (-1.94 to -1.65) |
| Dominican Republic | 842 (528 to 1156) | 9.71 (7.02 to 13.14) | 456 (330 to 832) | 4.26 (3.05 to 7.94) | -2.66 (-3.11 to -2.22) |
| Ecuador | 954 (802 to 1234) | 9.56 (7.77 to 11.99) | 957 (812 to 1116) | 5.80 (4.94 to 6.74) | -1.71 (-2.07 to -1.35) |
| Egypt | 58615 (23940 to 86262) | 74.21 (37.61 to 105.38) | 17676 (13635 to 23381) | 16.51 (12.82 to 21.87) | -4.77 (-5.10 to -4.44) |
| El Salvador | 677 (422 to 882) | 11.91 (7.96 to 15.04) | 263 (190 to 441) | 4.18 (3.02 to 6.98) | -3.30 (-3.46 to -3.15) |
| Equatorial Guinea | 41 (22 to 83) | 9.48 (5.49 to 17.84) | 43 (24 to 75) | 3.83 (2.13 to 6.65) | -2.88 (-3.19 to -2.58) |
| Eritrea | 460 (256 to 967) | 13.09 (7.07 to 27.04) | 470 (239 to 931) | 8.01 (3.80 to 15.23) | -1.67 (-1.74 to -1.61) |
| Estonia | 19 (16 to 22) | 1.17 (1.02 to 1.36) | 24 (21 to 28) | 1.12 (0.96 to 1.29) | -0.27 (-2.41 to 1.92) |
| Eswatini | 44 (31 to 68) | 6.76 (4.70 to 10.53) | 53 (35 to 73) | 5.30 (3.57 to 7.29) | -0.76 (-0.95 to -0.57) |
| Ethiopia | 5663 (3177 to 11304) | 10.79 (5.32 to 20.84) | 5398 (2500 to 11569) | 5.53 (2.30 to 11.22) | -2.14 (-2.30 to -1.98) |
| Fiji | 44 (28 to 78) | 7.00 (4.49 to 13.49) | 44 (25 to 83) | 5.17 (2.93 to 9.99) | -0.97 (-1.10 to -0.83) |
| Finland | 173 (152 to 195) | 3.12 (2.74 to 3.50) | 178 (160 to 198) | 2.13 (1.95 to 2.35) | -1.20 (-1.56 to -0.85) |
| France | 5949 (4979 to 7308) | 9.04 (7.70 to 11.20) | 6113 (5521 to 6689) | 6.05 (5.60 to 6.55) | -1.27 (-1.63 to -0.90) |
| Gabon | 61 (39 to 108) | 6.99 (4.07 to 13.04) | 60 (34 to 104) | 4.14 (2.32 to 7.11) | -1.67 (-1.92 to -1.41) |
| Gambia | 79 (46 to 145) | 8.36 (3.72 to 15.32) | 134 (70 to 241) | 6.52 (3.06 to 11.69) | -0.79 (-1.73 to 0.16) |
| Georgia | 756 (585 to 946) | 13.22 (10.31 to 16.55) | 1318 (1039 to 1665) | 27.83 (22.18 to 34.82) | 2.31 (1.71 to 2.92) |
| Germany | 8412 (7108 to 9616) | 9.48 (8.04 to 10.81) | 9441 (8306 to 10352) | 6.42 (5.86 to 6.91) | -1.35 (-1.91 to -0.79) |
| Ghana | 1066 (720 to 2056) | 8.01 (4.51 to 14.84) | 1256 (806 to 2062) | 4.49 (2.65 to 7.35) | -1.85 (-1.97 to -1.74) |
| Greece | 860 (809 to 912) | 7.86 (7.37 to 8.31) | 1495 (1340 to 1645) | 8.35 (7.67 to 9.13) | 0.19 (-0.33 to 0.71) |
| Greenland | 16 (6 to 23) | 31.58 (13.61 to 44.07) | 4 (3 to 6) | 7.06 (5.05 to 10.16) | -4.72 (-5.00 to -4.44) |
| Grenada | 28 (24 to 33) | 32.89 (28.29 to 38.70) | 9 (8 to 11) | 8.80 (7.66 to 10.03) | -4.16 (-4.50 to -3.82) |
| Guam | 5 (4 to 10) | 4.52 (3.26 to 8.95) | 4 (2 to 8) | 2.53 (1.37 to 4.91) | -1.86 (-2.24 to -1.47) |
| Guatemala | 863 (757 to 1064) | 9.37 (8.31 to 11.15) | 324 (275 to 386) | 2.36 (2.02 to 2.80) | -4.50 (-4.88 to -4.13) |
| Guinea | 773 (421 to 1603) | 10.26 (5.42 to 21.05) | 1090 (642 to 1803) | 8.02 (3.99 to 13.76) | -0.78 (-0.97 to -0.60) |
| Guinea-Bissau | 126 (74 to 252) | 12.24 (6.65 to 22.93) | 153 (85 to 261) | 8.52 (4.19 to 14.99) | -1.16 (-1.34 to -0.98) |
| Guyana | 79 (68 to 92) | 8.66 (7.69 to 9.79) | 119 (91 to 157) | 16.06 (12.29 to 21.11) | 2.15 (1.51 to 2.79) |
| Haiti | 3923 (1303 to 7297) | 45.54 (17.74 to 78.68) | 3454 (1446 to 6073) | 25.17 (11.87 to 42.29) | -1.85 (-2.01 to -1.70) |
| Honduras | 338 (215 to 467) | 8.15 (5.15 to 11.98) | 409 (283 to 617) | 5.30 (3.67 to 8.02) | -1.34 (-1.53 to -1.16) |
| Hungary | 1268 (1113 to 1451) | 10.13 (8.93 to 11.56) | 725 (590 to 908) | 4.66 (3.75 to 5.82) | -2.51 (-3.20 to -1.81) |
| Iceland | 14 (13 to 16) | 5.55 (5.16 to 5.99) | 17 (15 to 19) | 3.79 (3.36 to 4.27) | -1.18 (-1.56 to -0.81) |
| India | 99575 (57534 to 134279) | 11.73 (7.49 to 16.30) | 94737 (69017 to 131902) | 7.75 (5.52 to 11.36) | -1.32 (-1.47 to -1.16) |
| Indonesia | 10341 (5841 to 24422) | 6.00 (3.43 to 13.39) | 11947 (7868 to 24908) | 4.61 (3.03 to 10.44) | -0.83 (-0.94 to -0.72) |
| Iran (Islamic Republic of) | 28135 (17155 to 38911) | 50.93 (32.60 to 63.56) | 12801 (9989 to 14672) | 16.26 (12.86 to 18.66) | -3.66 (-3.93 to -3.40) |
| Iraq | 2476 (1291 to 3562) | 11.13 (6.20 to 16.49) | 1822 (993 to 3158) | 5.20 (2.84 to 10.09) | -2.46 (-2.65 to -2.28) |
| Ireland | 168 (158 to 180) | 4.59 (4.34 to 4.90) | 190 (172 to 209) | 3.02 (2.74 to 3.33) | -1.37 (-1.85 to -0.89) |
| Israel | 797 (706 to 1034) | 16.23 (14.40 to 21.01) | 796 (719 to 869) | 7.18 (6.52 to 7.81) | -2.65 (-2.84 to -2.46) |
| Italy | 4731 (4493 to 4968) | 7.16 (6.83 to 7.49) | 2779 (2504 to 3033) | 2.78 (2.58 to 2.99) | -3.09 (-3.46 to -2.72) |
| Jamaica | 230 (200 to 265) | 9.67 (8.40 to 11.00) | 78 (60 to 100) | 2.81 (2.16 to 3.62) | -3.86 (-4.42 to -3.30) |
| Japan | 15634 (15119 to 16190) | 14.12 (13.66 to 14.60) | 18490 (16064 to 20370) | 10.72 (10.03 to 11.46) | -0.89 (-1.43 to -0.35) |
| Jordan | 122 (67 to 444) | 2.77 (1.49 to 10.88) | 253 (124 to 686) | 2.45 (1.20 to 6.89) | -0.39 (-0.74 to -0.04) |
| Kazakhstan | 264 (217 to 310) | 1.74 (1.42 to 2.06) | 268 (220 to 316) | 1.49 (1.23 to 1.74) | -0.58 (-1.56 to 0.42) |
| Kenya | 1164 (748 to 2248) | 5.22 (2.64 to 10.69) | 1655 (820 to 3204) | 4.05 (1.93 to 7.82) | -0.82 (-0.91 to -0.73) |
| Kiribati | 7 (4 to 13) | 9.82 (5.55 to 19.60) | 8 (5 to 16) | 8.09 (4.85 to 15.83) | -0.63 (-0.69 to -0.57) |
| Kuwait | 68 (61 to 78) | 4.17 (3.73 to 4.70) | 207 (174 to 243) | 6.51 (5.41 to 7.73) | 1.81 (-0.86 to 4.56) |
| Kyrgyzstan | 138 (116 to 164) | 3.48 (2.92 to 4.17) | 105 (86 to 131) | 1.76 (1.46 to 2.16) | -2.21 (-3.14 to -1.28) |
| Lao People's Democratic Republic | 479 (154 to 1320) | 9.96 (3.82 to 24.56) | 479 (256 to 980) | 6.65 (3.69 to 13.98) | -1.29 (-1.41 to -1.17) |
| Latvia | 33 (28 to 39) | 1.17 (1.02 to 1.36) | 89 (77 to 102) | 2.89 (2.51 to 3.30) | 2.57 (1.22 to 3.94) |
| Lebanon | 948 (529 to 1451) | 31.97 (18.96 to 48.19) | 617 (485 to 814) | 11.26 (8.86 to 14.49) | -3.38 (-3.66 to -3.10) |
| Lesotho | 62 (39 to 105) | 4.93 (3.06 to 8.21) | 87 (59 to 116) | 5.40 (3.74 to 7.19) | 0.29 (0.13 to 0.46) |
| Liberia | 354 (202 to 711) | 11.46 (6.85 to 20.21) | 339 (173 to 575) | 6.94 (3.10 to 12.33) | -1.76 (-2.19 to -1.32) |
| Libya | 1146 (647 to 2165) | 20.28 (11.95 to 37.85) | 1004 (437 to 1782) | 20.43 (8.75 to 36.65) | 0.01 (-0.53 to 0.56) |
| Lithuania | 63 (55 to 72) | 1.71 (1.51 to 1.95) | 100 (87 to 113) | 2.30 (2.04 to 2.58) | 0.70 (-1.15 to 2.57) |
| Luxembourg | 36 (33 to 38) | 8.29 (7.72 to 8.79) | 46 (41 to 51) | 5.06 (4.54 to 5.64) | -1.63 (-2.06 to -1.21) |
| Madagascar | 2391 (1395 to 5242) | 18.04 (10.75 to 35.43) | 3454 (1961 to 6745) | 12.80 (6.85 to 24.47) | -1.11 (-1.17 to -1.05) |
| Malawi | 1165 (640 to 2319) | 9.49 (5.69 to 17.02) | 1046 (546 to 1827) | 6.17 (3.08 to 10.65) | -1.32 (-1.60 to -1.04) |
| Malaysia | 511 (367 to 1189) | 3.22 (2.26 to 8.39) | 757 (519 to 1871) | 2.48 (1.66 to 6.63) | -0.90 (-1.65 to -0.14) |
| Maldives | 55 (33 to 79) | 22.06 (14.86 to 29.92) | 34 (27 to 50) | 7.48 (5.98 to 10.28) | -3.53 (-3.70 to -3.37) |
| Mali | 879 (465 to 1797) | 9.00 (4.50 to 19.24) | 1667 (918 to 2938) | 6.95 (3.08 to 13.92) | -0.81 (-1.07 to -0.56) |
| Malta | 12 (11 to 13) | 3.19 (2.93 to 3.48) | 18 (16 to 20) | 2.90 (2.55 to 3.27) | -0.28 (-0.71 to 0.14) |
| Marshall Islands | 3 (2 to 6) | 10.40 (6.53 to 19.46) | 4 (2 to 7) | 8.04 (4.72 to 15.81) | -0.82 (-0.89 to -0.76) |
| Mauritania | 136 (86 to 268) | 6.92 (3.84 to 14.05) | 177 (101 to 300) | 4.73 (2.40 to 8.52) | -1.26 (-1.40 to -1.13) |
| Mauritius | 110 (102 to 119) | 10.91 (10.11 to 11.82) | 398 (351 to 434) | 28.96 (25.61 to 31.57) | 3.69 (1.76 to 5.65) |
| Mexico | 4819 (4095 to 5871) | 5.59 (5.00 to 6.65) | 3759 (3349 to 4226) | 3.11 (2.78 to 3.53) | -1.89 (-2.39 to -1.40) |
| Micronesia (Federated States of) | 9 (5 to 16) | 10.85 (6.76 to 19.90) | 7 (4 to 13) | 7.48 (4.69 to 15.03) | -1.20 (-1.24 to -1.16) |
| Monaco | 3 (2 to 3) | 5.54 (4.21 to 7.60) | 3 (2 to 4) | 4.46 (3.07 to 6.12) | -0.65 (-0.79 to -0.51) |
| Mongolia | 801 (407 to 1220) | 50.71 (26.29 to 79.60) | 1230 (721 to 1614) | 43.92 (25.60 to 56.54) | -0.49 (-0.85 to -0.12) |
| Montenegro | 10 (5 to 41) | 1.54 (0.85 to 6.81) | 8 (5 to 38) | 1.05 (0.60 to 4.82) | -1.11 (-1.50 to -0.72) |
| Morocco | 4665 (2537 to 7927) | 15.60 (8.74 to 26.69) | 4559 (2226 to 6953) | 13.46 (6.51 to 20.47) | -0.48 (-0.66 to -0.30) |
| Mozambique | 1573 (837 to 2789) | 10.78 (5.94 to 20.22) | 2157 (1087 to 3608) | 8.59 (3.81 to 15.15) | -0.75 (-1.00 to -0.50) |
| Myanmar | 4138 (1916 to 10102) | 10.14 (5.16 to 22.81) | 3636 (2377 to 7104) | 6.76 (4.43 to 13.78) | -1.30 (-1.41 to -1.20) |
| Namibia | 66 (47 to 98) | 5.88 (4.21 to 8.48) | 99 (67 to 141) | 4.86 (3.36 to 6.76) | -0.60 (-0.73 to -0.47) |
| Nauru | 1 (1 to 2) | 10.68 (6.85 to 20.99) | 1 (1 to 2) | 10.30 (6.33 to 22.74) | -0.11 (-0.22 to -0.01) |
| Nepal | 3424 (1746 to 5125) | 14.75 (8.37 to 23.03) | 2557 (1465 to 3967) | 9.42 (5.15 to 15.02) | -1.43 (-1.55 to -1.32) |
| Netherlands | 873 (819 to 938) | 5.38 (5.07 to 5.74) | 1004 (915 to 1106) | 3.91 (3.61 to 4.26) | -1.05 (-1.53 to -0.57) |
| New Zealand | 192 (178 to 206) | 5.33 (4.94 to 5.71) | 226 (207 to 249) | 3.55 (3.26 to 3.89) | -1.30 (-2.40 to -0.19) |
| Nicaragua | 279 (143 to 397) | 6.13 (3.44 to 9.41) | 102 (69 to 302) | 1.76 (1.18 to 5.35) | -3.97 (-4.13 to -3.81) |
| Niger | 955 (461 to 1825) | 10.39 (4.50 to 20.95) | 1835 (768 to 3526) | 7.69 (2.46 to 15.66) | -0.96 (-1.20 to -0.72) |
| Nigeria | 9447 (6712 to 19379) | 9.36 (5.30 to 16.75) | 15081 (10537 to 21578) | 6.04 (4.07 to 8.84) | -1.42 (-1.50 to -1.33) |
| Niue | 0 (0 to 0) | 8.31 (5.42 to 16.73) | 0 (0 to 0) | 11.14 (7.04 to 23.27) | 1.12 (0.56 to 1.68) |
| North Macedonia | 89 (64 to 133) | 4.72 (3.35 to 7.28) | 90 (47 to 153) | 3.18 (1.68 to 5.83) | -1.17 (-1.77 to -0.58) |
| Northern Mariana Islands | 1 (1 to 3) | 3.90 (2.39 to 7.66) | 2 (1 to 3) | 3.36 (2.06 to 6.99) | -0.49 (-0.63 to -0.34) |
| Norway | 287 (274 to 301) | 7.21 (6.89 to 7.52) | 133 (121 to 148) | 1.92 (1.77 to 2.11) | -4.55 (-6.55 to -2.50) |
| Oman | 87 (46 to 226) | 4.18 (2.22 to 12.07) | 134 (51 to 241) | 3.90 (1.49 to 7.22) | -0.22 (-0.67 to 0.23) |
| Pakistan | 19706 (10809 to 28216) | 14.41 (8.92 to 20.63) | 26574 (16921 to 38115) | 11.96 (7.86 to 18.26) | -0.61 (-0.82 to -0.40) |
| Palau | 1 (0 to 2) | 6.30 (3.58 to 12.73) | 1 (0 to 2) | 4.37 (2.21 to 9.17) | -1.18 (-1.26 to -1.09) |
| Palestine | 293 (131 to 482) | 10.15 (4.91 to 19.39) | 212 (91 to 500) | 4.66 (1.98 to 11.88) | -2.50 (-2.73 to -2.27) |
| Panama | 132 (114 to 150) | 6.27 (5.44 to 7.04) | 89 (74 to 104) | 2.08 (1.73 to 2.46) | -3.45 (-3.77 to -3.13) |
| Papua New Guinea | 533 (313 to 852) | 12.35 (7.61 to 19.88) | 1243 (832 to 2020) | 11.31 (7.43 to 19.45) | -0.27 (-0.52 to -0.02) |
| Paraguay | 122 (87 to 254) | 3.18 (2.24 to 7.27) | 157 (103 to 384) | 2.42 (1.58 to 6.14) | -0.87 (-1.33 to -0.41) |
| Peru | 2504 (1458 to 3769) | 10.57 (6.89 to 14.85) | 1607 (1197 to 2184) | 4.65 (3.45 to 6.29) | -2.63 (-2.89 to -2.37) |
| Philippines | 3227 (2173 to 5819) | 4.93 (3.35 to 10.27) | 3531 (2584 to 7427) | 3.28 (2.37 to 7.50) | -1.33 (-1.72 to -0.94) |
| Poland | 2435 (2143 to 2770) | 6.04 (5.34 to 6.87) | 2112 (1912 to 2297) | 3.95 (3.58 to 4.29) | -1.51 (-1.90 to -1.12) |
| Portugal | 886 (833 to 941) | 8.98 (8.47 to 9.56) | 1022 (917 to 1117) | 5.82 (5.35 to 6.28) | -1.48 (-2.11 to -0.85) |
| Puerto Rico | 832 (773 to 899) | 23.73 (22.03 to 25.57) | 158 (132 to 185) | 3.98 (3.33 to 4.67) | -5.50 (-6.31 to -4.69) |
| Qatar | 19 (13 to 37) | 5.34 (3.73 to 10.72) | 69 (37 to 102) | 3.75 (1.96 to 5.57) | -1.21 (-1.95 to -0.46) |
| Republic of Korea | 2475 (1692 to 3462) | 6.52 (4.49 to 9.50) | 1246 (842 to 2906) | 2.40 (1.67 to 4.75) | -3.08 (-3.42 to -2.74) |
| Republic of Moldova | 17 (13 to 23) | 0.39 (0.29 to 0.52) | 22 (18 to 29) | 0.54 (0.44 to 0.68) | 1.07 (0.74 to 1.40) |
| Romania | 3536 (2925 to 4196) | 14.19 (11.85 to 16.68) | 3949 (3384 to 4491) | 13.92 (12.01 to 15.83) | -0.12 (-0.47 to 0.22) |
| Russian Federation | 18577 (16747 to 21771) | 12.67 (11.52 to 14.59) | 6389 (5934 to 6897) | 3.62 (3.38 to 3.89) | -3.97 (-4.89 to -3.03) |
| Rwanda | 1150 (686 to 2622) | 14.89 (8.47 to 30.80) | 725 (356 to 1529) | 6.17 (2.87 to 13.12) | -2.75 (-3.27 to -2.22) |
| Saint Kitts and Nevis | 3 (3 to 4) | 8.12 (7.10 to 11.40) | 1 (1 to 2) | 2.32 (1.90 to 2.81) | -3.84 (-4.40 to -3.27) |
| Saint Lucia | 35 (30 to 39) | 28.45 (25.20 to 31.93) | 16 (13 to 19) | 8.36 (6.78 to 10.22) | -3.90 (-4.30 to -3.49) |
| Saint Vincent and the Grenadines | 5 (4 to 5) | 4.30 (3.81 to 4.87) | 3 (2 to 3) | 2.45 (2.08 to 2.88) | -1.76 (-2.33 to -1.19) |
| Samoa | 12 (8 to 20) | 8.64 (5.80 to 15.51) | 12 (8 to 23) | 6.56 (4.40 to 12.94) | -0.88 (-0.94 to -0.81) |
| San Marino | 1 (1 to 2) | 3.20 (2.46 to 5.88) | 1 (1 to 2) | 1.61 (1.07 to 3.13) | -2.29 (-2.51 to -2.06) |
| Sao Tome and Principe | 9 (5 to 18) | 6.57 (3.75 to 12.34) | 8 (3 to 15) | 4.36 (1.85 to 8.40) | -1.30 (-1.55 to -1.06) |
| Saudi Arabia | 952 (573 to 1681) | 5.65 (3.56 to 11.82) | 872 (465 to 1518) | 2.62 (1.45 to 5.39) | -2.50 (-2.88 to -2.12) |
| Senegal | 820 (535 to 1575) | 9.81 (5.09 to 17.60) | 904 (433 to 1528) | 6.40 (2.74 to 11.21) | -1.26 (-1.55 to -0.98) |
| Serbia | 369 (262 to 789) | 3.76 (2.64 to 8.71) | 337 (215 to 618) | 2.43 (1.58 to 4.62) | -1.37 (-1.56 to -1.17) |
| Seychelles | 3 (2 to 6) | 4.26 (2.88 to 9.42) | 3 (2 to 7) | 2.86 (1.75 to 6.44) | -0.95 (-1.61 to -0.28) |
| Sierra Leone | 644 (353 to 1346) | 11.59 (6.90 to 19.85) | 774 (457 to 1268) | 8.24 (4.47 to 13.62) | -1.12 (-1.56 to -0.69) |
| Singapore | 311 (289 to 335) | 11.63 (10.81 to 12.49) | 207 (187 to 228) | 3.83 (3.44 to 4.27) | -3.57 (-4.45 to -2.67) |
| Slovakia | 190 (129 to 326) | 3.40 (2.31 to 6.14) | 154 (100 to 268) | 2.04 (1.32 to 3.61) | -1.68 (-1.91 to -1.46) |
| Slovenia | 30 (26 to 34) | 1.29 (1.13 to 1.48) | 38 (33 to 44) | 1.03 (0.88 to 1.21) | -0.86 (-1.40 to -0.31) |
| Solomon Islands | 21 (13 to 32) | 7.65 (4.64 to 12.89) | 34 (23 to 59) | 6.31 (4.18 to 11.93) | -0.61 (-0.90 to -0.32) |
| Somalia | 1328 (604 to 2412) | 15.22 (5.96 to 29.72) | 2163 (667 to 4513) | 10.58 (2.79 to 22.97) | -1.15 (-1.35 to -0.95) |
| South Africa | 1545 (1220 to 1881) | 4.50 (3.66 to 5.65) | 1932 (1491 to 2297) | 3.58 (2.77 to 4.23) | -0.74 (-1.09 to -0.38) |
| South Sudan | 974 (440 to 1977) | 13.88 (7.57 to 26.60) | 1106 (628 to 2080) | 10.34 (5.26 to 18.60) | -0.93 (-1.24 to -0.62) |
| Spain | 3743 (3526 to 3949) | 9.18 (8.66 to 9.70) | 4292 (3845 to 4655) | 6.00 (5.52 to 6.41) | -1.42 (-1.78 to -1.06) |
| Sri Lanka | 2711 (1853 to 4476) | 17.23 (12.16 to 28.77) | 3523 (2180 to 5135) | 15.14 (9.48 to 21.88) | -0.29 (-0.56 to -0.02) |
| Sudan | 5213 (2387 to 10407) | 18.24 (9.41 to 32.75) | 7551 (3740 to 11346) | 17.35 (8.60 to 25.58) | -0.14 (-0.26 to -0.01) |
| Suriname | 143 (78 to 178) | 36.76 (21.37 to 45.17) | 79 (58 to 109) | 14.26 (10.62 to 19.62) | -3.02 (-3.40 to -2.64) |
| Sweden | 578 (541 to 617) | 6.08 (5.70 to 6.46) | 604 (540 to 678) | 4.14 (3.73 to 4.63) | -1.21 (-1.85 to -0.56) |
| Switzerland | 927 (811 to 1136) | 12.09 (10.29 to 14.95) | 715 (634 to 794) | 5.12 (4.64 to 5.63) | -2.72 (-2.97 to -2.47) |
| Syrian Arab Republic | 732 (373 to 2390) | 4.54 (2.50 to 16.61) | 284 (133 to 1390) | 2.32 (1.05 to 11.57) | -2.17 (-2.49 to -1.85) |
| Taiwan (Province of China) | 405 (382 to 432) | 2.40 (2.28 to 2.55) | 1515 (1353 to 1649) | 4.19 (3.77 to 4.54) | 1.69 (0.75 to 2.64) |
| Tajikistan | 1916 (1193 to 2736) | 39.71 (24.24 to 53.52) | 2491 (1593 to 3452) | 27.43 (17.75 to 37.43) | -1.23 (-1.38 to -1.08) |
| Thailand | 2105 (1431 to 4590) | 4.14 (2.75 to 10.59) | 2149 (1425 to 6094) | 3.09 (2.13 to 7.50) | -0.99 (-1.33 to -0.66) |
| Timor-Leste | 76 (33 to 194) | 8.01 (4.14 to 17.11) | 88 (52 to 174) | 6.29 (3.77 to 13.06) | -0.81 (-1.18 to -0.44) |
| Togo | 305 (195 to 570) | 8.38 (4.43 to 15.55) | 460 (237 to 734) | 6.36 (3.03 to 10.38) | -0.88 (-1.10 to -0.66) |
| Tokelau | 0 (0 to 0) | 9.27 (5.82 to 17.72) | 0 (0 to 0) | 13.26 (8.10 to 31.56) | 1.46 (0.55 to 2.38) |
| Tonga | 5 (4 to 9) | 6.11 (4.36 to 11.58) | 4 (3 to 9) | 4.46 (2.97 to 9.33) | -1.02 (-1.13 to -0.91) |
| Trinidad and Tobago | 233 (209 to 260) | 20.94 (18.88 to 23.27) | 110 (85 to 142) | 7.72 (5.91 to 9.91) | -3.00 (-3.51 to -2.49) |
| Tunisia | 901 (574 to 1529) | 10.56 (6.84 to 18.90) | 1064 (438 to 1721) | 9.23 (3.88 to 14.80) | -0.45 (-0.60 to -0.30) |
| Türkiye | 31949 (16439 to 49530) | 53.91 (30.72 to 77.55) | 12652 (9856 to 15875) | 16.56 (12.93 to 20.56) | -3.76 (-3.89 to -3.63) |
| Turkmenistan | 196 (163 to 245) | 6.31 (5.00 to 7.84) | 380 (290 to 499) | 7.85 (5.96 to 10.27) | 0.74 (0.35 to 1.13) |
| Tuvalu | 1 (1 to 2) | 12.52 (7.35 to 23.75) | 1 (1 to 2) | 7.30 (4.76 to 14.38) | -1.73 (-1.82 to -1.64) |
| Uganda | 2518 (1509 to 4482) | 11.72 (5.79 to 22.80) | 2531 (1529 to 4433) | 5.98 (3.20 to 10.36) | -2.16 (-2.55 to -1.78) |
| Ukraine | 1697 (1533 to 1901) | 2.99 (2.69 to 3.39) | 1547 (1172 to 1956) | 3.05 (2.37 to 3.83) | 0.05 (-1.52 to 1.66) |
| United Arab Emirates | 418 (232 to 632) | 26.79 (14.50 to 38.89) | 434 (266 to 541) | 10.31 (5.63 to 13.04) | -3.06 (-4.19 to -1.92) |
| United Kingdom | 3646 (3234 to 4483) | 6.44 (5.57 to 7.93) | 3037 (2872 to 3181) | 3.73 (3.54 to 3.92) | -1.56 (-1.74 to -1.38) |
| United Republic of Tanzania | 2892 (1921 to 5624) | 10.23 (5.60 to 18.80) | 3514 (2012 to 6433) | 6.27 (3.28 to 11.36) | -1.57 (-1.77 to -1.36) |
| United States of America | 27123 (24675 to 29590) | 10.00 (9.20 to 10.94) | 36198 (33004 to 38648) | 8.08 (7.49 to 8.60) | -0.69 (-0.92 to -0.46) |
| United States Virgin Islands | 18 (10 to 22) | 16.97 (9.78 to 21.31) | 6 (4 to 9) | 6.40 (4.53 to 8.82) | -3.01 (-3.25 to -2.77) |
| Uruguay | 305 (273 to 337) | 9.71 (8.69 to 10.74) | 218 (200 to 238) | 5.44 (4.97 to 5.99) | -1.97 (-2.26 to -1.68) |
| Uzbekistan | 3241 (2716 to 3827) | 17.14 (14.13 to 20.56) | 3874 (3133 to 4699) | 12.20 (9.83 to 14.82) | -1.13 (-1.85 to -0.40) |
| Vanuatu | 13 (8 to 21) | 9.35 (5.88 to 16.81) | 22 (14 to 41) | 7.80 (5.12 to 15.18) | -0.55 (-0.88 to -0.22) |
| Venezuela (Bolivarian Republic of) | 759 (657 to 866) | 4.25 (3.69 to 4.81) | 492 (391 to 609) | 1.83 (1.46 to 2.27) | -2.53 (-3.70 to -1.35) |
| Viet Nam | 3268 (2110 to 6754) | 5.39 (3.27 to 12.30) | 3747 (2123 to 8856) | 3.80 (2.13 to 9.56) | -1.11 (-1.16 to -1.07) |
| Yemen | 3171 (1563 to 5313) | 16.75 (8.15 to 27.97) | 5975 (2917 to 8721) | 19.30 (9.07 to 28.92) | 0.46 (0.20 to 0.72) |
| Zambia | 771 (450 to 1777) | 9.20 (5.51 to 18.24) | 1343 (567 to 2581) | 8.64 (3.22 to 17.19) | -0.18 (-0.34 to -0.02) |
| Zimbabwe | 402 (207 to 678) | 5.22 (2.65 to 8.94) | 955 (595 to 1432) | 7.16 (4.65 to 10.44) | 1.11 (0.65 to 1.56) |

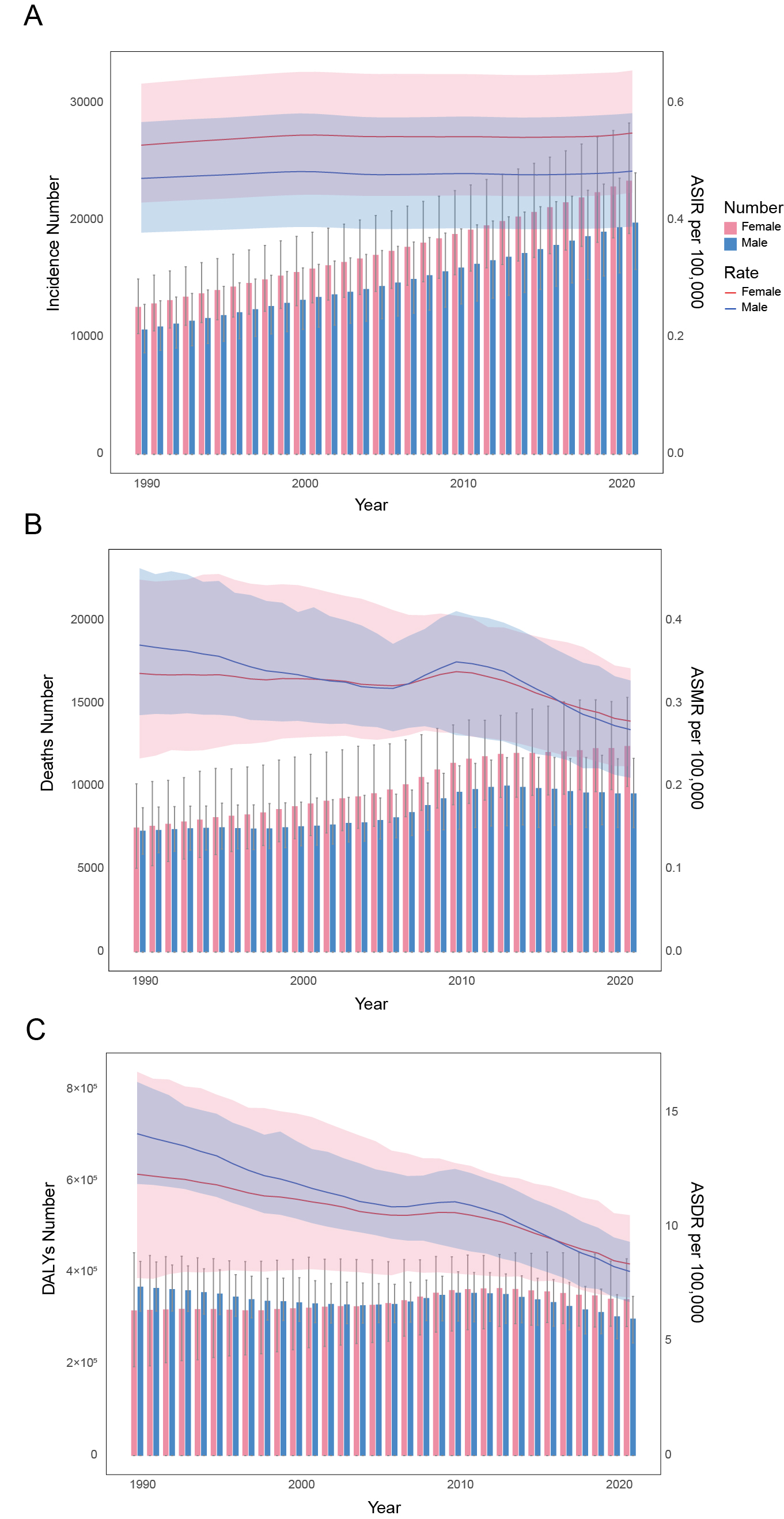

**Supplemental Figure 1. Temporal trends in the all-age cases and age-standardized incidence, mortality, and DALY rates of PAH by sex from 1990 to 2021.** (A) Incidence number and ASIR. (B) Mortality number and ASMR. (C) DALYs number and ASDR. PAH, pulmonary arterial hypertension; ASIR, age-standardized incidence rate; ASMR, age-standardized mortality rate; ASDR, age-standardized disability-adjusted life-year rate.

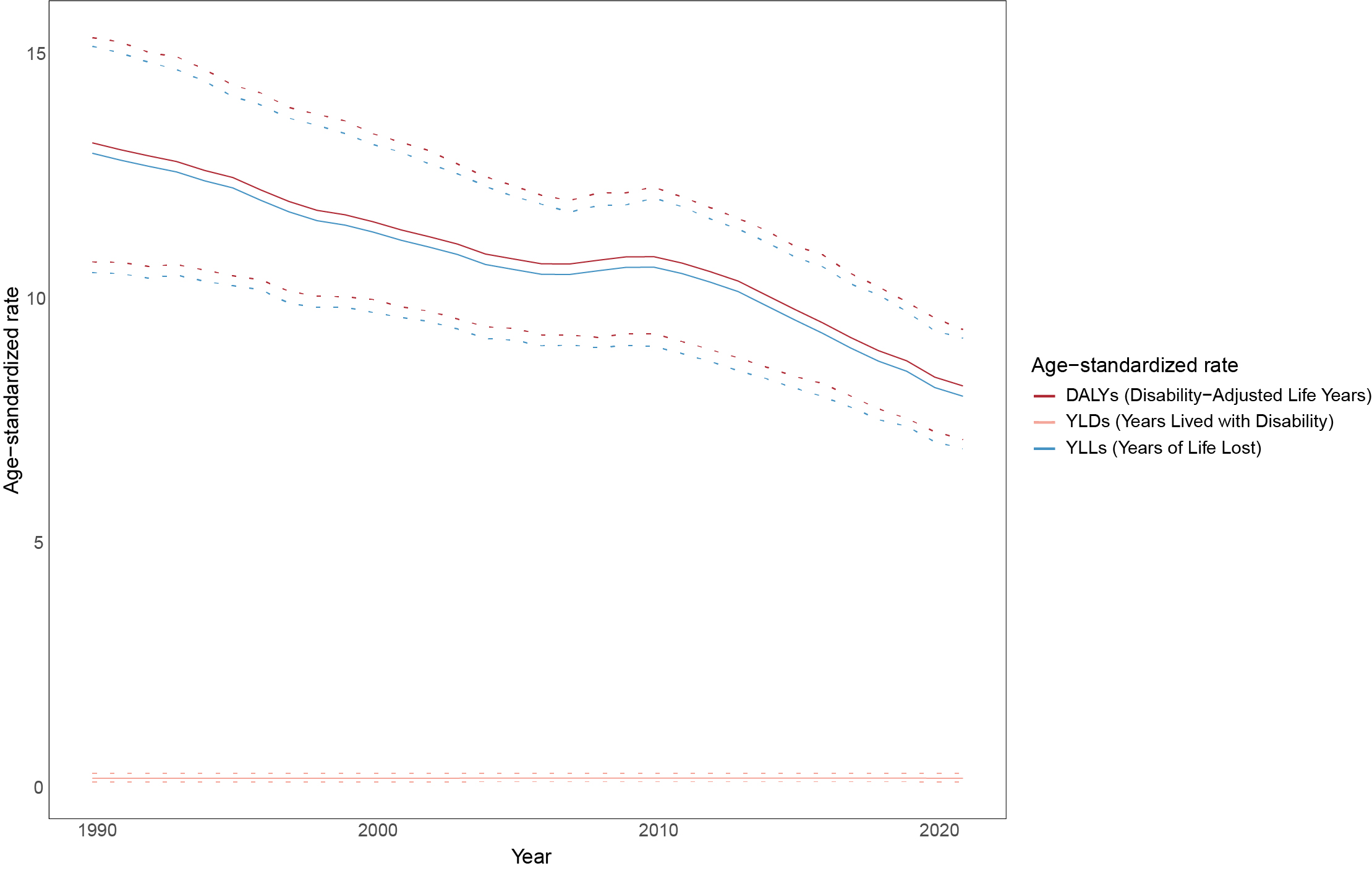

**Supplemental Figure 2. Age-standardized rate of DALY, YLL and YLD of PAH from 1990 to 2021.** The dotted lines indicate 95% upper and lower uncertainty intervals.

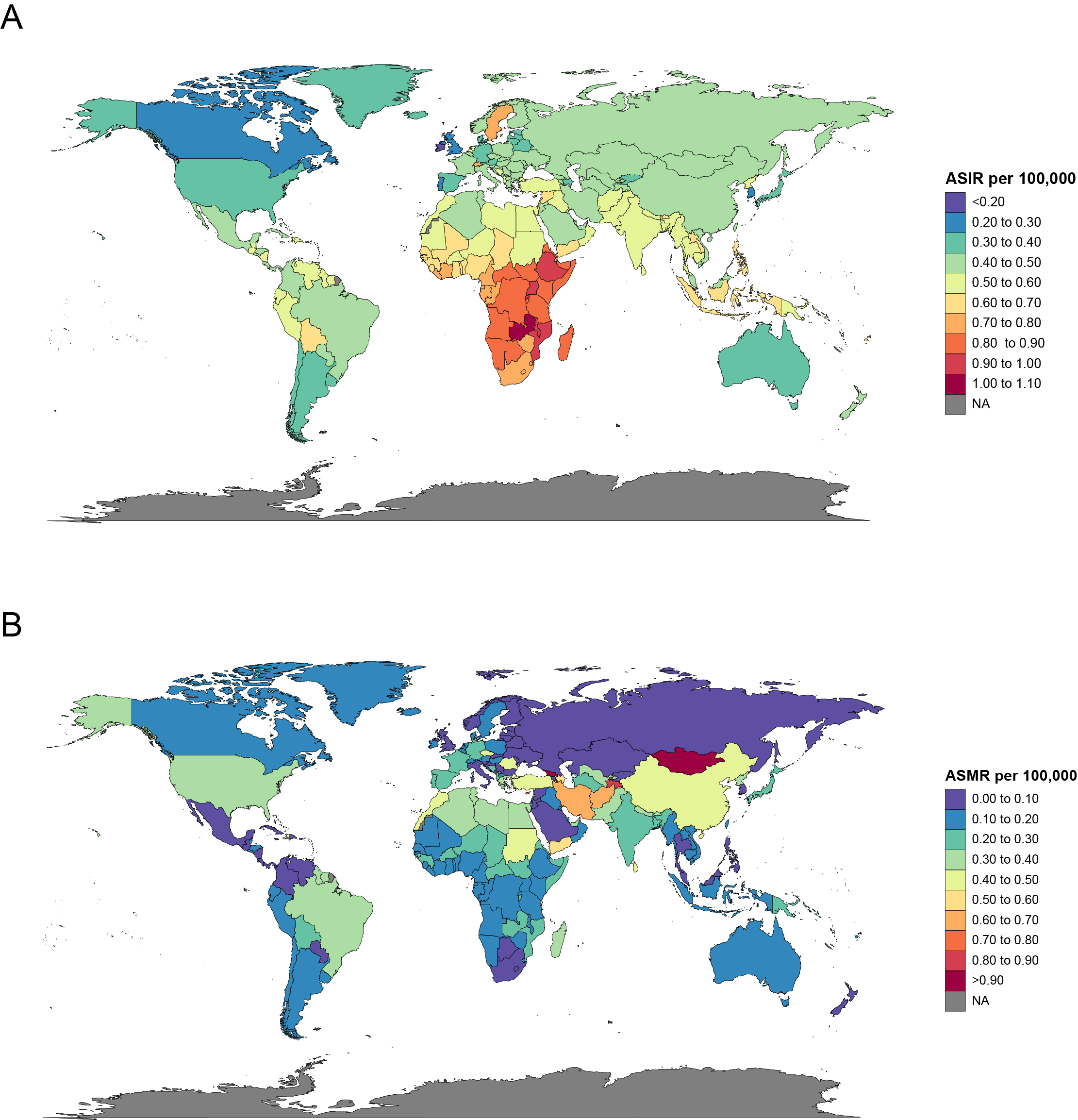

**Supplemental Figure 3. Age-standardized incidence and mortality rates of PAH in 204 countries and territories in 2021.** (A) Disease burden of ASIRs. (B) Disease burden of ASMRs.

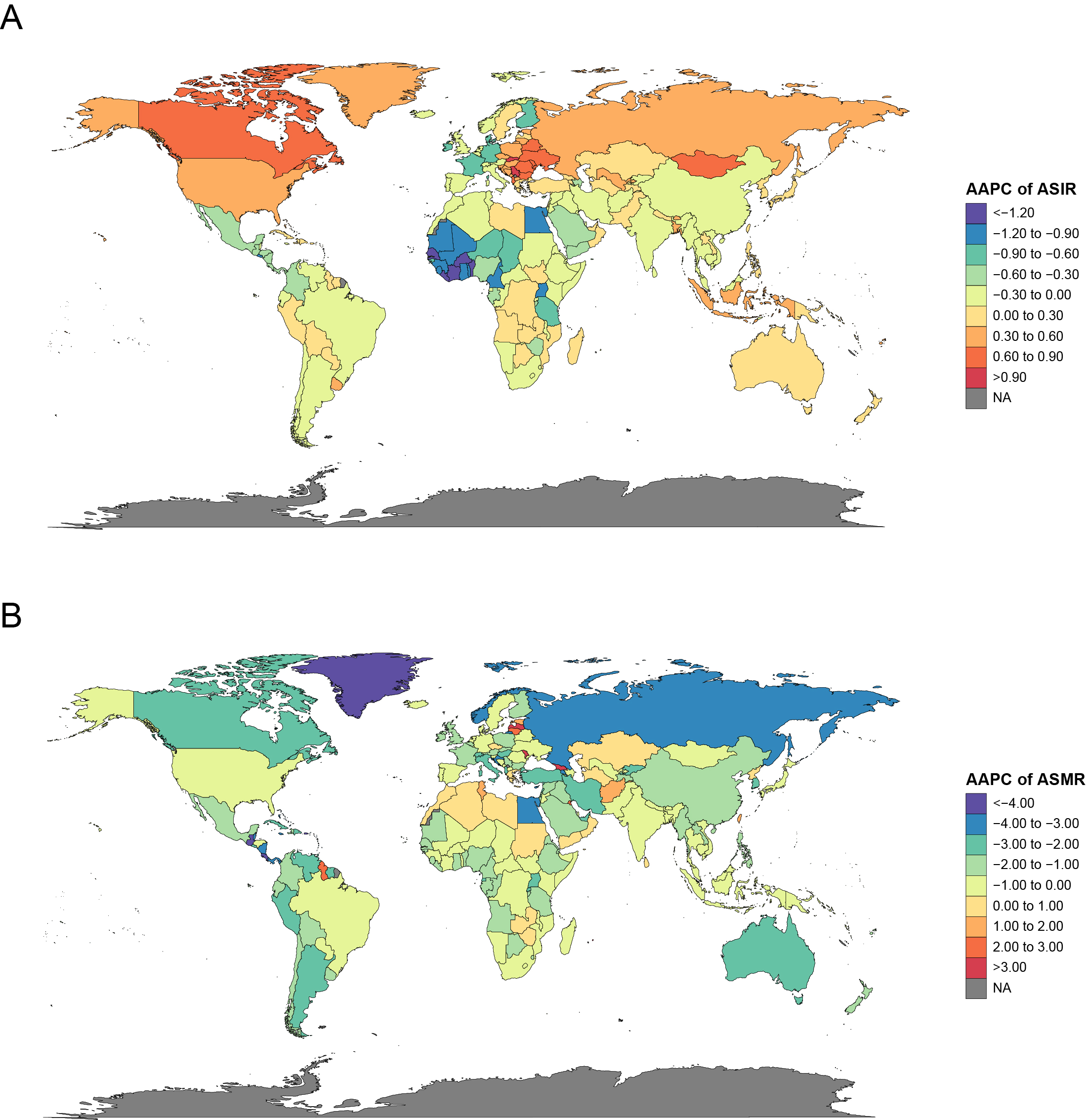

**Supplemental Figure 4. Annual trends of age-standardized incidence and mortality rates of PAH in 204 countries and territories from 1990 to 2021.** (A) AAPC of ASIRs. (B) AAPC of ASMRs. AAPC, average annual percent change.

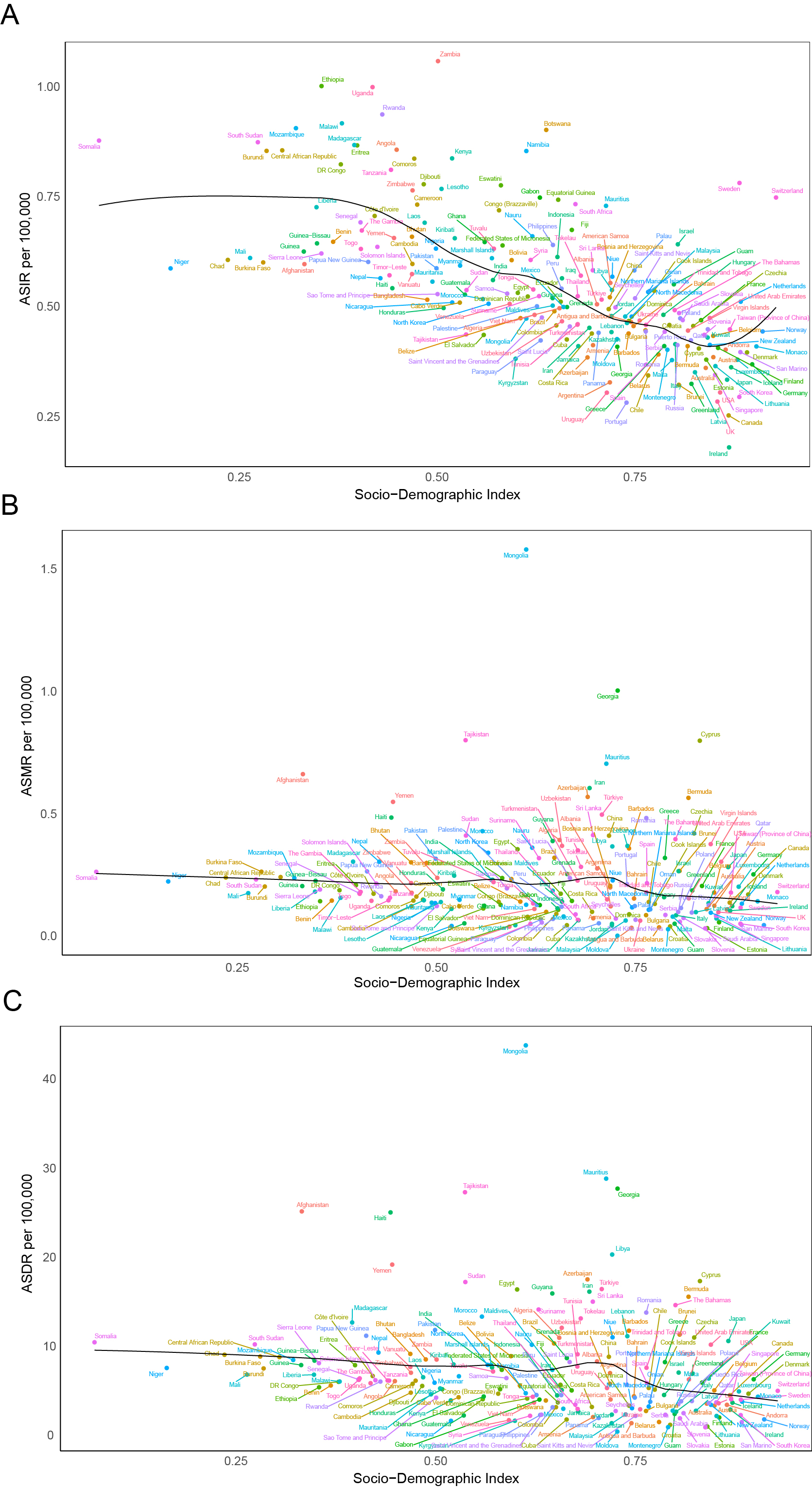

**Supplemental Figure 5. Age-standardized incidenc­­e, mortality and DALY rates of PAH by 204 countries and territories and Socio-Demographic Index in 2021.** The black line represents the expected values, and each point shows observed age-standardized rate for that specific country or territory. (A) Disease burden of ASIRs. (B) Disease burden of ASMRs. (C) Disease burden of ASDRs.
